# Family experiences during illness outbreaks: A systematic review

**DOI:** 10.1101/2022.11.16.22282428

**Authors:** J. Sheen, L. Chen, B. Lee, A. Aridas, A. Reupert

**Author notes:** Correspondence concerning this article should be addressed to first author J. Sheen.

## Abstract

**Background:** During the pandemic and associated lockdowns, many families from around the world experienced financial and confinement stress and the reorganisation of family caregiving responsibilities. Several studies have been conducted about the impact of the pandemic on family wellbeing. The aim of this systematic review was to identify, synthesize and critique relevant studies in this field.

**Methods:** Following Cochrane Collaboration and PRISMA guidelines, a systematic search was performed in databases including MEDLINE, PsycINFO, Embase, SocINDEX and PubMed. Peer-reviewed studies that examined the experiences of families during infectious disease outbreaks were included. Quality assessment was undertaken using the Mixed Methods Appraisal Tool. A narrative synthesis approach was employed.

**Results:** Eighty-four papers were found, all conducted during the Covid-19 pandemic, with the majority from the USA and presented from the perspective of parents/caregivers. Synthesized results focused on how family experiences, the dyad relationship and parenting behaviours were impacted during Covid-19.

**Conclusion:** Although some families reported positive growth, socially and financially vulnerable families were more negatively impacted than others during the pandemic. The review highlights the important role of families during times of stress and possible intervention targets.

---

The Covid-19 virus has led to over 591 million infections and at least six million deaths (WHO, 2022). The control measures employed to manage the spread of the virus, such as social distancing, self-isolation and quarantine, have resulted in significant changes to daily life around the world (WHO, 2020). Stay at home orders, also referred to as ‘lockdowns’, have also been widely employed to reduce virus transmission (WHO, 2020). For many, this has meant increased family time with little interaction outside of households (Sheen et al., 2021). It is critical to consider the wellbeing of families during the pandemic to inform practice, policy and further research. This systematic review aimed to explore available research examining the impact of infectious disease outbreaks on families in the 21^st^ century.

The mental health impacts of infectious disease outbreaks and associated restrictions on individuals has been previously researched. For example, in the USA, Adams-Prassl et al. (2020) found that stay at home orders were related to significant reductions in mental health, with the most adverse impacts reported by women. Another study, in Germany, found a deterioration in mental health over lockdown periods occurred in all age groups, though notably the elderly, socially isolated and those with pre-existing mental health conditions were most negatively impacted (Ahrens et al. 2021). A rapid review by Brooks et al., (2020) corroborated these findings, with evidence indicating that strict control measures can lead to post-traumatic stress symptoms.

Berger et al., (2021) extended this knowledge by reviewing literature on the psychological impacts of infectious outbreaks on children and adolescents. They found that young people experienced high rates of anxiety, depression, stress and loneliness during periods of isolation and quarantine, and those with pre-existing mental health difficulties fared worse. Similarly, Sprang and Silman (2013) found that during the H1N1 and SARS pandemics, families and children experienced high rates of Post-Traumatic Stress Disorder (PTSD) symptoms. The authors suggested that the control measures of isolation and quarantine contributed to the trauma of the experience. These studies establish a clear link between disease control measures and poor mental health, with findings consistent amongst adults, children and adolescents.

Families have their own way of responding to external threats. Within family systems theory, the family system is thought to be dynamic and changing over time (Minuchin, 1988). Change in the functioning of one subsystem is thought to result in changes in the functioning of other subsystems (Lucassen et al., 2021). Within this model, the changes in mental health noted above, have the potential to result in broader systemic impacts. For example, poor parental mental health may result in changes to other subsystems, such as the parenting relationship or the mental health of dependents. Protectively, healthy family functioning can play a vital role in helping individuals to cope with major life circumstances and stresses (Power et al., 2015). Understanding the complex and unique impacts of the pandemic on family systems is therefore vital. To date, no reviews identified have comprehensively addressed the impacts of infectious disease outbreaks including the COVID-19 pandemic, on family functioning.

Outside of pandemics, the impact of large-scale events such as natural disasters on families has been explored (Bolin, 1982; Cao et al., 2013; Pujadas Botey & Kulig, 2014; Lindgaard et al., 2009; McDermott & Cobham, 2012). The literature posits that families enter a period of adjustment in the face of a traumatic or adverse event (Bolin, 1989) with complex and varied impacts on family functioning (Lindgaard et al., 2009). To illustrate, Cao et al., (2013) found high levels of family dysfunction persisted eighteen months after the Wenchuan earthquake in China. Conversely, other studies have found increased cohesion and closeness and the creation of new family routines in the face of disaster (Bolin, 1982; Pujadas Botey & Kulig, 2014; Lindgaard et al., 2009). In their qualitative study of 19 families, Pujadas Botey and Kulig (2014) found that after the Slave Lake wildfires, parents described the importance of routines to promote family adjustment and noted a sense of appreciation for what was most important to their families.

There are some similarities that can be drawn from natural disasters and infectious disease outbreaks such as the widespread impact of the disaster, a sense of uncontrollability, financial implications and concern about loss of life, but there are important differences. Significantly, the measures taken to manage infectious disease outbreaks require individuals to isolate and remain in their immediate family group. Additionally, parents with dependent children have experienced unique burdens during the pandemic, including increased pressures and expectations to assume multiple roles (Aridas et al., 2021; Coyne et al., 2020; Sheen et al., 2021), changes in roles and routines, financial hardships and a reduction in personal time (Sheen et al., 2021). An increase in parental stress and harsh parenting practices (Prime, Wade & Browne, 2020) and undetected and unreported domestic violence (Campbell, 2020; Humphreys et al., 2020) is also emerging

This review aims to provide a comprehensive exploration of research examining the impacts of infectious disease outbreaks on family functioning in the 21^st^ century. Such information will provide an understanding of the impact of infectious outbreaks on families, including parenting, and the ways in which family members communicate, make decisions and express emotions to each other. Understanding more about how families function during this time can inform service improvement and policy. Given that there have been seven pandemics in the last 20 years (Reupert, 2021), it is critical that we learn from the past to achieve better outcomes in the future.

## Methods

This systematic review employed the method prescribed by the Cochrane Collaboration (Bero, 2017; Perry & Hammond, 2002). The review protocol was developed following the Preferred Reporting Items for Systematic Reviews and Meta-Analyses (PRISMA) model (Moher et al., 2009) and was registered on PROSPERO (CRD42020208995).

### Search Strategy

Five databases, MEDLINE, PsycINFO, Embase, SocINDEX and PubMed, were searched for research investigating the impacts of infectious disease outbreaks on families. The search terms and strategies were developed after consultation with a university librarian (see Table 1). Two searches were conducted; the first search captured papers published between 2002 and February 2021 when this review was initiated, and the second was for papers published between February 2021 and 22^nd^ March 2022 to provide an update of the review and ensure results were as recent as possible. Reference lists of included papers were searched manually to identify additional relevant papers.

**Table 1.**
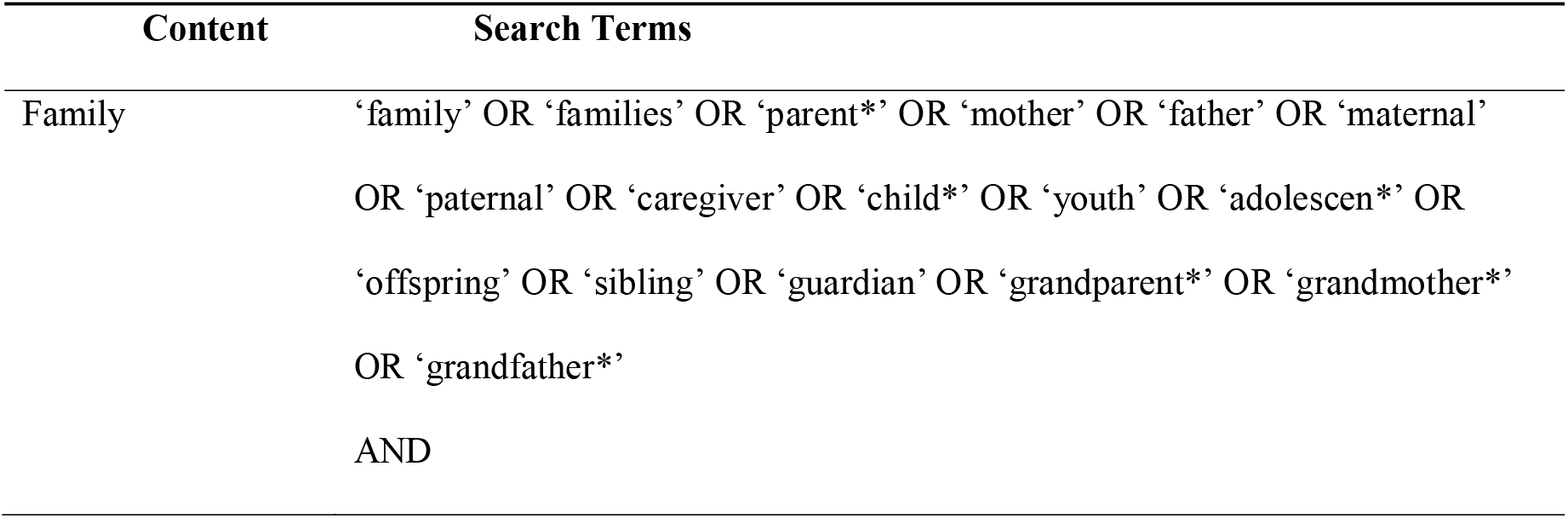
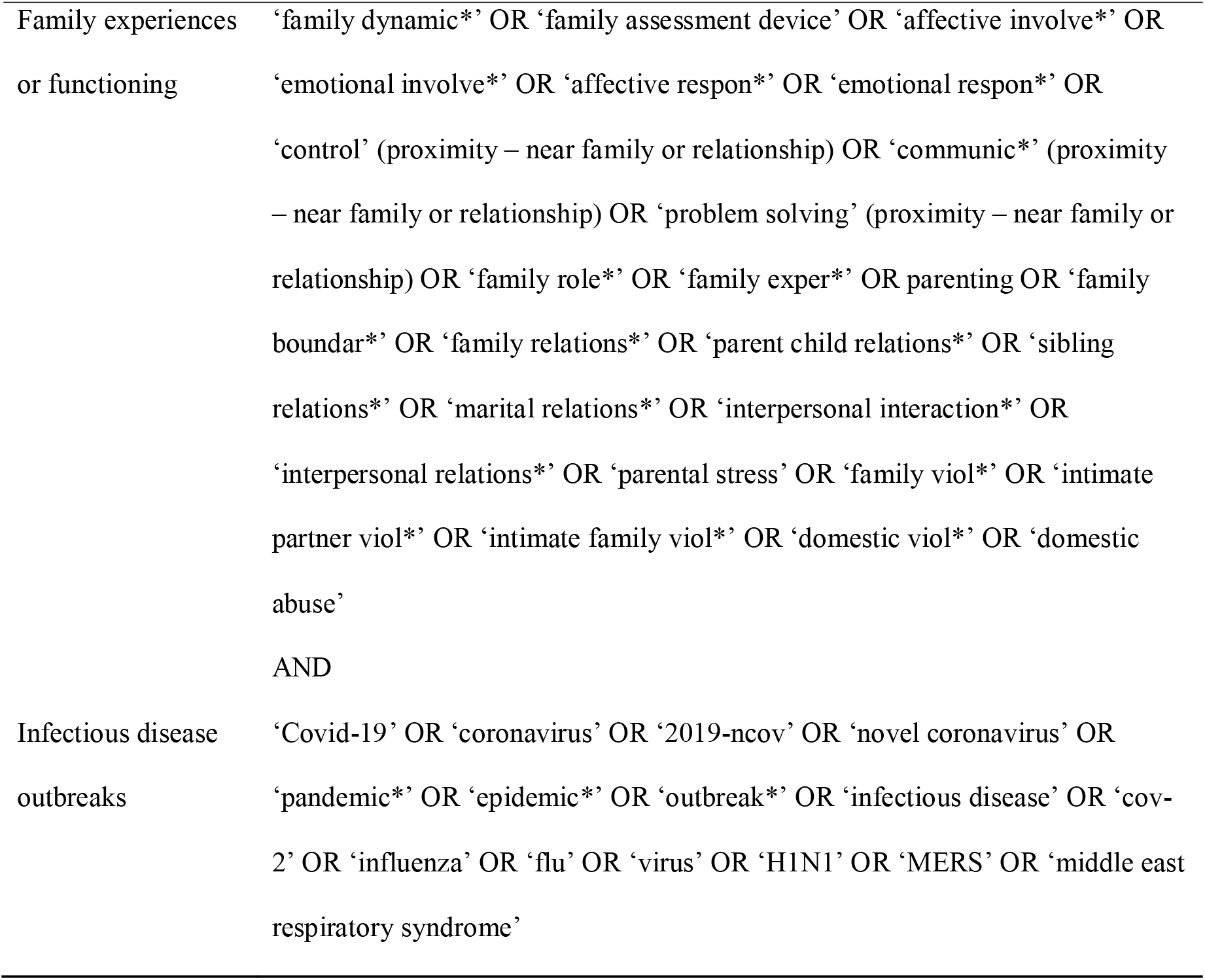
Search Terms Used Within the Databases

| Content | Search Terms |
| --- | --- |
| Family | 'family' OR 'families' OR 'parent*' OR 'mother' OR 'father' OR 'maternal'<br>OR 'paternal' OR 'caregiver' OR 'child*' OR 'youth' OR 'adolescen*' OR<br>'offspring' OR 'sibling' OR 'guardian' OR 'grandparent*' OR 'grandmother*'<br>OR 'grandfather*'<br>AND |
| Family experiences<br>or functioning | 'family dynamic*' OR 'family assessment device' OR 'affective involve*' OR<br>'emotional involve*' OR 'affective respon*' OR 'emotional respon*' OR<br>'control' (proximity – near family or relationship) OR 'communic*' (proximity<br>– near family or relationship) OR 'problem solving' (proximity – near family or<br>relationship) OR 'family role*' OR 'family exper*' OR parenting OR 'family<br>boundar*' OR 'family relations*' OR 'parent child relations*' OR 'sibling<br>relations*' OR 'marital relations*' OR 'interpersonal interaction*' OR<br>'interpersonal relations*' OR 'parental stress' OR 'family viol*' OR 'intimate<br>partner viol*' OR 'intimate family viol*' OR 'domestic viol*' OR 'domestic<br>abuse'<br>AND |
| Infectious disease<br>outbreaks | 'Covid-19' OR 'coronavirus' OR '2019-ncov' OR 'novel coronavirus' OR<br>'pandemic*' OR 'epidemic*' OR 'outbreak*' OR 'infectious disease' OR 'cov-2' OR 'influenza' OR 'flu' OR 'virus' OR 'H1N1' OR 'MERS' OR 'middle east<br>respiratory syndrome' |

The web-based software platform Covidence (Babineau, 2014) was used to screen the identified studies from the two searches. In the first search conducted in February 2020, 6883 records were identified after removing 15 duplicates, and 23 articles were included in the final review after title/abstract and full-text screening. Two additional articles were identified by manually searching reference lists of the 23 included articles. In the update search on 22nd March 2022, 222 records were identified after duplicates were removed. Forty-one articles were included for synthesis after the full-text screening, and their reference lists were manually screened, resulting in 19 additional articles meeting the inclusion criteria. As a result of the two searches, a total of 85 articles were included in this review. The PRISMA flowchart is presented in Figure 1.

**Figure 1.**
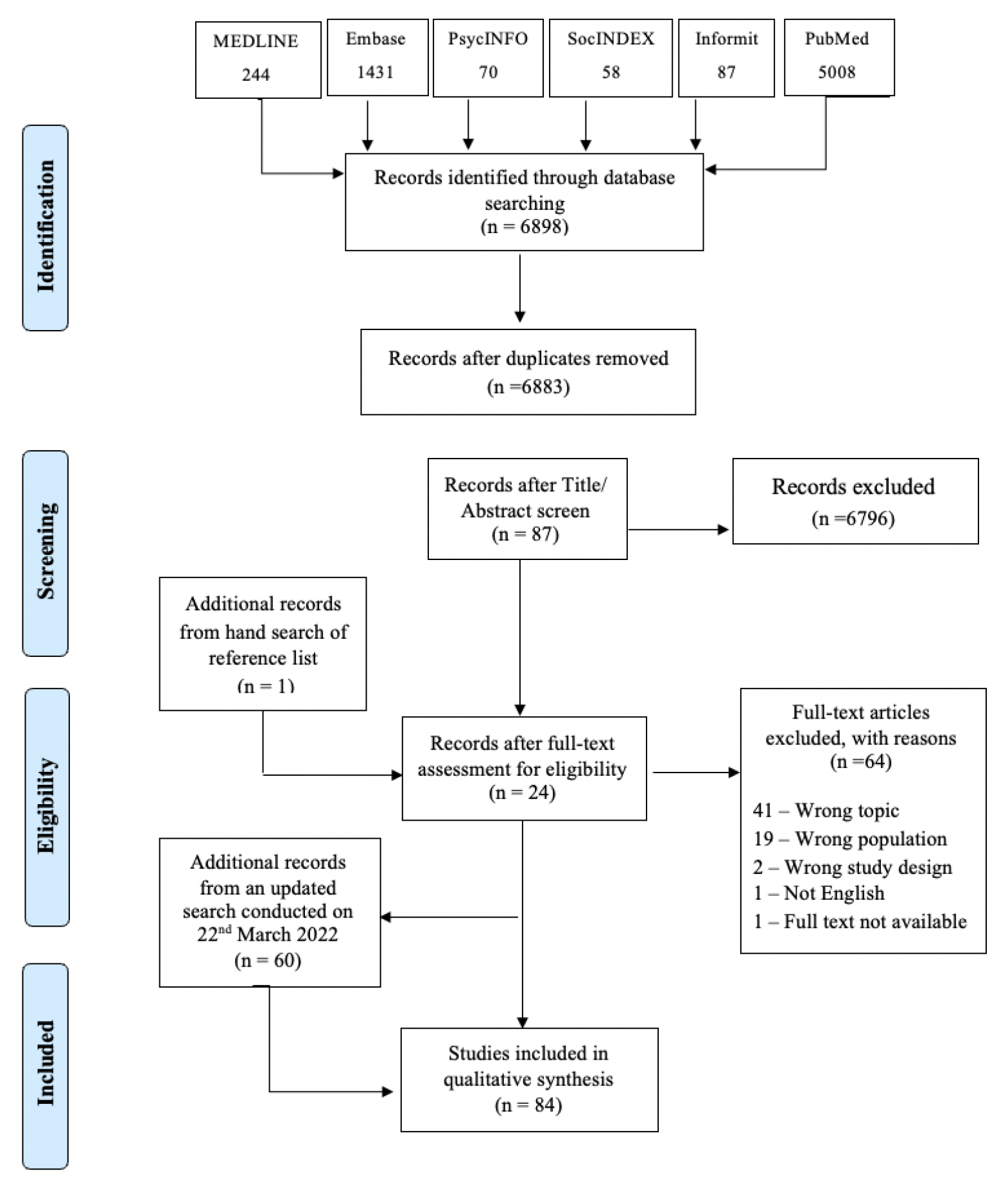
PRISMA Flowchart of the Study Selection Process

### Study Selection

This review included papers that were original empirical studies (not reviews), peer-reviewed, available in full text and published from 2002 onwards (as this marks the beginning of the SARS pandemic, considered to be the first pandemic of the 21^st^ century). Other eligibility criteria were studies that i) investigated respiratory outbreaks, ii) included participants who were parents or guardians of children under 18, and iii) examined at least one of the following domains of family dynamics, (a) family functioning, (b) family dynamics, (c) family communication, (d) family support, (e) family roles, (f) family experiences, and (g) parenting behaviours.

We focused on families with children under 18 as a discrete developmental period for children and parents. Our definition of family functioning and dynamics was drawn loosely from the Process Model of Family Functioning (Skinner et al., 2000; Steinhauer et al., 1984) and the McMaster Model of Family Functioning (Epstein, Bishop & Levin, 1978). Hence, we investigated general themes around i) roles (who does what in the family), ii) affective involvement and expression, iii) bonds, iv) communication, interactions and control, and v) parenting behaviours. These themes were not intended to be definitive but provided an initial structure for screening papers.

### Quality Assessment

The Mixed Methods Appraisal Tool (MMAT, Hong et al., 2018) was employed to assess identified papers. In this process, each study was assessed by two screening questions to decide whether it was an empirical study and then assessed against five criteria that varied for study type. To ensure the reliability of the assessment, three reviewers independently assessed five studies, with each study assessed by two reviewers. The details of the assessment criteria were clarified through discussion, and after an agreement was reached, the remaining 80 studies were assessed by one of the reviewers. As suggested by Hong et al. (2018), rather than calculating an overall score, a detailed presentation of each criterion rating was provided for each study (refer to Additional file 1).

The main issues for articles with a quantitative design (*n* = 73) included uncertainty in sample representativeness, unreported response rates and lack of information on the quality of measure employed. The three qualitative studies were of high quality, with the interpretation of results sufficiently supported by data and coherence found between qualitative data sources, collection, analysis, and interpretation. Quality issues for mixed-method studies (*n* = 9) included a lack of rationale for using a mixed-method design and inadequate integration of the qualitative and quantitative results.

### Data Extraction and Synthesis

A data extraction table was designed, including author/year, country, type of outbreak, research aim/s and research questions, sample demographics, methodology, time of data collection, analytic approach, family experiences, parenting, dyadic relationships, and other family relationships. Five papers were initially extracted by one reviewer and then cross-checked by two other reviewers to ensure that the required data were captured. Thereafter, the remaining 80 papers were extracted independently by one reviewer. (See online Supplementary Materials Table 1 for full summary of included papers) A narrative synthesis approach was employed to synthesize findings (Popay et al., 2006). First, qualitative and quantitative were analysed separately and then combined to present a narrative. Integral and representative extracts from identified papers were coded in response to the research question. The research team met regularly to discuss themes and ensure the final results were distinct. In-text references and quotations are presented to illuminate themes as appropriate, clearly distinguishing between primary data and the interpretations and conclusions drawn by researchers. Patterns within and across countries and different sample groups were examined and reported where found.

## Results

### Study characteristics

Eighty-four papers, including 85 separate studies (one paper included two separate studies; Rodriguez et al., 2021) conducted in 19 countries, were included in the final review. The majority were from the United States of America (USA; *n =* 29), with the remainder from mainland China (*n* = 10), Italy (*n* = 9), Canada (*n* = 5), the Netherlands (*n* = 5), Australia (*n* = 4), Germany (*n* = 4), Spain (*n* = 3), and other countries/regions (two each from Belgium, Israel and New Zealand; and one each from Bangladesh, Guatemala, Hong Kong, Ireland, Japan, Singapore, Slovakia, the United Kingdom, and Turkey). Seventy-three studies employed a quantitative design, nine employed a mixed-method design, and three used a qualitative design. Longitudinal designs, across different time points during the pandemic, were employed by 32 studies. All studies focused on the Covid-19 pandemic, and most (*n* = 72) were conducted between March and July 2020. Twenty studies used data collected before the Covid-19 pandemic as a point of comparison. Three studies compared data collected during and before the pandemic while others compared different child related variables e.g. children with a neurodevelopmental disability and typically developing children (Bentenuto et al., 2021) or when comparing children’s mental health status across different regions (e.g. Chen et al., 2020). Others investigated children and parents’ wellbeing during the pandemic (e.g. Cusinato, et al., 2020) or used correlation analysis to identify the association between different parenting variables and family functioning during the pandemic (e.g. Daks et al., 2020). Some examined child maltreatment and/or parenting in association with job loss and income reduction (e.g. Penner et al., 2021).

The sample size ranged from 12 (Gibbons et al., 2021) to 11,180 (Cao et al., 2021). Nearly half of the studies (*n* = 41) did not report sample cultural characteristics (e.g., race or ethnicity). Of the 44 studies reporting on participants’ race/ethnicity, 39 were conducted in Western countries (e.g., USA, Canada, Australia, New Zealand, Italy, etc.), where the majority of participants were Caucasians or non-Latinx White, with two exceptions including a predominantly Latinx group (Penner et al., 2021; Sun et al., 2021). The other five studies were conducted in other countries (China, Guatemala, Israel, and Singapore), with two investigating Han Chinese (Qu et al., 2021; Ren et al., 2021), one mainly examining mixed Indigenous and European descents (Gibbons et al., 2021), one focusing on a Jewish sample (Shorer & Leibovich, 2020), and the remaining one including a majority of Chinese and some Malay and Indian groups (Chung et al., 2022).

Thirty-nine studies were conducted during lockdown or where restrictions were in place and eight were conducted over a period encompassing both the lockdown/restricted period and the removal of restrictions. Examples of lockdown restrictions varied between papers in accordance with the rules of different jurisdictions and included school closures, reduced social access and reduced activity outside of the home. Five studies were conducted after lockdown. The remaining papers did not specify whether they were conducted during a lockdown or under restricted conditions.

Sixty-four papers did not report on the composition of the family. Of the 20 papers that reported family composition, one exclusively focused on extended households where grandparents were primary caregivers (Xu et al., 2022), and one had a specific focus on blended families where parents were in same-sex relationships (Goldberg et al., 2020). The other 18 included various types of families, most of which are two-parent families.

For studies that reported children’s ages (*n* = 80), five focused specifically on families with pre-schoolers, nine included school-aged children(Aznar et al., 2021; Chen et al., 2021; Dubois-Comtois et al., 2021; Hussong et al., 2022; McRae et al., 2021; Nyanamba et al., 2022; Ren et al., 2021; Rodriguez et al., 2021; Sun et al., 2021), 16 included adolescents and the other 50 studies included families with children in a wide age range (e.g., children under 12). Five studies investigated families where children had special needs, including children with neurodevelopmental disability (Bentenuto et al., 2021; Montirosso et al., 2021), externalising difficulties (Berry et al., 2021), Autism Spectrum Disorder (Colizzi et al., 2020), and children enrolled in a trial of preventive iron supplementation (Hamadani et al., 2020).

Synthesized results include how family experiences, dyad relationships, and parenting behaviours were impacted during the Covid-19 pandemic.

### Family experiences

Twenty-seven articles investigated family experiences during the Covid-19 pandemic, which highlighted, family cohesion and discord, communication, boundaries, family roles, and the impact of family experiences on children’s mental health and wellbeing.

### Family cohesion and discord

Family cohesion and discord were identified across countries, suggesting the experiences of the pandemic were dichotomous. For example, Chavez et al. (2021) reported that US households with children reported more conflict during the pandemic than prior, particularly when compared to households without children. These same families also reported greater family cohesion when compared with households without children. Günther□Bel et al. (2020) noted varied experiences amongst the 329 adults with romantic partners and/or children living through lockdown in Spain. Some participants identified increased family connection and support, illustrated by the development of shared goals, enjoying time together, improving extended family relations, feeling mutual support, and growing empathy and tolerance. Other participants in the study reported isolation and distance from family members, including their extended family (Günther□Bel et al., 2020). Having children at home was associated with improved family connections and less distance and empty nest families (i.e., families where children were no longer at home) reported comparatively less family connection and more distance (Günther□Bel et al., 2020).

Improved family cohesion and closer relationships was highlighted by Gibbons et al. (2021) in a qualitative study of 12 mothers of children under 7 years of age from Guatemala. These authors found that despite daily stressors related to the pandemic, there were also unexpected benefits including for some, personal and occupational growth, increased partner contributions and more time with their children. Rudolph et al (2021) similarly found increased family demands and increased satisfaction with family life in a sample of 1042 German parents.

Cassinat et al. (2021) explored the notion of ‘family choas’ and discord, noting that family chaos increased in their sample with the onset of pandemic-related shutdowns. These authors noted that with more noise, crowding and disruption, parents and children in the sample experienced difficulties in their relationships with one another.

Across studies, higher rates of discord and disrupted family functioning have been identified amongst socially vulnerable family groups, for example, single parents with young children (Rudolph et al, 2021); those experiencing economic stress (Peltz et al, 2021; Westrupp et al., 2021); families with children with neurodevelopmental disabilities (Westrupp et al., 2021) or mental health concerns (Polack et al., 2021); and, families with parental mental illness (Taha et al., 2022; Westrupp et al., 2021; Wu et al., 2021). Lower levels of family cohesion have also been associated with weekly spikes in health-related stress (Peltz et al, 2021), suggesting for some, a relationship between Covid-19 health fears and family functioning.

Positively, the pandemic appears to have increased parental knowledge and educational involvement in children’s’ lives (Cassinat et al., 2021); increased child autonomy and independence (Cassinat et al., 2021) even while curbing freedoms (Bülow et al., 2021); provided more ‘time in’ with children (Eales et al., 2021; Evans et al., 2020; Fioretti et al, 2020; Gibbons et al, 2021); and, an opportunity to ‘rediscover’ family (Fioretti et al., 2020). Further, positive life events served as buffers, supporting positive family functioning (Hussong et al, 2022).

### Communication

Gunter-Bel et al. (2020) observed shifts in communication patterns between family members during pandemic related lockdown. Some families reported frequent and higher quality conversations compared to the beginning of home confinement, including communicating their needs, and improving conflict resolution, while some experienced more conflict or identified old issues that resurfaced (Günther□Bel et al., 2020). Sociodemographic characteristics played an important role in the perceived quality of communication. Specifically, females and younger participants reported more family conflict than others; families identified as ‘empty nest families’ predicted less conflict; and those with higher education and who had been employed demonstrated improved communication compared to the beginning of lockdown (Günther□Bel et al., 2020). Looking at age and communication experiences through the pandemic, Polack et al. (2021) noted that younger adolescents experienced an increase in positive sibling interactions and communication.

### Boundaries

During lockdowns, families reported a balance between meeting individual and shared needs by respecting each other’s space and ensuring that there was individual and shared time (Günther□Bel et al., 2020). Other families experienced a lack of personal space and no collaboration (Günther□Bel et al., 2020). Parents with children at home were most likely to report what the authors described as unbalanced individual and shared needs (e.g., overwhelming childrearing responsibilities and no personal space), compared to couple-only families, divorced families, and empty nest families (Günther□Bel et al., 2020). The challenges associated with smaller living spaces and strict lockdown protocols appeared to factor into these discussions; the participants in an Australian study noted that they felt “trapped” and “stuck” in their homes, (Evans et al., 2020). Similarly, in a USA sample Eales et al., (2021) noted that families were challenged when having to live, work and school in one space. Although families in the sample were spending more unstructured and enjoyable time together, concerns regarding the lack of personal time/space were raised.

### Family roles

Family roles and the division of labour were highlighted across studies, particularly as the pandemic progressed. Some families reported that they distributed household tasks such as housework and childrearing (Günther□Bel et al., 2020). Other studies raised concerns regarding the distribution of labour and where studies reported that the housework and childcare associated with the pandemic typically fell on women (Del Boca et al, 2020; Evans et al, 2020; Frank et al., 2021; Gibbons et al., 2021), in some cases resulting in dissatisfaction and/or conflict (Craig & Churchill, 2021; Verweij et al, 2021). In a study of 274 US dual earner couples with young children, Shockley et al. (2021) found that of all the workload sharing models between couples identified during the pandemic, the model described as ‘remote wife does it all’ resulted in the lowest wellbeing and performance, whereas parents alternating work and parenting days resulted in the best overall strategy to preserve both wellbeing (for both parents) and maintain adequate job performance (only for mothers). In some families, children assumed additional chores to ease the burden on overworked parents (Evans et al., 2020; Goldberg et al, 2020).

### Family experiences impact children’s mental health/development

The impact of family experiences during the pandemic on children’s mental health/development was identified, with impacts across life stages explored. Amongst pre-schoolers, those with a more “harmonious family atmosphere” (not defined) and increased parent-child verbal communications were associated with fewer sleep problems (Liu et al., 2021). In adolescents, family discussions about the pandemic were associated with less depressive, anxiety, and stress symptoms, compared to adolescents living in families who did not talk about the pandemic (Tang et a., 2021). Young people with mental health concerns were associated with families who did not engage in joint family activities and where rules were not enforced during the pandemic (Ezpeleta et al., 2020). Conversely, children in families which had better relationships with parents and siblings and who experienced less parent-child conflict were less likely to experience mental health problems (Penner et al., 2021). Likewise, those families which reported greater parental ability to help children manage stress were less likely to have children experience mental health concerns (Penner et al., 2021).

### Dyad/couple relationships in families with dependent children

Nine papers reported on the dyad or couple relationship during the pandemic with three papers focusing exclusively on the dyad relationship in families with dependent children with specific themes identified below.

### Intimate partner violence

An increase in intimate partner violence during the pandemic was found across several studies. For example, a study of 2174 women living with their husbands and children in Bangladesh found increased intimate partner violence and feelings of being unsafe during lockdown when compared with data prior to the Covid-19 pandemic. Specifically, 19.9% of the sample reported emotional violence, 6.5% physical violence, and 3.0% sexual violence (Hamadani et al., 2020). In a sample of 618 Canadian parents, Gadermann et al (2021) identified parenting as a potential risk factor, noting 11.5% of the sample who identified as parents reported being concerned about physical or emotional domestic violence, significantly higher than the 7.9% of non-parents reporting similar concerns. Hostile sexism (defined as beliefs stipulating that men should possess social power and women are competing for men’s power, Overall et al., 2021), was identified as a risk for intimate partner violence. Overall et al. (2021) examined responses from 362 New Zealand parents in a longitudinal study, noting that men’s hostile sexism prior to the pandemic predicted residual changes in aggression towards intimate partners and children during the pandemic.

### Relationship quality

A study conducted in Spain found that the quality of couple relationships in families with children at home was significantly worse during lockdown while those without children at home appeared no worse and possibly better than pre-lockdown (Günther□Bel et al., 2020). However, as lockdown progressed, marital functioning for couples with children at home systematically improved while this was not the case for couples without children at home (Günther□Bel et al., 2020). Additionally, for parents, regardless of whether children were at home or had left home, telecommuting from home and being employed promoted positive relationship quality (Günther□Bel et al., 2020).

Examining 202 adoptive parents with school aged children in the US, Goldberg et al. (2020) noted that 70% of participants changed work situations during the pandemic, with most commencing work from home just as their children commenced home schooling. Although the division of labour was rarely identified as a stress between couples, the parent most involved in home schooling often reported increased stress and occasionally reported resentment towards their partner.

Two papers investigated the association between the quality of the dyadic relationship and individuals’ mental health outcomes during this time (Günther□Bel et al., 2020; Wu et al., 2020). In a sample of parents from China, Wu et al. (2020) found that parents with high levels of marital satisfaction and intimacy had lower rates of depression, anxiety and stress. These data suggest that the couple relationship may afford a type of buffering against distress. The results are not universal however, with Russell et al. (2021) noting that couple satisfaction was not a significant predictor of depression amongst their US sample.

Lockdown duration may also influence participant outcomes. Günther□Bel et al. (2020) found that relationship quality was negatively associated with individual distress and that this association strengthened as lockdown progressed. Westrupp et al. (2021) observed an increase in measures of couple verbal conflict during lockdown compared with prior to the pandemic.

### Parenting

Sixty-six papers described various parenting themes, including discipline, the parent-child relationship and the promotion of resilience.

### Discipline

A number of studies examined changes in parenting though the course of the Covid-19 pandemic. Amongst general population samples, some studies noted few if any significant changes in parenting (Donker et al., 2021; Penner et al., 2021; Verweij et al., 2021; Wong et al., 2021). To illustrate, Wong et al. (2021) found no change in the general prevalence of corporal punishment and aggression amongst parents from China during the pandemic. In the USA, Penner et al. (2021) reported little variation in adolescents’ perceptions of their parents’ stress, conflict and relationships with parents during the pandemic. In the Netherlands, Janseen et al. (2020) failed to find any difference in parental warmth, criticism or parenting behaviours in comparisons of data before and during the pandemic. The authors did highlight however, that there was significant individual variation between households. It is likely that general population studies may disguise the heterogeneity that exists in parenting practice.

By contrast, a Canadian study by Gaderman, et al. (2020) found that yelling and conflict increased during the pandemic compared with prior. Similarly, Russell et al. (2020) found greater conflict and reductions in feelings of closeness amongst US parents during the pandemic, compared with prior. Further, Rodriguez et al. (2021) observed a 20.3% increase in harsh discipline during the pandemic, a 24.9% increase in yelling and a 30.7% increase in conflict.

Income reduction and financial concerns were significantly associated with harsher parenting and in some cases, child maltreatment and abuse during the pandemic. Higher financial needs have also been associated with increased rates of child-parent relational conflict (Russell et al., 2020). The odds that parents who lost their jobs during Covid-19 would also psychologically maltreat their children were 4.86 times higher than those of parents who did not lose their jobs (Lawson et al., 2020) while financial distress doubled the odds of verbal aggression from parents in another sample (Rodriguez et al., 2021).

Negative emotions associated with parenting tasks appeared to prevent parents from engaging in effective parenting practices and may trigger poorer parenting patterns (Romero et al., 2020). Young parental age (Lawson et al., 2020), hostile sexism (Overall et al., 2021); parental depression (Brown et al., 2020; Lawson et al., 2020) and anxiety (Brown et al., 2020l; Romero et al., 2020) were associated with harsher parenting during the pandemic. Links were also been identified between parental stress and harsh discipline (Anzar et al., 2021; McRae et al., 2021) or coercive parenting (Lucassen et al., 2021), such that increases in parental stress were associated with increases in problematic forms of parenting. Perceived partner support was seen to potentially buffer these effects (McRae et al., 2021).

Higher levels of harsh parenting was associated with more child conduct problems, while higher parental harshness and lower warmth were associated with higher callous unemotional traits in children (Waller et al., 2021). Parental worries about contracting Covid-19 and a family member contracting Covid-19 were independently associated with higher child conduct problems (Waller et al., 2021).

Chung et al. (2022) asserted that Covid-19 indirectly impacted parenting by impacting parental stress. Some studies indicated that parents experienced higher stress during the pandemic both in samples with neurodiversity (Bentenuto et al., 2021; Montirosso et al., 2021) and neurotypical samples (Spinelli et al., 2020). Further, parents of children who experienced mental health difficulties experienced greater parental stress (Berry et al., 2021) and distress (Browne et al., 2021) than parents in non-clinical samples. Children with high rates of mental health difficulties experienced lower levels of parenting quality compared with siblings (Browne et al., 2021), suggesting a complex relationship between parental stress, children’s mental health and relationship quality (Essler et al., 2021). This relationship may also extend to grandparents acting in a kinship caregiving role, with Xu et al., (2021) reporting that parenting stress and mental distress were associated with elevated psychological aggression, corporal punishment and neglect towards their grandchildren.

### Parent-child relationships

Findings from studies regarding the impact of the pandemic on parent-child relationships varied. In a study of middle and high school students from China, Zhang et al., (2021) reported that Covid-19 improved parent child relationships. Conversely, a quarter of the Australian adolescents surveyed by Magson et al. (2021) reported that conflict with their parents had increased during the lockdown period, which was associated with decreases in life satisfaction. Amongst this cohort, increased conflict with fathers was associated with more depressive symptoms.

In a study of US parents, Feinberg et al. (2020) reported significantly worse individual wellbeing from pre-pandemic to pandemic measures and smaller but substantial declines in co-parenting quality and parenting quality. Similarly, Chen et al., (2020) found that parents experiencing lockdown in Wuhan were seen as less warm, more overprotective and more rejecting compared to their counterparts not experiencing lockdown. Low maternal relationship quality was positively associated with experiencing parent-child conflict as a burden during Covid-19 lockdown measures (Janssens et al., 2021).

Parents who reported clinically significant depressive symptoms experienced high anxiety and perceived stress, resulting in lower perceived closeness with children (Russell et al., 2021). Parental stress was also associated with higher rates of child distress (Spinelli et al., 2020; Wang et al., 2021), hyperactivity and impulsivity (Marchetti et al., 2020) and negatively associated with children’s social emotional competence (Wang et al., 2021). Protectively, mothers who receive support in their parenting role may experience better relationship quality and communication with their children than those who receive less support (Uzun et al., 2021).

Data regarding the impact of the structure of the parenting unit during the pandemic were mixed. Russell et al. (2020) found that partnered parents reported increased rates of child-parent relational conflict during Covid-19 compared to single parents. By contrast, Cusinato et al. (2020) found that single parenthood, a change in working routine, having a child with a disability and a high number of children living at home all adversely impacted parental stress during the pandemic (Cusinato et al., 2020).

### Multiple roles

Parents across several studies highlighted the impact of school closures (Bentenuto et al., 2021; Hiraoka & Tomoda, 2020); being responsible for meal preparation (Bentenuto et al., 2021; Colizzi et al., 2020); having less time to themselves (Bentenuto et al., 2021) and managing children’s time (Bentenuto et al., 2021; Colizzi et al., 2020; Eales et al., 2021), boredom (Gibbons et al., 2021) and disruptive behaviour (Colizzi et al., 2020), all of which was seen to increase parents’ stress and demands on their time. Excessive load and difficulties were reported by some parents (Evans et al., 2020), with a focus on managing multiple roles in lockdown including that of parent, teacher and therapist (Bentenuto et al., 2021). Balancing work or study with home schooling was seen as particularly burdensome and beyond the capabilities of many parents (Evans et al., 2020). Some Australian parents reported having to work late into the night in order to accommodate the demands of employment and home schooling (Evans et al., 2020), and parents in a USA sample experienced school closures as a “significant disruption” (Lee et al., 2021, p. 1).

Some studies investigated the impacts of home schooling on parent-child relationships. A study of UK parents found those using higher levels of self-blame experienced increased stress, in turn leading to worse home-schooling relationships (Anzar et al., 2021). Mothers interviewed by Gibbons et al. (2021) identified concerns regarding their ability to adequately support their child’s learning. Chen et al. (2021) reported that the impacts of home schooling may impact families disproportionately. In a study of 223 US parents, they noted that low and low middle-income earners and parents of colour experiencing more instrumental and financial hardships due to the pandemic, than other parents. These parents also reported more stress associated with structuring home learning environments and planning educational and physical activities at home for their children. (Chen et al., 2021).

For some, home schooling served as a source of relational conflict. Schmidt et al. (2021) noted that parents in their sample reported more negative parent-child interactions on days where children were working on school tasks. This effect was further amplified on days where parents were more heavily involved in learning. The length of home schooling also exacerbated concerns, with Deacon et al., (2021) noting that parental depression, anxiety and stress increased the longer children were being home schooled and where Walters et al., (2021) found that parental support for home schooling decreased over time. Of note, parents played an important role in children’s transition back to school, with Qu et al. (2021) reporting that youth in families with higher levels of parent–child conflict and lower levels of parent–child intimacy before schools reopened showed increased depressive symptoms and anger problems after school reopened.

### Positive interactions and resilience

Some parents saw lockdown as an opportunity to spend more time with their child(ren) (Bentenuto et al., 2021). This is supported by Gadermann et al. (2021), who reported that parents experienced increased positive interactions with their children during the pandemic, including having more quality time, feeling closer, showing love or affection to their children and observing increased resilience in their children.

Good parent-child relationships and resilience were seen as protective factors against anxiety and depression in adolescents during the pandemic (Cao et al., 2021), as were parental emotional warmth (Chen, Cheng, & Wu., 2020), paternal playfulness (Shorer & Leibovich, 2020) and social connection (Magson et al., 2021). Routine maintenance and specific parenting practices (e.g. focussed, soothing, structured) also correlated with positive child mental health outcomes (Romero et al., 2020).

For parents, resilience was expressed and developed through everyday activities such as going outdoors (Eales et al., 2021; Gibbons et al., 2021), talking to friends (Eales et al., 2021), developing new hobbies and new neighbourhood connections (Eales et al., 2021), engaging in mindful self-care (Gibbons et al., 2021) and a parental attitude of grit and determination to problem solve challenges (Eales et al., 2021). Moreover, parents who had a clear view of who they are as a parent and who consider parenting as inherently valuable, experienced greater protection against negative experiences and poor mental health during the pandemic (Schrooyen et al., 2021).

Factors that mitigated the risk of poor/harsh parenting included higher levels of confidence in managing behaviours designed to reduce Covid-19 transmission in children (e.g. handwashing and distancing)(Wong et al., 2021); financial stability (Lawson et al., 2020); a history of using reframing strategies (Lawson et al., 2020); routines that provided children with a sense of security and predictability (Liu et al., 2021); parental support (Brown et al., 2020) and perceived Covid-19 related control (i.e. the degree to which an individual believe they can reduce or manage the chances of falling ill from Covid-19) (Brown et al., 2020). Further, parents experienced less general stress when they were able to redirect their thoughts from the pandemic to more positive experiences (Aznar et al., 2021). The impact of pre-existing relationships between parents and their children in determining pandemic functioning was highlighted, and to a lesser extent, adolescents’ legitimacy beliefs with respect to pandemic rules and oppositional defiance (Bülow et al., 2021). Importantly, close relationships with parents predicated adolescents’ adjustment, irrespective of Covid-19 related stress, highlighting the critical nature of these relationships (Campione-Barr et al., 2021).

## Discussion

This review aimed to identify and synthesize previous research that explored the impacts of infectious disease outbreaks on the functioning of families with children under the age of 18. All identified studies focused on the Covid-19 pandemic, at varying times during the course of the pandemic including periods of lockdown and periods after lockdown though some (*n* = 10) did not specify a particular timeframe. Most studies were conducted in the USA (*n* = 29), with other countries commonly represented being Italy (*n* = 9), mainland China (*n* = 10), Canada (*n* = 5) and the Netherlands (*n* = 5). As well as China, other developing countries in which family studies occurred included Guatemala (Gibbons, et al., 2021), and Bangladesh (Hamadani et al., 2020), highlighting a missed opportunity to reflect on the experiences of families in other developing nations. Further, the majority of participants across studies were Caucasians or non Lantinx-White and most focused on two-parent families with one study only that included heterosexual and same-sex adoptive parents of school-age children (Goldberg et al., 2020). Thus, diversity amongst the populations recruited to date is limited both culturally and to a predominantly heteronormative perspective of family life.

Of the studies reported, many showed that families with dependent children were distressed but at the same time, felt more connected with each other during Covid-19. To illustrate, Gadermann et al., (2021) found that during the pandemic, parents with dependent children reported increased feelings of closeness and described showing more love to their family, but simultaneously, having more family conflicts, than prior to the pandemic. Similarly, families with children reported more conflict but more cohesion, than households without children (Chavez et al., 2021). Even though some parents felt trapped by stay at home conditions (Evans, et al., 2021) and lived in what Cassinat et al., (2021) described as chaotic living circumstances, for some families, the pandemic and associated lockdowns provided an opportunity for family connection (Eales et al., 2021; Evans, et al., 2021; Fioretti, et al., 2021; Gibbons, et al., 2021; Günther-Bel et al., 2020; Rudolph et al., 2021).

Numerous studies have demonstrated the extent to which family conflict is associated with negative outcomes for children and adolescents including but not limited to problem behaviours, substance misuse, depression and distress (for a review see Behar-Zusman et al., 2020). Moreover, high levels of family conflict contribute to a potentially negative family environment which serves as a stressor that interacts with, and can intensify, the negative effects of stressful life events (Avison, 2009) such as unemployment and financial insecurity arising from lockdowns. Nonetheless, this review found that similar to other family systems research, in times of stress, families with children have many strengths they may draw on (Turliuc et al., 2013). A family’s ability to address conflict in the event of a stressful situation may have what Chavez et al., (2021, p. 244) described as “reparative potential” that might contribute to post-traumatic growth (Bai & Repetti, 2015) and promote positive family functioning (Turliuc et al., 2013), which may help to explain the results of this review.

Thus, the nature of lockdowns increased the amount of time that families spent together, which although overwhelming for many parents and children, also provided opportunities for families to communicate, and engage in activities as a family, outcomes that were not necessarily reported with families without dependent children (Chavex et al., 2021) some of whom reported feeling distant from their extended families (as they would in lockdown; Günther-Bel et al., 2020). Whether these positive experiences are sustainable over long periods of lockdown is however questionable; Deacon et al., (2021) found parental depression, anxiety and stress increased the longer children were home schooled. In regard to other disaster research such as property loss, (Bonanno et al., 2010) found that though families may initially experience a phase of cohesiveness, when coupled with ongoing external stress or poor coping mechanisms, some experience a significant increase in family conflict.

Not all families reported positive experiences during the pandemic. Some families were more vulnerable than others, including single parents, those experiencing financial stress and unemployment, those with children with special needs or mental health issues and parents with their own pre-existing mental health issues (Polacak, et al., 2021; Peltz et al., 2021; Rudolph et a., 2021; Taha et al., 2022; Westrupp et al., 2021). Mothers in particular bore the brunt of the additional housework and childrearing responsibilities that occurred during the pandemic (Craig & Churchill, 2021; Del Boca, et al., 2021, Evans, et al., 2021; Frank et al., 2021; Gibbons, et al., 2021; Verweij et al., 2021) resulting in negative wellbeing impacts on women, and conflict in the dyad relationship (Craig & Churchill, 2021; Evans, et al., 2021). Moreover, during the pandemic, an increase in intimate partner violence was reported, and where parenthood was a potential risk factor (Gadermann et al., 2021). It would appear that those families that were vulnerable prior to the pandemic were particularly negatively impacted at this time.

The findings speak to the cumulative and dynamic impact of various stressors on families during Covid-19. To illustrate, parents facing unemployment or financial insecurity due to lockdowns, reported high stress (Oeltz, et al., 2021). In turn, other studies showed that parents experiencing stress tended to employ harsh and/ coercive discipline (Anzar etal., 2021; Lucassen et al., 2021; McRae et al., 2021) resulting in poor family relationships and child conduct problems (Waller at al., 2021). Likewise, parenting stress was associated with high rates of children’s distress, hyperactivity and impulsivity (Marchetti et al., 2020; Spinelli et al., 2020; Wang et al., 2021). Other research has highlighted that an accumulation of several risk factors, such as poverty and harsh parenting might impact negatively on children’s mental health and academic achievement (Wigelsworth et al., 2016). These findings are broadly consistent with the family stress model, which posits that socioeconomic disadvantage impacts child outcomes via its effects on parents (Acquah, Sellers, Stock & Harold, 2017; Gard, Mcloyd, Mitchell & Hyde, 2020). Based on the literature it is conceivable that increased financial pressures in some families due to shift reduction or job loss during the pandemic increased parental stress and harsh parenting, directly impacting the mental health of dependents in the family. The pandemic has clearly provided a global opportunity to explore these disproportionate impacts of trauma on families, highlighting a need for urgent attention and care to be directed towards socially and financially disadvantaged families.

Conversely, the review showed that a strong marital relationship, a previous positive child-parent relationship, family conversations about the pandemic and financial stability appeared to buffer the negative impact of pandemic for children. Others have also found that positive family experiences, and social companionship with family, are associated with fewer mental health problems for adolescents (O’Loughlin et al., 2017) which suggests that such experiences help to mitigate the stressors arising from the pandemic. The positive impact of close family ties is well established (Fuller-Iglesias et al., 2015), and the results of this review suggest that positive family experiences are beneficial in the context of a pandemic.

Supporting parents, and those who assume the parenting role, is a critical public health priority. The review findings highlight the need for societal level interventions that aim to eliminate social inequalities such as unemployment and financial insecurity. Other findings indicate that although many families experienced elevated stressors during Covid-19, psychoeducation on positive parenting, especially when stressed, and assistance given to help parents deal with children’s sometimes challenging behaviour and their mental health, especially their anxiety about the pandemic, are likely to be important intervention targets. Family discussions about the pandemic were shown to be a buffer for adolescents (Tang et al., 2021), similar to other findings which have shown family discussions about major life issues, such as parental mental illness, can promote positive family functioning (Reupert et al., 2015). Supporting parents in holding these discussions (the language to use, and how to respond to children’s questions) may also be needed.

### Limitations

Given the review focused exclusively on English language studies, there is a risk of bias in the findings presented. Additionally, a minority of studies sought the views of children and young people. Instead, most studies relied on parents’ reports of their children’s mental health and wellbeing. As children’s views about preferred supports is often very different from their parents (Maybery et al., 2005) it is imperative that further studies are conducted privileging children and adolescents’ voices and experiences, to better inform initiatives in this field. The experiences of families in developing nations, especially from Africa, is missing, as are the experiences of different family types (e.g. single parent families, same sex families, step families) and needs to be the focus of future research. Ongoing longitudinal studies, that continue to track families over time, including the years to come, would provide further clarification as to the potentially enduring impact of the pandemic on family members and the interrelationships between parenting stress, parenting, and children’s outcomes.

## Conclusion

This review presented the findings of recent research about family functioning during Covid-19. The disproportionate impacts of the pandemic were clear, with socially and financially vulnerable families bearing a significant burden. Families with fewer vulnerabilities were often able to reflect on positive growth and change, such as increased time together and a greater understanding of each other’s worlds. Over time however, the mental health and educational impacts appear to worsen, highlighting the delicate balance for policy makers caught between containing physical illness and supporting psychological health and wellbeing. This review highlights the essential role of the family unit in times of trauma. With support, family connection can be vital and protective. Without appropriate supports, distress can be heightened with ripples felt across the whole of the family system

## Data Availability

All data produced in the present work are contained in the manuscript

## Notes

**Author Note** I have no conflicts of interest to disclose. This review was registered on Prospero (CRD42020208995)

### Competing Interest Statement

The authors have declared no competing interest.

### Funding Statement

Preparation of this paper was supported by using award money from the Victorian
COVID-19 Research Fund-Stream B, State Government of Victoria. The funders of this
study had no role in study design, data collection and analysis, decision to publish, or
preparation of the manuscript.

