## Supplements for "Family experiences during illness outbreaks: A systematic review"

#### **Table. 1 Characteristics of Identified Studies**

| **Author/s (year), country** | **Aim/s** | **Sample** | **Time of data collection** | **Methodology and analytic approach** |
| --- | --- | --- | --- | --- |
| Aznar et al. (2021), the U.K. | To examine the effects of stress on parenting and home-schooling outcomes during Covid-19 lockdown between March and June 2020 in the U.K and to identify the factors that may buffer negative outcomes. | **Participants:** Parents who home-schooled at least one school-aged child **n = 183** **Age of children:** M = 8.18 (SD = 2.55) **Age of parents:** M = 41.51 (SD = 5.87) **Gender of parents:** 161 females, 10 males. **Living status:** 22% lived with one child, 53.8% lived with two children, and the rest lived with three or more children. 83.2% cohabited with the child’s other parent. **Household income**: M = £81,540.74 (SD = 63,331.37). **Time of helping children with schoolwork during lockdown:** M = 11.13 hr (SD = 7.28) per week. **Working status:** 66% were working from home and worked for 25.81 hr (SD = 14.23) per week at the time of lockdown while worked 17.04 hr per week (SD = 15.18) during the lockdown. **Race/ethnicity**: Not reported. **Parental educational level**: Not reported. | Between 1st May and 24th July 2020 (The U.K government imposed a national lockdown between March and June 2020, including closing schools; a further lockdown was imposed from January to March 2021) | **Quantitative design.** **Data collected:** 1) Demographics and household information. 2) Parents’ conﬁdence and enjoyment while home-schooling and parenting their children (researcher-developed questions). 3) Parent–child relationship during the covid-19 lockdown (researcher-developed questions). 4) Children’s discipline during the covid-19 lockdown (researcher-developed questions). 5) The Revised Child Rearing Practices Report (CRPR. 6) Stress questions (developed by the authors). 7) Cognitive Emotion Regulation Questionnaire (CERQ-Short Version). 8) The subscale “Self/Everyday Creativity” of Kaufman Domains of Creativity Scale (K-DOCS). 9) Creative Self-Efﬁcacy (researcher-developed questions).  **Analytic approach:** Principal component analyses, and hierarchical regression analyses. |
| Bacikova-Sleskova et al. (2021), Slovakia | To explore the changes in perceived maternal psychological control, autonomy support, and feeling of being overly controlled as a result of the Covid-19 lockdown in Slovakia. | **Participants:** Adolescents **n = 155** **Age**: M = 14.5 years. **Gender:** 60.4% females, 39.6 males. **Race/ethnicity:** Not reported. **Living status:** 1.8% living with a step-mother and others living with mothers. | The data for Time 1 were collected in February 2020 and the first week of March 2020, and the data for Time 2 was collected in May 2020 (during the 8th week of lockdown and the following weeks up to the 12th week). | **Quantitative design.** **Data collected:** 1) The maternal employment situation. 2) Perceived Parenting: The Psychological Control (the Psychological Control Scale-Youth Self-Report, PCS-YSR), Autonomy Support (the Perceptions of Parents Scale, POPS), and Feelings of Being Overly Controlled. 3) Quality of mother–child relationship: The Satisfaction with the Relationship with the Mother, and Disobeying Parents (the Disobeying parents subscale of the Problem behaviour scale).  **Analytic approach:** Descriptive analysis, correlation analysis, paired t-tests, and hierarchical multiple linear regressions. |
| Bentenuto et al. (2021), Italy | Investigated the adjustment of parents, couples and children with a neurodevelopmental disability (NDD) and those with typically developing (TD) children during the lockdown. | **Participants**: Parents of children with NDD and parents of TD children. **n = 164** (NDD group, n = 82; TD group, n = 82) **Age of parents (year)**: NDD group, M = 42.01 (SD = 6.59); TD group, M = 41.28 (SD = 6.39) **Gender of parents**: NDD group, 73 (89.0%) females, 9 (11.0%) males; TD group, 74 (90.2%) females, 8 (9.8%) males **Age of children (year)**: NDD group, M = 7.63 (SD = 3.77); TD group, M = 7.67 (SD = 3.86) **Gender of children**: NDD group, 20 (24.1%) females, 63 (75.9.0%) males; TD group, 23 (28%) females, 59 (72%) males. **Race/ethnicity**: Not reported. **Type of family**: Single parents, or mothers and fathers not living with their offspring or with the other parent of the child at the time of recruitment. | During the lockdown, not clear when the lockdown began and ended. | **Mixed-methods design. Data collected: 1)** Change in the amount of time spent by the parent doing activities with the child, children’s therapy situation. 2) Parental stress: The Parental Stress Scale (PSS). 3) Coparenting: the Coparenting Relationship Scale (CRS). 4) Child externalizing behaviour: Strengths and Difficulties Questionnaire (SDQ). 5) An open question about the most significant aspect of participants’ experience as a parent during the Covid-19 outbreak.  **Analytic approach:** 1) For quantitative data: 2 × 2 mixed ANOVAs, hierarchical regression analysis, and linear regression analysis; 2) For quantitative data: content analysis. |
| Berry et al. (2021), Ireland | This longitudinal cohort study aimed to examine the impact of the first wave of the Covid-19 pandemic in Ireland on parents of children with externalising difficulties, in comparison to parents of children without such difficulties. | **Participants:** Parents of children aged 4-16. **n = 159** (divided into two groups) **1) Clinical group:** Parents of children with externalising difficulties, n = 115. **Role of parents:** Mother, n = 109 (95.78%); father, n = 11 (4.35%); other, n = 1 (0.87%). **Marital status:** Married, n = 80 (69.57%); cohabiting, n = 12 (10.43%); separated, n = 12 (10.43%); single, n = 11 (9.57%). **Number of children:** M = 2.29 (SD = 0.915) **SES:** Professional workers, n = 17 (14.78%); managerial & technical, n = 46 (40.00%); non-manual, n = 16 (13.68%); skilled manual, n = 20 (17.39%); semi-skilled manual, n = 9 (7.83%); gainfully occupied, not stated, n = 5 (4.35%); unemployed, n = 1 (0.87%). **Children's diagnoses:** ADHD, n = 83 (72.17%); ASD, n = 37 (32.17%); Conduct Disorder, n = 4 (3.48%); Other, n = 31 (26.96%); None, n = 26 (22.61%).  **Race/ethnicity**: Not reported. **2) Non-clinical group:** Parents of children without externalising difficulties), n = 44. **Role of parents:** Mother, n = 38 (86.36%); father, n = 6 (13.64%). **Marital status:** Married, n = 34 (84.09%); cohabiting, n = 4 (9.09%); separated, n = 1 (2.27%); single, n = 2 (4.55%).  Underlying health problem: n = 5 (11.36%). **Number of children:** M = 2.189 (SD = 1.018). **SES:** Professional workers, n = 10 (22.73%); managerial & technical, n = 24 (54.55%); non-manual, n = 8 (18.18%); skilled manual, n = 1 (2.27%); semi-skilled manual, n = 1 (2.27%). **Children's diagnoses:** ADHD, n = 4 (9.09%); ASD, n = 3 (6.82%); Other, n = 3 (6.82%); None, n = 39 (88.64%).  **Race/ethnicity**: Not reported. | Data were collected at three time points during the Covid-19 pandemic in Ireland.  Time 1 data were collected during the Delay and Mitigation Phase (28 March 2020 to 18 May 2020), time 2 during the Reopening of Society Phase following Wave 1 (10 June 2020 to 19 July 2020) and time 3 during the Wave 2 Case Acceleration Phase (21 September 2020 to 21 October 2020). | **Quantitative design (longitudinal).** **Data collected:** 1) Demographic information. 2) The Strengths and Difficulties Questionnaire—Parent version (SDQ). 3) The Parental Stress Scale (PSS). 4) The Effects of Covid-19 Questionnaire (ECQ). 5) The Impact of Event Scale—Revised (IES-R). 6) The World Health Organization Well-Being Index (WHO-5). 7) The Brief Coping Orientation to Problems Experienced Inventory (Brief COPE).  **Analytic approach:** ANOVA and MANOVA. |
| Brown et al. (2020), USA | To examine the influence of risk and protective factors on parents’ perceptions of stress and risk of child abuse potential during the Covid-19 pandemic. | **Participants**: Parents with a child under 18 **n = 183 Age of parents (year)**: M = 35.37 (SD = 7.30, Range = 18-55) **Gender of parents**: 164 (89.6%) females, and 19 (10.4%) males. **Parent race/ethnicity**: Black/ African American, n = 9 (4.9%); Latinx, n = 39 (21.3%); Non-Latinx White, n = 122 (66.7%); Other or mixed race/ ethnicity, n = 13 (7.1%) **Parent Education**: Less than high school, n = 12 (6.6%); high school graduate/ GED, n = 30 (16.4%); some college, n = 40 (21.9%); associates degree or trade school, n = 23 (12.6%); four-year college degree, n = 53 (29.0%); post graduate degree, n = 25 (13.7%). **Relationship Status**: Single or partner not living in home, n = 39 (21.3%); married or partner living in home, n = 142 (77.6%). | From April 21st to May 9th, 2020. | **Mixed-methods design.** **Data collected**: 1) Covid-19 risk factors involving a checklist of stressors, with an open-ended question about how their life was affected. 2) Mental health risk factors: The General Anxiety Disorder-7 scale (GAD-7), and the Center for Epidemiologic Studies Depression Scale (CESD-R).  3) Protective factors: The parental support subscale from the Parent-Child Relationship Inventory (PCRI), the present control subscale of the Perceived Control Over Stressful Events Scale, and the acceptance subscale of the Cognitive Emotion Regulation Questionnaire-Short Form (CERQ/SF). 4) Primary outcomes: The Perceived Stress Scale (PSS-10), and the brief Child Abuse Potential Inventory (CAP Inventory).  **Analytic approach:** 1) For quantitative data, using descriptive analysis, correlation analysis, chi-square and one-way ANOVA analyses, and hierarchical multiple regression analyses. 2) Qualitative template analysis was used for qualitative data. |
| Browne et al. (2021), Canada | To explore longitudinal associations between child/parent mental health, parenting quality, and family functioning in relation to Covid-19 disruption over a two-month window of the pandemic. | **Participants**: Caregivers with children between 5 and 18 years old **n = 549** families with **1098** children  **1) Children demographics:**  **Younger child**: mean age = 9.62 (SD = 3.21), 45.9% female. **Older child**: mean age = 11.80 (SD = 3.32), 49.0% female. **2) Caregiver demographics:**  **Age**: M = 41.33 (SD = 6.33). **Gender**: Females, n = 372 (67.8%); male, n = 158 (28.8%).  **Ethnicity**: White-European, n = 332 (73%); White—North American, n = 56 (12.3%); Asian—South, n = 17 (3.7%); Asian—South East, n = 8 (1.8); Black—African, n = 10 (2.2%); Black—North American, n = 5 (1.1%); mixed, n = 12 (2.6%); other, n = 13 (2.8%); declined, n = 95 (17.3%).  **Relationship status:** Lone parent, n = 44 (8.0%); living apart couple, n = 8 (1.5%); married/common-law, n = 495 (90.2%); same-sex, n = 2 (0.4%). **Employment**: Full-time 276 (52.2%); part-time, n = 129 (24.4%); unpaid work, n = 93 (17.6%); unemployed, n = 14 (2.6%). | May (Time 1) and July (Time 2) 2020.  This study began two months into the pandemic, where increases in caregiver psychopathology were already reported. However, the restraints during Time 1 and 2 were not reported. | **Quantitative design (longitudinal)**. **Data collected:** 1) Disruptions related to Covid-19 including the effects of the pandemic on income/economics and pandemic-specific chaos. 2) The Kessler Psychological Distress Scale (K10). 3) Family functioning: The General Functioning Subscale of the McMaster Family Assessment Device (FAD-GF6+). 4) Parenting quality: The parenting practices scale from the 2014 Ontario Child Health Study. 5) Child mental health problems: The Patient Reported Outcomes Measurement Information System (PROMIS).  **Analytic approach:** Bayesian multilevel path analysis using MPlus 8. |
| Bülow et al. (2021), the Netherlands | To provide a preregistered examination to which extent parent-adolescent relationships among Dutch families were affected by Covid-19 LD measures, how families differ from each other, and which factors may explain differences in family risk and resilience. | **Participants**: Adolescents (n = 179) and their parents (n = 144) **1) Adolescents**  **Age**: M = 14.26 years (SD = 1.62, Range =12–17). **Gender**: 69% female. **Number of siblings:** 91% had at least one.  **Race/ethnicity**: Not reported.  **Type of family**: Not reported. **2) Parents** **Age**: M = 47.01 years (SD = 5.19, Range = 36–76). **Role of parents**: 117 (81%) biological mothers and 27 (19%) biological fathers. **Educational leve**l: 12% low-educated (<1% did not finish high school, 11% had high school diploma), 33% medium educated (vocational/technical training), and 55% highly educated (college or university degree).  **Race/ethnicity**: Not reported.  **Type of family**: Not reported. | Eight biweekly measurements waves were used, spanning 16 weeks.  Four assessments (January 2020 through February 2020) took place prior to the detection of the first Covid-19 patient in the Netherlands and start of the lockdown.  The assessments took place during the Covid-19 lockdown (end of March until mid-May 2020). | **Quantitative design (longitudinal).** **Data collected:** 1) Parental warmth and parent–adolescent Conflict: The Network of Relationships Inventory (NRI). 2) Autonomy support (measured by three scales): The parents’ promotion of adolescents’ volitional functioning, Psychological Control, and Behavioural Control. 3) Time with Parents and Peers. 4) Covid-19-Related Rules in the household. 5) Oppositional Defiance: The Oppositional Defiance Scale; Legitimacy Beliefs of Parental Authority. 6) Parental Anxiety and Worry (About Covid-19).  **Analytic approach:** Piecewise growth models in MPlus. |
| Campione-Barr et al. (2021), USA | To examine the associations between adolescents’ perceptions of positive and negative relationship qualities across four important close relationship partners on adolescent adjustment during the pandemic (controlling for pre-pandemic adjustment), and to examine the moderating role of Covid-related stress on this association. | **Participants**: Youth **n = 170** (including 67 pairs of siblings) **Age**: M = 16.21 (SD = 1.95, Range = 12–20) **Gender**: 86 females, 82 males and two did not identify as female or male. **Family income**: M = $70,000–$79,000/year (15% made <$40,000/year; 34% made more than $100,000/year). **Parental educational level**: 80% had at least one parent with a four-year college degree or more. **Type of family**: Not reported.  **Race/ethnicity:** European American, 80%; African American, 14%; Latinx, nearly 10%; American Indian/Alaskan Native, less than 5%; and Hawaiian/Pacific Islander, less than 5%. | Time 1 data (pre-pandemic) were collected during 2017–2018 as part of two separate and larger studies in Central Missouri and Southern Florida.  Time 2 data (during pandemic) were collected in June and July of 2020. | **Quantitative design (longitudinal).** **Data collected:** 1) Relationship quality (Time 2): Items selected from the Network of Relationships Inventory (NRI). 2) Anxiety symptoms at Time 1: The Revised Children’s Manifest Anxiety Scale (RCMAS) for Missouri sample and Anxiety subscale of the Depression Anxiety Stress Scale (DASS) for Florida sample. 3) Anxiety symptoms at Time 2 were measured by RCMAS for both Missouri and Florida sample. 4) Depressive symptoms at Time 1: The Centre for Epidemiological Studies Depression Scale (CES-D) for the Missouri sample and the Depression subscale of the Depression Anxiety Stress Scale (DASS) for the Florida sample. 5) Depressive symptoms at Time 2 were measured by CES-D for both Missouri and Florida sample. 6) Problem behaviour at Time 1: A scale developed by other researchers for the Missouri sample and the Problem Behaviour Scale (PBS) for the Florida sample. 7) Problem behaviour at Time 1: A scale developed by other researchers for both samples.  8) Covid-19-Related Stress: The Covid-19 Adolescent Symptom and Psychological Experience Questionnaire (CASPE).  **Analytic approach:** Descriptive analysis and structural equation path analyses. |
| Cao et al. (2021), mainland China | To assess the impact of only-child status on the mental health of adolescents confined at home during the Covid-19 outbreak. | **Participants**: Adolescents aged from 12 to 18 years (Grade 7 to Grade 9) **n = 11,180 Age of adolescents (year)**: M = 14.22 (SD = 1.101) **Gender of adolescents**: 5582 (49.9%) females, 5598 (50.1%) males. **Only-child state**: 2744 (24.5%) only child, 8436 (75.5%) non-only child. **Race/ethnicity**: Not reported. **Employment status**: Not reported. **Educational level of fathers**: Elementary school and below, n = 1026 (9.2%); middle school or senior school, n = 7892 (70.6%); college degree and above, n = 1926 (17.2%). **Educational level of mothers**: Elementary school and below, n = 1631 (14.6%); middle school or senior school, n = 7568 (67.7%); college degree and above, n = 1654 (14.8%). **Parent’s marital status**: 10,194 (91.2%) married, 276 (2.5%) remarried, 710 (6.4%) divorced or separated. | From March 20^th^ to 31^st^, 2020. | **Quantitative design.** **Data collected**: 1) The self-evaluated parent-child relationship (poor, general, or good. 2) Potential exposure risk of COVID-19: Answer the questions “Is any relative or friend infected with COVID-19?" and "Whether anyone in the community where you live is infected with COVID-19?" 3) Depressive symptoms: The Chinese version of the Patient Health Questionnaire for depression (PHQ-9). 4) Anxious symptoms: The Chinese version of the Generalized Anxiety Disorder 7-item (GAD-7). 5) Childhood maltreatment: The Chinese version of the Childhood Trauma Questionnaire (CTQ). 6) Resilience: The Chinese version of the Connor- Davidson Resilience Scale (CD-RISC).  **Analytic approach:** Descriptive analysis, Pearson Chi-square test, multivariate analyses of variance (MANOVA), and binary logistic regression. |
| Cassinat et al. (2021), USA | To explore how family chaos, parenting processes, parent–child relationship qualities, and sibling relationship qualities changed before versus the early months of the Covid-19 pandemic. | **Participants**: Families (parents and their adolescent-aged children) **Time 1 (between March 2019 and March 2020): n = 2,046 participants** (from 682 families**)**  **Time 2 (between May 1 and June 15, 2020): n = 1,622** **participants** (568 parents, 528 older siblings, and 526 younger siblings)  **Demographics (only Time 1 data were reported): Older siblings:**  Age: M = 15.67 (SD = .68) years  Gender: 51% female, 49% male, 1% transgender. **Younger siblings:** Age: M = 13.14 (SD = 1.11) years Gender: 48% female, 52% male. **Gender of sibling pairs:** 173 older sister– younger sister pairs, 172 older sister–younger brother pairs, 155 older brother–younger sister pairs, 180 older brother–younger brother pairs, and two sibling pairs with a transgender older sibling. **Sibling relationship**: 97% of siblings were biologically-related, 2% were step-siblings, and 1% were adopted siblings.  **Parental demographics:** **Age**: M = 45.15 (SD = 5.37) years. **Gender**: 85% mothers, 15% fathers, 1% transgender. **Race/ethnicity of parents:** 87% White, 9% Black/African American, and 4% other racial groups; 5% Latino.  **Marriage status**: 82% of parents were married.  **Parent-child relationship:** 97% were children’s biological parents, 1% stepparents, 1% adopted parents, and 1% other kin. **Marriage status:** 82% of parents were married. Employment status: 59% worked full-time, 23% worked part-time, 3% were looking for work, 3% were students, 10% were retired/not looking for work, and 2% were disabled.  **Family socioeconomic status:** Varying from working class to upper class, as indexed by parental education and household income. | Time 1 data collected between March 2019 and March 2020.  Time 2 data collected between May 1 and June 15, 2020, during the Covid-19 shutdown. | **Quantitative design (longitudinal).** **Data collected:** 1) Demographic Information (T1 & T2): 2) Family Chaos (T1 & T2): The 15- item Confusion, Hubbub, and Order Scale (CHAOS). 3) Parenting Processes: a. Parental Knowledge (T1 & T2) of children’s Behaviours and everyday activities: A scale developed by previous research. b. Parental Educational Involvement (T1 & T2): Items adapted from previous research. c. Autonomy Granting (T1 & T2): The 10-item Parent Behaviour Measure (PBM).  4) Parent–Child Relationship Qualities: a. Parent–Child Intimacy (T1 & T2) with both mothers and fathers: Relational intimacy item developed by previous research. b. Parent–Child Conflict (T1 & T2) with mothers and fathers: Items adapted from previous research. 5) Sibling Relationship Qualities: a. Sibling Intimacy (T1 & T2): The same questionnaire as Parent–Child Intimacy but the items were adapted to target siblings. b. Sibling Disclosure (T1 & T2): Items adapted from previous research. c. Sibling Conflict (T1 & T2): Items from the Revised Network of Relationships Inventory.  **Analytic approach:** A two-wave latent change score model. |
| Chavez et al. (2021), USA | To examine the impacts of Covid-19 on conflict and cohesion in households with children compared to households without children and to assess how family conflict and cohesion are related to social vulnerabilities in the context of the pandemic. | **Participants**: Households with and without children **n = 4,122** (households with children, n = 2,666; households without children, n = 1,456)  **Age**: M = 35.9 (SD = 10.5) for households with children; M = 35.7 (SD = 14.2) for households without children. **Gender**: 93.5% females among households with children; 88.2% female among households without children. **Educational level**: In households with children, 20.3% with a high school diploma or less, 26.1% with some college/university, 53.6% with a college/university degree. In households without children, 12.8% with a high school diploma or less, 22.0% with some college/university, 65.3% with a college/university degree.  **Age of children (in households with children)**: 43.0% of respondents have at least one child under 5, 51.2% have at least one child between ages 5 and 11, and 53.9% have at least one child between 12 and 18 years of age.  **Region**: In households with children, 52.9% from North America, 36.9% from South America, 0.7% from Europe, 8.4% from Africa, 0.2% from Asia and 1.0% from Oceania. In households without children, 65.4% from North America, 26.2% from South America, 0.6% from Europe, 7.2% from Africa, 0.3% from Asia and 0.4% from Oceania. **Employment affected by Covid-19**: In households with children, 56.0% stopped working, 72.3% working from home, 37.4% working in a job with high risk for Covid-19 exposure, 10.8% working in healthcare with direct patient contact. In households without children, 52.8% stopped working, 71.2% working from home, 38.6% working in a job with high risk for Covid-19 exposure, 14.3% working in healthcare with direct patient contact.  **Race/ethnicity**: Not reported. **Type of family**: Not reported. | Between April and September of 2020. | **Quantitative design.** **Data collected:** 1) Covid-19 Household Environment Scale (CHES), assessing two dimensions of family functioning (conflict and cohesion) and collecting socio-demographic information. 2) Household Social Vulnerability Score was calculated using indicators of unemployment, overcrowding, presence of an individual with caregiving needs, and low educational attainment. Each household was allotted up to 1 point per domain and the four domains were added to generate a total social vulnerability score.  **Analytic approach:** Descriptive analysis, bivariate analysis, confirmatory factor analysis and latent class analysis. |
| Chen et al. (2020), mainland China | To 1) investigate Chinese adolescents’ mental health status during the Covid-19 period and examine whether there is are significant differences in anxiety, depression, and parental rearing style when comparing adolescents from areas that have different levels of severity of Covid-19 (i.e., Wuhan vs. other cities in China);  2) examine whether sociodemographic (e.g., gender, grade in school, single child status) and pandemic related factors (e.g., parents’ involvement in Covid-19 related work) would demonstrate adolescents’’ significant differences in anxiety and depression. | **Participants**: Students in Grade 7 to 12. **n = 7,772** **Grade of students**: Grade 7 to 12. **Gender of students**: 4059 (52.23%) females, 3713 (47.77%) males **Race/ethnicity**: Not reported. **Type of family:** Not reported.  **Areas**: 2850 from Wuhan, 4922 from other urban areas (i.e., Beijing and Hangzhou). | From February 22^nd^ to March 8^th^, 2020 (when Wuhan was completely locked down). | **Quantitative design.** **Data collected**: 1) Anxiety symptoms: Generalized Anxiety Disorder-7 (GAD-7). 2) Depressive symptoms: Patient Health Questionnaire-9 (PHQ-9). 3) Perceived parental rearing style in adolescents: Short Egna Minnen Beträffande Uppfostran (S-EMBU)  **Analytic approach:** Descriptive analysis, independent sample t-test, Chi-Square test, and Structural Equation Modeling (SEM) using R. |
| Chen et al. (2021), USA | To examine the roles of income level and race/ethnicity in families’ experiences coping with the Covid-19 pandemic, focusing on those with school-aged children. | **Participants**: Parents of school-aged children **n = 223**  **Age of parents**: M = 41.31 years (SD = 8.54) **Gender of parents**: 202 (90.6%) females, 21 (9.4%) male.  **Race/ethnicity**: White/Caucasian, n = 145 (65%); Latin/Hispanic, n = 33 (14.8%); Asian and Pacific Islander, n = 16 (7.2%); Black/African American, n = 16 (7.2%); Mixed/Biracial/Multicultural, n = 7 (3.1%); American Indian and Alaska Native, n = 3 (1.3%); Other, n = 3 (1.3%).  Those who identified as Asian/Pacific Islander, American Indian/Alaska Native, Black/Africian American, Latinx/Hispanic, or mixed/biricial/multiracial were coded as people of colour (POC). **Family structure**: Single-father household, n = 3 (1.3%); single-mother household, n = 33 (14.8%); two-parent home, n = 181 (81.2%); other, n = 5 (2.2%); no response, n = 1 (0.4%) **Household income** (US Dollar): 18% low-income and lower-middle class (≤ $50,000), 23% middle class ($50,001–$100,000), and 54% upper-middle and high-income (> $100,000). **Employment status**: Full-time employed, n = 130 (58.3%); part-time employed, n = 23 (10.3%); self-employed, n = 20 (9.0%); homemaker, n = 18 (8.1%); out of work at the moment, n = 23 (10.3%); other, n = 6 (2.7%); no response, n = 1 (0.4%). | During a period of 2.5 months between April 8, 2020, and June 15, 2020. During this time,  the majority of schools in the US were closed and remained closed for the rest of the 2019–2020 school year. | **Quantitative design. Data collected:** 1) Demographic information. 2) Employment and job status: Whether participants’ and their partner’s employment status had changed since the pandemic and the degree to which they had worked remotely due to the pandemic. 3) Learning at home: Whether parents had clear structure and routines to guide their children’s learning at home and to assess the methods that children used to continue their education at home during the Covid-19 pandemic. 4) Consequences of school closure associated with the Covid-19 outbreak: Adapted items used in a 2009 influenza A (H1N1) study and four newly designed items.  **Analytic approach:** Descriptive analysis and Chi-square tests. |
| Chung et al. (2022), Singapore | To investigate how parents perceive the impact of Covid-19 on increased harsh parenting and reduced parent-child relationship closeness through the mediating effects of parenting stress. | **Participants**: Parents with at least one child aged under 12 **n = 258 Age of parents (year)**: 26-30, n = 13 (5%); 31-35, n = 77 (30%); 36-40, n = 83 (32%); 41-45, n = 48 (19%); 46-50, n = 31 (12%); 51-55, n = 6 (2%). **Gender of parents**: 165 (64%) females, 93 (36%) males. **Parent race:** Chinese, n = 212 (82%); Malay, n = 22 (9%); Indian, n = 11 (4%); More than one race/ others, n = 13 (5%).  **Parent education:** Non-University, n = 38 (15%); University, n = 220 (85%). **Parent Employment:** Stay-at-home parent, n = 33 (13%); Employed (full), n = 188 (73%); Employed (temporary/casual), n = 15 (6%); Unemployed (would like to work), n = 8 (3%); Others/Retired/Students, n = 14 (5%). **Youngest Child Age (year)**: 0–1, n = 63 (24%); 2–3, n = 54 (21%); 4–5, n = 47 (18%); 6–7, n = 31 (12%); 8–9, n = 33 (13%); 10–12, n = 30 (12%). **Type of family**: Not reported. | From April 22^nd^ to May 5^th^, 2020 (Circuit-breaker measures were imposed on April 21^st^, 2020, and was extended for a second month) | **Quantitative design.** **Data collected:** 1) The perceived impact of Covid-19: The shortened 6-item version of Coronavirus Impacts Questionnaire (CIQ). 2) Parental stress: The Parental Stress Scale (PSS. 3) Harsh parenting behaviours: Using similar items from large-scale studies in the US such as the Fragile Families Study. 4) Parent-child relationship closeness: Items were constructed based on the relationship quality dimension in the Child-Parent Relationship Scale. **Theoretical/conceptual framework:** The Parental Stress Model (Abidin 1992).  **Analytic approach:** Descriptive analysis, and mediation analysis in a structural equation modelling (SEM) framework using Mplus. |
| Colizzi et al. (2020), Italy | To 1) investigate the impact of the Covid-19 outbreak on individuals diagnosed with Autism Spectrum Disorder (ASD); 2) explore whether any pre-pandemic sociodemographic or clinical characteristics might predict a negative impact of the pandemic on ASD individuals’ wellbeing. | **Participants**: Parents and guardians of ASD individuals **n = 527** **Age of children (year)**: M = 13 (SD = 8.1) **Gender of children**: Not reported. **Race/ethnicity**: Not reported. **Employment status of parents**: 26.1% of mothers and 27.5% of fathers stopped working due to the emergency outbreak. **Type of family:** 88.2% were living in married or cohabiting couple families, 5.9% were separated, and 5.9% were single-parent families. | From April 6^th^ to 20^th^, 2020. | **Mixed-methods design. Data collected**: Impact of the Covid-19 outbreak on participants’ wellbeing, and needs to deal with the emergency.  **Analytic approach:** 1) Descriptive analysis, and logistic regressions were used for quantitative data. 2) For qualitative data, authors evaluated answers and pooled them into categories (e.g., healthcare, social, financial needs, etc.). |
| Connell and Strambler (2021), USA | To estimate household exposure to Covid-19 related stress and the association with parent report of neglectful, harsh, and positive discipline practices. | **Participants**: Caregivers of children under 18 **n = 2,068** (Wave 1, n = 1,019; Wave 2, n = 1,049. Wave 1 and 2 are two independent samples recruited at two different points) **Age of the focal child**: M = 8.2 years (SD = 5.2) **Gender of the focal child**: 49.0% females, 51.0% males. **Age of caregivers**: 18–24 years, 6.5%; 25–34 years, 28.9%; 35–44 years, 41.9%; 45–54 years, 18.2%; 55 years or older, 4.6%. **Gender of caregivers**: 63.2% females, 36.7% males.  **Race** **of caregivers**: White/Caucasian 75.3%; Black/African American 13.6%; Native American/American Indian 1.5%; Asian/Pacific Islander 6.3%; Other 5.5%.  **Ethnicity**: Hispanic, 13.2%. **Educational level of caregivers**: High school or lower, 19.9%; associate or bachelors, 51.7%; graduate degree, 27.8%. **Income**: Below $10,000, 6.2%; $10,000-$49,000, 32.3%; $50,000-$99,999, 27.7%; $100,000-$149,999, 15.2%; $150,000 or more, 15.7%; Prefer not to answer, 2.9 %. **Marital status**: Single (never married), 18.7%; married, 64.4%; cohabitating, 9.1%; divorced, 6.2%; widowed, 1.3%. | The survey was fielded on two separate occasions (wave 1 and wave 2, two independent samples), approximately 40 days apart in April and June 2020 (1–3 months after the onset of public health concerns related to Covid-19). | **Quantitative design. Data collected:** 1) Demographics. 2) Covid-19 Stress: a. A 12-item index reflecting personal and household member experiences of specific stressors due to Covid-19 or the public health response and an additional set of questions assessed whether the focal child had experienced school or childcare disruption. b. Covid-19 distress: 14 questions that measured the level of stress experienced due to Covid-19 concerns (e.g., “Difficulty finding childcare because of school/daycare closures”). 3) Parental neglect: Four subscales of the Multidimensional Neglect Behaviour Scale (MNBS), namely cognitive, supervisory, physical, and family-based neglect. 4) Parental discipline: Three subscales of the Parent-Child Conflict Tactics Scale (PC-CTS), namely positive (non-violent) discipline, psychological punishment, and physical corporal punishment.  **Analytic approach:** Chi-square analysis, t-test analysis, and logistic regression analysis. |
| Craig and Churchill (2021), Australia | To investigate how Covid-19 affected paid work, domestic work and care in Australian households and how gender differences in objective time spent on these activities, and in subjective feelings about it, changed due to the pandemic. | **Participants**: Dual-earner couples with children under 17 **n = 1,536**  **Age of parents**: Not reported. **Gender of parents**: Not reported.  **Age of children**: Not reported. **Gender of children**: Not reported.  **Aboriginal and Torres Strait Islander status**: Does identify as Aboriginal and Torres Strait Islander origin, 0.00 for men and 0.62 % for women; does not identify as Aboriginal and Torres Strait Islander origin, 100.00 % for men and 99.38% for women. **Household composition**: Living with an opposite-sex partner, 95.15% for men and 92.65% for women; Living with a same-sex partner, 8.00% for men and 9.67% for women; Living with trans or non-binary partner, 4.44% for men and 0.28% for women; Living with other family members, 18 years or older, 45.83% for men and 44.63% for women; Living with housemates, 2.44% for men and 7.34% for women; Living with children aged between 0 and 4, 77.27% for men and 75.23% for women; Living with children aged between 5 and 12, 90.43% for men and 87.25% for women; Living with children aged between 13 and 17, 57.75% for men and 64.44% for women. **Have persons in household who were sick**: 4.07% for men and 3.81% for women. **Have persons in household who needed assistance with activities**: 12.28% for men and 18.53% for women. Average number of people in household: M = 3.60 (SD = 0.1) for men and M = 3.68 (SD = 0.1) for women. **Average weekly earnings before Covid-19**: $1623.42 for men and 1324.13 for women. **Average weekly earnings during Covid-19**: $1563.01 for men and 1276.67 for women. **Average age of respondent:** M = 44.8 (SD = 0.8) % for men and M = 42.7 (SD = 0.2) **Country of birth**: Australia, 76.57% for men and 81.59% for women; other, 23.43% for men and 18.41% for women. **Location**: Urban, 84.09% for men and 80.41% for women; Regional, 10.80% for men and 14.55% for women; Rural, 5.11% for men and 5.03% for women. **Highest level of educational qualifications**: Bachelor’s degree or higher (incl. masters, PhD), 81.61% for men and 88.46% for women; Graduate diploma, graduate certificate, 8.62% for men and 6.18% for women; Certificate I, II, III, 5.75% for men and 3.50% for women; Year 12 or below, 4.02% for men and 1.86% for women. | Between 7 and 30 May 2020 (over a 3-week period during lockdown). | **Quantitative design. Data collected:** 1) Demographic information, and change in employment arrangements (before and after the advent of Covid-19). 2) Time allocation: Respondents were asked how many hours per day they spent in paid work, domestic work and care. 3) Subjective feelings about time allocation (before the COVID- 19-related restrictions were imposed and the time while the restrictions were in place).  **Analytic approach:** Descriptive analysis, t-tests, and chi-square tests. |
| Cusinato et al. (2020), Italy | To 1) investigate parents’ and children’s well-being, parental stress, and children’s resilience during the Covid-19 pandemic, especially during the quarantine. | **Participants**: Italian-speaking parents of children aged 5-17 **n = 463 Age of parents (year)**: M = 43.3 (SD = 5.88, Range =29-64) **Gender of parents**: 90.5% females, 9.5% males. **Age of the focal child (year)**: M = 9.72 (SD = 3.29)  **Gender of the focal child**: 56.2% male.  **Race/ethnicity**: Not reported. **Educational level of parents**: Primary school, n = 4 (0.9%); Secondary school, n = 34 (7.3%); High school, n = 201 (43.4%); University, n = 167 (36.1%); Post-university (including postgraduate specialization and PhD) , n = 57 (12.3%). **Working conditions:** 45.2% worked from home (teleworking), 27.6% were unemployed or fired, 21.2% had a temporary interruption to their job, and 6.0% worked part-time.  **Type of family**: 87.7% nuclear (i.e., two parents and children), 6.7% included a single parent (i.e., the children were being raised by one parent only), 4.1% were blended (i.e., a parent raises his/her children with a partner who is not their biological parent; the partner may, in turn, have children from a previous relationship) and 1.5% were extended (i.e., many relatives living together). | From April 25^th^ to May 8^th^, 2020 (containment measures started from March 9^th^ and ended on May 3^rd^, 2020). | **Quantitative design.**  **Data collected:** 1) Parent’s well-being: The Psychological General Well Being Index (PGWB). 2) Parental stress: The Parent Stress Scale (PSS). 3) Children’s psychopathological symptoms: The Strengths and Difficulties Questionnaire (SDQ). 4) Children’s resilience: The Child and Youth Resilience Measure (CYRM-R). 5) Parents’ perception of the relationship with their child, parents’ degree of satisfaction with the relationship.  **Analytic approach:** Descriptive analysis, reliability analysis and confirmatory factor analysis (CFA), T-test, general linear models (GLM), multiple linear regression, analysis of variance (ANOVA), and correlation analysis. |
| Daks et al. (2020), USA | To examine the links between parents’ psychological flexibility/inflexibility and family functioning in the midst of the upheaval associated with the Covid-19 pandemic. | **Participants**: Parents **n = 742 Age of parents (year)**: M = 40.7 (SD = 8.13) **Gender of parents**: 71% females, 27% males, 1% transgender, 1% other. **Race of parents**: 84% Caucasian, 5% African American, 4% Latino/Hispanic, 2% Asian/Pacific Islander, 2% Native American, and 3% other/biracial.  **Age of children (year)**: M = 9.4 (SD = 5, Range =5-18) **Gender of children**: 50% females, 50% males.  **Marital status**: 97% in romantic relationships, 85% were married/engaged, 11% in committed relationships, and 3% single/dating. **Educational level of parents**: 4% had only a high school level of education, 21% completed some college or trade school, 32% had bachelor’s degrees, and 43% had graduate degrees.  **Working conditions during the pandemic:** 70% work from home (39% full-time) in the last week. **Type of family**: All co-parents were “raising the child with the help of another adult living in the home” (p. 19). | From March 27^th^ (after roughly 10 days of press briefings by the White House Coronavirus Task Force and after over half of the states had enacted formal stay-at-home orders) to the end of April 2020. | **Quantitative design.** **Data collected**: 1) Psychological flexibility/inflexibility: The 60-item Multidimensional Psychological Flexibility Inventory. 2) Family discord: The Confusion, Hubbub, and Order Scale (CHAOS). 3) Coparenting discord: The Coparental Interaction Scale. 4) Family cohesion: The Family Assessment Device scale (FAD). 5) Caustic Parenting: The Parenting Practices Questionnaire (PPQ) and the Alabama Parenting Questionnaire (APQ). 6) Constructive parenting: Same questionnaire with Caustic Parenting. 7) Child distress: The Child behaviour Checklist (CBCL). 8) Parent depressive symptoms: The Patient Health Questionnaire (PHQ-9); 9) Covid-19 risk (researcher-developed questions): Three items to assess perceptions of oneself and/or their social/familial network’s risk of contracting Covid-19. 10) Stress from new demands (researcher-developed questions): Three items to assess stress resulting from new work and parenting demands.  **Analytic approach:** Descriptive analysis, correlation analysis, and path analysis using Mplus. |
| Deacon et al. (2021), Canada | To examine whether home schooling during the Covid-19 lockdown was associated with couples’ mental health and substance use outcomes and whether this association is differently for men and women. | **Participants**: Romantic couples **n = 758** romantic couples (n = 1,516 individuals) **Age:** M = 54.7 years (SD = 13.9).  **1) Non-homeschooling (n = 1094):** **Gender**: 48.18% female, 51.55% male, 0.18% non-binary, 0.09% unknown. **Ethnicity**: 82.08% White, 11.97% Asian or Arab / West Asian (e.g., Armenian, Egyptian, Iranian, Lebanese, Moroccan), 1.56% Latin American, or Black or First Nations, 2.19% multiracial, 1.46% other, 1.10% unknown. **Relationship length**: M = 30.24 years (SD = 15.32). **Relationship status**: 93.60% mixed sex, 6.40% same sex. **Marital status**: 98.91% married/ Common Law, 1.09% in a serious relationship. **Employment status**: 26.60% full-time, 10.24% part-time, 55.85% unemployed/students, 7.31% unknown. **Highest level of education**: 0.91% elementary school, 4.19% some high school, 17.85% high school graduate, 18.12% some college/university, 43.08% college/university graduate, 4.19% some post-graduate, 11.39% post-graduate degree (e.g., Master’s, Ph.D., LLB, MD), 0.27% unknown. **COVID Diagnosis**: 96.36% not having a diagnosis, 3.19% suspected and recovered, 0.18% Yes – diagnosed and recovered, 0.27% Yes – diagnosed and still ill.  **2) Homeschooling (n = 422):** **Gender**: 50.24% female, 49.53% male, 0.24% non-binary. **Ethnicity**: 59.95% White, 30.57% Asian or Arab / West Asian (e.g., Armenian, Egyptian, Iranian, Lebanese, Moroccan), 4.27% Latin American or Black or First Nations, 1.66% multiracial, 2.37% other, 1.18% unknown. **Relationship length**: M = 18.68 years (SD = 8.85). **Relationship status**: 94.76% mixed sex, 5.24% same sex. **Marital status**: 99.05% married/ Common Law, 0.95% in a serious relationship. **Employment status**: 59.00% full-time, 8.06% part-time, 27.01% unemployed/students, 5.93% unknown.  **Highest level of education**: 095% some high school, 10.66% high school graduate, 7.82% some college/university, 52.13% college/university graduate, 5.45% some post-graduate, 22.98% post-graduate degree (e.g., Master’s, Ph.D., LLB, MD).  **COVID diagnosis**: 92.18% not having a diagnosis, 6.87% suspected and recovered, 0.71% Yes – diagnosed and recovered, 0.24% Yes – diagnosed and still ill. | Survey questions were about experiences during April 2020 (during the lockdown) but the data collection was completed in July 2020. | **Quantitative design.** **Data collected:** 1) Homeschooling Assessment, including demographic and relationship variables, COVID diagnosis, and time spent homeschooling in hours per week. 2) Emotional Responses: The Generalized Anxiety Disorder scale (GAD-7), the Patient Health Questionnaire (PHQ-9), and the short versions of two COVID Stress Scales – Socioeconomic and Traumatic Stress scales. 3) Behavioural Responses (Coping-related cannabis use and coping-related alcohol use): The Brief Alcohol Motives Measure and the Brief Cannabis Motives Measure. 4) Optimism: The Life Orientation Test- Revised (LOT-R).  **Analytic approach:** Descriptive analysis, association analysis, and the actor–partner interdependence model (APIM). |
| Del Boca et al. (2020), Italy | To investigate the effect of Covid-19 on work, housework and childcare arrangements of women and their male partners, both working before Covid-19. | **Participants**: Women from dual-worker families before the COVID outbreak **n = 520** (women with children, n = 350; women without children, n = 170)  **Age of women**: M = 44 years (SD = 9.21, Range = 26-64) **Gender**: All females. **Race/ethnicity**: Not reported. **Educational leve**l: 47% having a degree. **Having children**: 67% having children. **Number of children** among those living with a partner and at least one child: M = 1.66 (SD = 0.74). Number of children aged 0–5: M = 0.36 (SD = 0.59). Number of children aged 6–10: M = 0.37 (SD = 0.53). Number of children aged 11–14: M = 0.25 (SD = 0.47). Number of children aged ≥15; M = 0.68 (SD = 0.89). **Employment status during the pandemic**: 1) Working in their usual workplace: 23% for women and 33% for their male partners. 2) Working from home: 44% for women and 30% for their male partners. 3) Stopped working because of the emergency: 33% for women and 37% for their male partners. | In April 2020 (during the first phase of the emergency).  At the beginning of March 2020, the Italian government imposed drastic measures to contain the growing epidemic including a lockdown on activities and public services, and school closures. | **Quantitative design. Data collected:** 1) A questionnaire to gather information on changes in the respondents’ employment status, hours of work, childcare, income and satisfaction regarding their work and family during the emergency. 2) A set of ad-hoc questions regarding the time spent on housework and childcare before and after the Covid-19 outbreak. 3) Women were also asked similar questions about their partners.  **Analytic approach:** Descriptive analysis and multivariate regression analysis. |
| Donker et al. (2021), the Netherlands | To examine changes in parenting and parent-adolescent relationship quality from before Covid-19 till during the Covid-19 pandemic in a sample of Dutch adolescents and their parents; and to investigate the associations between perceived stress and these changes, as well as the potential moderating role of pre-existing coping tendencies on the association between stress and changes in relationship quality. | **Participants**: Parents and adolescents   **Time 1 (Fall 2019)** **n = 240** adolescents and **236** parents **Age of parents**: M = 44.2 years (SD = 5.04). **Age of children**: M = 11.4 years (SD = 0.50). **Gender of parents**: 85% mothers. **Gender of children**: 120 (50%) girls and 118 (49.2%) boys. **Living status**: 80.4% adolescents living with both parents, 12.9% not living with both parents, and 6.7% missing. **Monthly family net income**: Median = 4000–4500 euros/month (SD = 4.44, about 2220 euros).  **Race/ethnicity of parents or children**: Not reported.  **Time 2 (Spring 2020)** n = 190 adolescents and 195 parents  Demographics not reported. | Pre-Covid-19 (Fall 2019) and the Covid-19 period (Spring 2020).  Schools were closed from halfway March till the beginning of May in the Netherlands. Large gatherings were forbidden during the whole Spring. Parents were mostly working from home during this period. | **Quantitative design (longitudinal)** **Data collected:** 1) Demographics including monthly family net income and perceived SES. 2) Parent-Adolescent Relationships: a. Parent-Adolescent Relationship Quality (T1 & T2): The Network of Relationships Inventory (NRI). b. Parenting (T1 & T2): The Alabama Parenting Questionnaire (APQ). 3) Parent and Adolescent Stress (T2): a. Four items assessing the amount of worrying because of the Covid-19 measures. b. Life Experience Survey: Parents and adolescents answered whether they experienced a large range of life events and how positive or negative the event was for them (48 items for adolescents and 47 items for parents). c. Parent and Adolescent Coping (T1): The Utrechtse Coping List (UCL).  **Analytic approach:** Descriptive analysis, correlation analysis, Confirmatory Factor Analyses, and univariate Latent Change Score models |
| Dubois-Comtois et al. (2021), Canada | To assess how multiple factors from different ecological levels (sociodemographic, family, parent, parent-child, and child) relate to child adaptation during the Covid-19 lockdown, from both parents’ and children’s perspectives. | **Participants**: Children and their parents **n = 144** parent-child pairs **Age of children:** M = 10.44 years (SD = 1.09; Range: 9-12). **Gender of child**: 70 (48.6%) females, 74 (51.4%) males. **Age of parents**: M = 40.1 years (SD = 5.11; Range: 27-59). **Gender of parents:** mother, n = 131, fathers, n = 13. **Race/ethnicity of parents**: Not reported. **Educational level of parents**: 12.5% had a high school degree, 34.7% a college or professional degree, and 52.8% a university degree. **Family income before Covid-19**: M = 98,265 $CAN (SD = 64,416; Range = 10,000-450,000 $CAN). 23.6% less than 50,000 $CAN, 13.9% between 50,000 $CAN and 70,000 $CAN, 29.9% between 70,001 $CAN and 110,000 $CAN, and 31.3% more than 110,000 $CAN. Two families refused to disclose their annual income. **Location of residence**: Families came from 14 of the 17 administrative regions in the province of Quebec and lived in metropolitan, suburban, or rural areas. | Between April 18, 2020, and May 18, 2020, in various areas in Quebec, where the lockdown was mandatory. | **Quantitative design.** **Data collected:** 1) Sociodemographic Characteristics. 2) Parent Depressive Symptoms (parent-report): The Brief Symptom Inventory-18 (BSI-18). 3) Family Functioning (parent-report): a. Household disorder: The Confusion, Hubbub, and Order Scale (CHAOS). b. General family functioning: The short version of the General Functioning subscale of the McMaster Family Assessment Device (GF61). 4) Parent-Child Relationship and Security of Attachment: a. Quality of the parent-child relationship (parent-report): The Child-Parent Relationship Scale (CPRS)—Short Form. b. Security of attachment (child-report): The short version of the Inventory of Parent and Peer Attachment (IPPA-R). 5) Child Aversion to Aloneness (child-report): The Negative Experienced Aloneness Scale of the Loneliness and Aloneness Scale for Children and Adolescents. 6) Parent and Child Anxiety Toward Covid-19 (parent- and child-report): The Fear of Covid-19 Scale (FCV- 19S). 7) Child Behaviour Problems since the beginning of the lockdown: a. Parents completed the Externalizing Scale of the Child Behaviour Checklist for Ages 6 to 18 (CBCL/6–18). b. Child internalizing problems were self-reported using the Youth Self-Report (YSR).  **Analytic approach:** Descriptive analysis, correlation analysis, t tests and multiple linear regression analysis. |
| Eales et al. (2021), USA | To examine how the Covid-19 pandemic has affected U.S. families during this time and to examine moderators of the relation between Covid-19 impact and psychological distress of U.S. children and parents. | **Participants**: parents of children aged 2–13 years  **n = 469 parents**  **Age of children**: 5.45 years (SD = 2.41, Range = 2-13). **Age of parents**: M = 38.21 years (SD = 4.45, Range = 25–52). **Gender of parents**: 459 mothers and 10 fathers. **Gender of children**: 239 girls, 228 boys, 1 nonbinary child, and 1 “prefer not to answer” selection.  **Race/ethnicity:** 86.1% of the families, both parent and child were mono-ethnically White and non-Hispanic/Latino. Of the parents, 92.9% were White; 2.1% were Hispanic/Latino; 0.9% were Black/ African American; 3% were Asian; and 3% were multiethnic (which included combinations of White, Black/African American, Asian, and Native American/Alaska Native ethnicities). **Average family income last year**: $125,000–$149,999 (Min ≤ $25,000, Max = $200,000+).  **Educational level of parents**: 52% had received a graduate or professional degree; 6% some graduate school; 33% bachelor’s degree; 8% some college; 1% high school diploma. **Marital status**: 92% married to child’s other parent; 0.4% married, but not to child’s other parent; 4% divorced, 1% separated; 0.3% widowed, 1% single; 1% never married. | Data collection lasted approximately 5 weeks (end of May– early July 2020), right after the state Governor began “opening up” the state and dialing back stay-at-home orders. | **Mixed-method design.**  **Data collected:** 1) Changes Attributable to the Covid-19 Pandemic–Qualitative: Two open-ended questions using text responses: “Please describe anything else you would like to share about the impact of Coronavirus/COVID-19 on your child, whether positive impacts or concerns” and “If you have any final thoughts regarding your child’s media use or how the COVID-19 pandemic has impacted you, your child, or your family, please write them out here.”  2) Changes Attributable to the Covid-19 Pandemic–Quantitative: a. Changes to Work: The adapted version of the Environmental Influences on Child Health Outcomes (ECHO) scale. b. Daily Covid-19 Impact: The Pandemic Stress Index scale. c. Child-Specific Impacts: Items drawn from the CoRonavIruS Health Impact Survey (CRISIS). d. Family-Specific Impacts: An item from the CRISIS scale (changes to the quality of the relationship between the child and other family members). e. Family Coping Strategies were reported by parents on how they and their child were coping with the Covid-19 pandemic by checking all coping strategies from a nine-item list.  3) Potential Moderators of Covid-19 Impact: a. Parental Mediation: An adapted version of TV mediation scale, assessing parental mediation of news and media related to Covid-19. b. Parent Trust in the Federal Government: Parents rated on the statement, “I trust the information I receive about the Coronavirus (COVID-19) from my Federal government” on a 7-point scale. c. Child’s Liking of the United States. Parents reported on a 5-point scale how much they think their child “likes the United States in terms of admiring what this culture/country stands for, enjoying its media (e.g., movies/shows, books, games), using its common language/ slang, or pursuing friendships with others from this background.” 4) Parent and Child Well-Being: a. Parent Mental Health: The Patient Health Questionnaire–4 (PHQ-4). b. Child Psychological Distress: An adapted version of the Child Life Challenges Scale.  **Analytic approach:** Content and thematic analyses for qualitative data and descriptive statistics and regression analysis for quantitative data. |
| Feinberg et al. (2020), Australia | To explore family experiences in response to the early stages of Australia’s Covid- 19 social distancing/isolation restrictions through an inductive approach. | **Participants**: Parents with children under 18 **n = 2,130**  **Age of parents**: M = 38.4 years (SD = 7.1). **Age of children**: M = 8.6 years (SD = 5.2). **Gender of parents**: Cisgender men, n = 398 (19%); Cisgender women, n = 1672 (81%); Transgender or non-binary, n = 1 (0.1%). **Gender of children**: Cisgender boy, n = 1086 (51%); Cisgender girl, n = 1031 (49%); Transgender or non-binary, n = 9 (0.4%). **Aboriginal or Torres Strait Islander:** n = 44 (2%).  **Parent born overseas:** n = 380 (18%). **Geographic location**: Major cities of Australia, n = 1; 267 (60%); Inner Regional Australia, n = 610 (29%); Outer Regional Australia, n = 201 (10%); Remote Australia, n = 38 (2%). **Number of children**: 1 child, n = 613 (29%); 2 children, n = 979 (46%); 3 children, n = 389 (18%); 4 or more children, n = 148 (7%) **Language other than English**: n = 94 (4%). **Low household income** (<$52,000 per year): n = 298 (14%). **Receiving government benefit**: n = 123 (6%). **Single parent household**: n = 239 (11%). **Did not complete high school**: n = 197 (9%). **Highest qualification**: Trade certificate: diploma: or apprenticeship: n = 502 (24%); University: n = 1; 467 (69%). **Unemployment**: One parent unemployed, n = 435 (23%); Two parents unemployed, n = 34 (2%). | Between April 8 and April 28, 2020, when Australians were experiencing social distancing/isolation measures for the first time. | **Qualitative design. Data collected:** 1) Demographic information. 2) An open-ended question asking parents to describe the impact of Covid-19 on their families during the initial stage of lockdown in Australia, which began on March 23, 2020.  **Analytic approach:** Inductive template thematic analysis. |
| Essler et al. (2021), Germany | To investigate temporal dynamics of child wellbeing and problem Behaviour during the early phase of the Covid-19 pandemic and to examine if and to what extent children’s well-being and problem Behaviour change alongside the changes in distal factors (e.g., loosening of public health-related lockdown restrictions from first to second measurement point). | **Participants**: Parents of children aged 3-10 years  **Time 1: n = 2,921; Time 2: n = 890** **Age of children**: 3-10 years (T1). **Gender of children**: Not reported. **Age of parents**: Not reported. **Gender of parents**: Not reported.  **Race/ethnicity of parents or children**: Not reported. **Vocational degree (T1):** 49% university degree, 24% vocational training, 15% university of applied sciences degree, 8% professional academy, 3% Master training, 1% no vocational degree. **Current job status** **(T1):**44% home office, 18% job outside of the home, 17% parental leave, 6% reduced working hours, 5% no job, 4% exempted, 6% other. **Current job status (T2):** 31% home office, 35% job outside of the home, 18% parental leave, 4% reduced working hours, 5% no job, 2% exempted, 5% other. **Change in attendance of educational institutions:** 52% Yes, my child visits preschool again; 34% Yes, my child visits school again; 6% No, my child continues to visit an institution; 4% Yes, my child visits a day-care centre again; 3% No, my child continues to visit no institution due to Covid-19; 1% No, my child continues to visit no institution. **Change in further extra familiar childcare (grandparents, nanny, …):** 40% No, my child continues to receive no extra familial childcare; 34% Yes, my child receives extra familial childcare again; 18% No, my child continues to receive no extra familial childcare due to Covid-19; 7% No, my child continues to receive extra familial childcare. | Time 1: At the peak of the lockdown restrictions in Germany (end of April—beginning of May 2020)  Time 2: After restrictions (e.g., meeting people from other households) had been majorly loosened (in the middle of July 2020). | **Quantitative design (longitudinal).** **Data collected:** 1) Parents’ demographics and children’s demographics at T1 and T2. 2) Parental Strain: Questions assessing parental strain compared to before the pandemic and changes of extra familiar child care situation between T1 and T2 (only T2). 3) Child Well‑Being (T1 and T2): The adapted 52-items KIDSCREEN Health-Related Quality of Life Questionnaire for Children and Adolescents. 4) Child Problem Behaviours (T1 and T2): The adapted Strengths and Difficulties Questionnaire [SDQ]. 5) Parental Self‑efficacy (only T1): The Parenting Self-Efficacy Questionnaire—FSW. 6) Parent–Child Relationship Quality (T1 and T2): The adapted Network of Relationships Inventory [NRI].  **Analytic approach:** Descriptive analysis, correlation analysis, paired-sample t-test, multiple linear regression analysis, cross-lagged panel analyses and True Intraindividual Change models. |
| Ezpeleta et al. (2020), Spain | To describe the life conditions of a sample of adolescents from Barcelona (Spain) during lockdown and identify which factors (e.g., discussions in the family, rule enforcement in the household, relationships with parents) are associated with adolescents’ mental health problems. | **Participants**: Parents **n = 226 Gender of parents**: 191 females, and 35 males. **Age of children (year)**: M = 13.9 (SD = 0.28)  **Gender of children**: 117 females, and 109 males. **Socioeconomic level (SES)**: 30.1% belonged to a high SES, 54.5% to the middle, and 15.4% to low (not clear how the SES was measured). **Race/ ethnicity**: Caucasian (92.9%), American-Hispanic (3.6%), and other ethnicities (3.5%). **Type of family**: Not reported. | Launched on June 9^th^ (lockdown imposed between 13 March and 24 May in Spain). | **Quantitative (longitudinal) design.** **Data collected:** 1) Questionnaire about lockdown (a total of 57 yes/no response format questions referring to lockdown: physical environment, Covid-19 disease, the adults sharing the house, the adolescents’ relationships, the adolescents’ activities, and the adolescents’ feelings/behaviours). 2) Children’s mental health: The Strengths and Difficulties Questionnaire (SDQ; five scales including emotional problems, conduct problems, hyperactivity/inattention, peer relationship problems, and prosocial behaviour).  **Analytic approach:** Descriptive analysis, and paired t-test, and forward stepwise linear regression analysis. |
| Feinberg et al. (2022), USA | To examine change in parent and child mental/Behavioural health and family relationships from before the pandemic to the initial months of the pandemic in a single sample of families with children approximately 8- to 10-year-old, and to  examine factors (e.g., Co-parenting relationship, pre-pandemic levels of parent education or income) that moderate the impact of the pandemic on family member and family well-being. | **Participants**: Parents **n = 206** (122 mothers and 84 fathers from 129 families) **Number of children**: M = 2.3 (SD = 1.0; Range = 1–5). **Age of the oldest child in the family**: M = 9.9 (SD = 1.0) **Gender of the oldest child in the family**: 45.6% female. **Age of parents**: Mothers, M =39.5 years (SD = 4.4); fathers, M = 41.2 years (SD = 4.7). **Race/ethnicity**: 94.7% of parents were non-Hispanic white.  **Educational level of parents**: M = 15.9 years (SD = 1.3). **Living arrangement**: 95% of parents reported they continued to live with the partner with whom they enrolled in the study originally. | April and May 2020 (whether there were any restrictions during this time period is not clear in this study). | **Quantitative design. Data collected:** 1) Depressive symptoms: The Center for Epidemiologic Studies Depression Scale (CES-D Scale). 2) Anxiety: The Penn State Worry Questionnaire. 3) Co-parenting relationship quality over the past month: 14 items drawn from the Co-parenting Relationship Scale. 4) Children’s externalizing and internalizing Behaviour problems during the past month: the Strengths and Difficulties Questionnaire. 5) Parenting quality: Three items from the Parental Behaviour Inventory. 6) Household income and education: Total pre-tax household income in the past year and the “highest grade of school or year of college completed”.  **Analytic approach:** Descriptive analysis, paired t tests and hierarchical linear modeling (HLM). |
| Fioretti et al. (2020), Italy | To investigate Italian adolescents’ subjective experience related to Covid-19 and the national lockdown, and the potential impact of the biographical disruption on developmental tasks (e.g., autonomy acquisition and identity development). | **Participants**: Italian adolescents **n = 2,758**  **Age**: M = 16.64 years (SD = 1.43, Range = 14 - 20). **Gender**: 74.8% females.  **Race/ethnicity:** Not reported. **Type of school the adolescents attended**: lyceums (76.9%), technical high schools (16.9%), and vocational high schools (5.5%).  **Regions of Italy**: 16.8% from Lombardy (the most impacted region), 20.7% from medium impacted regions (Emilia Romagna, Liguria, Marche, Piedmont, Trentino Alto-Adige, Valle d’Aosta, Veneto), and 62.5% from other Italian regions less impacted.  **Personal experiences involving Covid-19**: 7.8% experienced a Covid-19 infection within the family circle (e.g., parents, brothers/sisters, grandparents, etc.), 38.6% experienced Covid-19 infections within friendship, scholastic, or broader social circles (e.g., neighbors, acquaintances), ten participants (0.4%) reported to be infected themselves. | From April 1 to April 5, 2020, during the peak of the Covid-19 outbreak in Italy.  Schools and universities were shut down on March 5. On the March 9, the government declared lockdown status (citizens were required to stay home except for emergencies and primary needs). Over 8 million children and adolescents stopped their social and educational activities, which were reorganized online. | **Qualitative design. Data collected:** Participants were invited to fill in two narrative tasks. The time frame of 2 weeks was referred to time approximately spent between the beginning of lockdown and data collection.  1) The most negative experience: “Please, think about your memories surrounding COVID-19 and the ‘quarantine’. Would you please tell us your most negative experience during the last two weeks? Take your time and narrate what happened and how you experienced it. There are no limits of time and space for your narrative”  2) The most positive experience of life during Covid-19 pandemic: “Referring again to your memories surrounding COVID-19 and the ‘quarantine’, would you please tell us your most positive experience of the last two weeks? Please, narrate what happened and how you experienced that episode. There are no limits of time and space for your narrative”.  **Analytic approach:** Modeling emergent themes analysis using T-Lab Software. |
| Frank et al. (2021), USA | To assess work-family factors and mental health symptoms among physician mothers and fathers during the Covid-19 pandemic using survey data collected from a national longitudinal cohort of early-career US physicians. | **Participants**: Physician parents whose children aged under 18 **n = 215** **Gender**: 114 (53.0%) female and 101 (47%) male. **Age**: weighted mean age was 40.1 years (SD = 3.57).   **Demographics of female participants:** **Age**: 39.6 years (SD = 2.0) **Race/ethnicity**: Not reported. **Relationship status**: Single, n = 1 (0.9%); In a committed relationship, n = 5 (4.4%); Married, n = 99 (86.8%); Separated / Divorced / Widowed, n = 9 (7.9%).  **Number of children**: One child, n = 27 (23.7%); two children, n = 62 (54.4%); three children and more, n = 25 (22.0%). **Pre-Covid-19 employment**: Full-time (≥ 40 h), n = 77 (67.5%); part-time (< 40 h), n = 31 (27.2%); missing, n = 6 (5.3%). **Partner's current employment**: Full-time, n = 83 (72.8%); part-time, n = 10 (8.8%); not employed, n = 21 (18.4%). **Partner's profession**: Physician, n = 39 (34.2%); Non-physician n = 75 (65.8%). **Specialty**: Surgical, n = 14 (12.3%); Non-surgical, n = 99 (86.8%); Missing, n = 1 (0.8%).  **Demographics of male participants: Age**: 39.8 years (SD = 2.2) **Race/ethnicity**: Not reported. **Relationship status of male**: In a committed relationship n = 4 (4%); Engaged, n = 3 (3.0%); Married 92 (91%); Separated / Divorced / Widowed, n = 2 (2.0%).  **Number of children**: One child, n = 17 (16.8%); two children, n = 48 (47.5%); three children and more, n = 36 (35.6%). Pre-Covid-19 employment: Full-time (≥ 40 h), n = 88 (87.1%); part-time (< 40 h), n = 7 (6.9%); missing, n = 6 (6.0%). **Partner's current employmen**t: Full-time, n = 40 (39.6%); part-time, n = 25 (24.8%); not employed, n = 36 (35.6%). **Partner's profession**: Physician, n = 35 (34.7%); Non-physician, n = 66 (65.4%). **Specialty**: Surgical, n = 21 (20.8%); Non-surgical, n = 78 (77.2%); Missing, n = 2 (2.0%). | August 2020 (during the Covid-19 pandemic) but not clear if any restrictions were imposed during this period. | **Quantitative design.** **Data collected:** 1) Demographics and work information including partner’s occupation and employment status. 2) Work and family experiences during the Covid-19 pandemic, including whether participants experienced a loss of childcare or a school closure, whether participants or their partner worked primarily from home, etc. 3) Family-to-work and work-to-family conflict: The Work and Family Conflict Scale. 4) Depressive symptoms: The Patient Health Questionnaire–9 (PHQ-9). 5) Anxiety symptoms: The Generalized Anxiety Disorder 7-item scale (GAD-7).  **Analytic approach:** Descriptive analysis, weighted Chi-square test, weighted t test, and multivariable weighted regression models. |
| Gadermann et al. (2021), Canada | To investigate i) how the Covid-19 pandemic is affecting the mental health of parents and children and what subgroups are most impacted by the pandemic; ii) How parent-child interactions have changed due to the pandemic; and iii) What the factors that support mental health in the family context are. | **Participants:** Parents  **n = 618**  **Age of parents (year)**: M = 43.0 (SD = 9.0, Range =23-58) **Gender of parents**: 324 (52.4%) females, 294 (47.6%) males. **Child age (year)**: 4 and under, n = 183 (29.6%); 5 to 11, n = 292 (47.2%); 12 to 17, n = 309 (50.0%); 18 and over, n = 70 (11.3%). **Ethnicity**: Indigenous origins (e.g., First Nations, Inuit, Métis), n = 17 (2.8%); Visible minority (e.g., Asian, Latin American, Middle Eastern, African), n = 122 (19.7%); European origins (e.g., British, German, Russian), n = 394 (63.8%). **Educational level**: High school or less, n = 62 (10.0%); Some college/university, n = 226 (36.6%); University+, n = 330 (53.4%).  **Employment status**: Unemployed (due to Covid-19), n = 86 (13.9%); Unemployed (prior to Covid-19), n = 21 (3.4%). **Marital status**: Single, never married, n = 39 (6.3%); Married or partnered, n = 517 (83.7%); Separated, divorced, widowed, n = 62 (10.0%).  **Household Living**: Living with a spouse or partner, n = 500 (80.9%); Living with other adult family members (e.g., parents, grandparents), n = 26 (4.2%); Living with grandchildren, n = 11 (1.8%). | Between May 14^th^ to May 29^th^, 2020 (during the first phases of re-opening across many Canadian provinces and territories, emerging from approximately 2 months of lockdown.) | **Quantitative design.** **Data collected:** 1) Questions about participants’ mental health, emotional responses to the pandemic, changes in substance use, experiences of suicidal thoughts, and self-harm. 2) Changes to parent-child interactions, impacts of the pandemic on their children’s mental health, and sources of stress and support for themselves and their children.  **Analytic approach:** Descriptive and bivariate analyses. |
| Gagné et al. (2021), Canada | To investigate changes that have occurred 1 year apart in parenting, child Behaviour, and family violence against children, using data collected before and after the onset of Covid-19. | **Participants**: Parents with at least one child aged 5 -17 **n = 127**  **Gender of parents**: 19.8% fathers.  **Age of the target children**: M = 10 years (SD = 3, Range = 5-17). **Living status of target children**: 79.5% lived with the responding parent full time (79.5%); 15.7% lived with the responding parent between 4 and 6 days per week (15.7%). **Race/ethnicity:** Almost all Canadian citizens (99.2%), the majority identifying with North American (73.2%) or European (14.2%) cultures.  **Location**: 60.6% from Quebec City and 39.4% from Montreal. **Annual family income before the pandemic:** 51.3% between 40,000 $ and 99,000 $; 23.9% earned less and 24.8% earned more.  **Educational level of parents**: 71.0% had a postsecondary degree. **Employment:** 80.0% had a paid job before and 73.6% after the pandemic.  **Working arrangement:** Among parents who were employed or self-employed during the confinement, 38.5% were teleworking, 38.5% commute to their usual work site, 16.5% combine both options, and 6.5% prefer not to answer this question.  **Exposure to the Covid-19 virus:** Participants have had little exposure to the Covid-19 virus. 18 (14.2%) had either been in direct contact with people or material suspected of being contaminated outside their home (n = 16), or lived with a similarly exposed person (n = 1) or a person having had the virus (n = 1). The only respondent who was diagnosed reported moderate symptoms and was not hospitalized. | Wave 1 (pre-Covid-19) data were collected between March 19 and May 22, 2019.  Wave 2 (post-Covid-19) data were collected between May 20 and July 7, 2020.  The Covid-19 containment measures including the closure of schools and day-care services were decreed on March 13, 2020, in Quebec, Canada. However, it is not clear if there were any restrictions during data collection at Wave 2 (post-Covid-19). | **Quantitative design (longitudinal).** **Data collected:** 1) Psychological Distress: The French version of the Kessler Psychological Distress Scale (K10). 2) Parental Self-Efficacy: An in-house translation of the 5-item Parent Self-Agency Measure (PSAM). 3) Parental Stress: The French version of the Parenting Stress Index-4 Short Form (PSI–4-SF). 4) Positive Parenting Practices: The French version of the Positive Parenting Practices subscale of the Alabama Parenting Questionnaire (APQ). 5) Dysfunctional Disciplinary Practices: The French version of the Parenting Scale (PS). 6) The Strengths and Difficulties Questionnaire (SDQ) for children aged 4–16, assessing Emotional Symptoms, Conduct Problems, Hyperactivity/Inattention, and Prosocial Behaviour. 7) Violence Toward the Child: The Psychological Aggression and Minor Physical Violence subscales of the adapted and French version of the Parent–Child Conflict Tactics Scales (PCCTS).  **Analytic approach:** Descriptive analysis, ANOVA and MANOVA, and McNemar’s nonparametric test. |
| Gibbons et al. (2021), Guatemala | To explore Guatemalan mothers’ experiences of parenting during Covid-19, especially their challenges, emotions, and agency during the pandemic. | **Participants**: Mothers who have at least one child under age 7 **n = 12**  **Age of parents**: M = 33.08 years (SD = 4.21, Range = 26–39) **Education** (highest level completed): Junior high school, n = 2; Secondary school, n = 3; Completed law school, n = 1; Licenciatura, n = 3; Master’s degree, n = 3.  **Ethnicity**: Ladina, n = 11; K’iche’, n = 1. **Religion**: Catholic, n = 8; Evangelical Christian, n = 3; Seventh Day Adventist, n = 1. **Marital Status**: Married, n = 9; Separated, n = 2; Single, n = 1.  **Number of children**: one child, n = 5; two children, n = 3; three children, n = 3; four children, n = 1. **Children’s gender:** female, n = 9; male, n = 13. **Children’s age:** three weeks to 11 years.  **Occupation or field of work:** Publicist (n = 2), Vendor of handicrafts, Homemaker, Waitress, Swimming instructor, Domestic worker, Marketing, Clinical psychologist, Cosmetologist, Teacher, Physician. **Working virtually or in-person:** In-person, n = 4; Virtual, n = 2; Hybrid (in-person and virtual), n = 2; Not employed, n = 4. | Between November 2 and December 4, 2020.   All schools were closed in March 2020, public transportation halted, mask-wearing mandated, and a night-time curfew instituted. It is not clear if there were any restrictions in place at the time of data collection. | **Qualitative design.** **Data collected:** Semi-structured interviews (via Zoom or telephone) lasted from 11 to 79 min (Mean of length = 41 min).  **Analytic approach:** Thematic analysis. |
| Goldberg et al. (2020), USA | To examine how a group of heterosexual and same-sex adoptive parents of school-age children are navigating a public health crisis with serious social, economic, and mental health consequences. Specifically, to explore how parents perceive the Covid-19 pandemic and associated home confinement and school closures as affecting the division of labour, their emotional and physical well-being, and their intimate relationship quality. | **Participants**: Adoptive parents with school-age children **n = 89**  **Age of the first adopted child**: M = 12.63 (SD =2.40, Mdn =13.00, Range = 8-21). **Gender of the first adopted child**: 45 (50.6%) girls, 38 (42.7%) boys, and 6 (6.8%) trans. **Race/ethnicity of parents:** 78 (87.6%) White, 2 Latinx, 1 African American, 1 Asian American, and 6 multiracial/biracial.  **Race/ethnicity of the first adopted child:** 34 (38.2%) White, 19 (21.3%) Latinx, 15 (16.9%) biracial/multiracial, 11 (12.4%) Black/African American, 9 (10.1%) Asian/Asian American, and 1 (1.1%) Native American (1, 1.1%).  **Family type**: 32 women in same-sex relationships (referred to as lesbian mothers; LM), 21 men in same-sex relationships (gay fathers; GF), 27 women in different-sex relationships (heterosexual mothers; HM), and eight men in different-sex relationships (heterosexual fathers; HF). **Educational level of parents:** 15 (16.9%) had a PhD, JD, or MD; 34 (38.2%) had a master’s degree; 28 (31.5%) had a bachelor’s degree; 10 (11.2%) had some college or an associate’s degree; and one (1.1%) had a high school diploma/GED. **Parents’ occupation**: 48 (53.9%) in the professional sphere, 23 (25.8%) were managers, 4 (4.5%) technicians/associate professionals, 1 (1.1%) service/sales workers, and 8 (9.0%) were homemakers. **Number of children**: 48 families (53.9%) were parents of only children, 34 (38.2%) had two children, 6 (6.7%) had three children, and 1 family had 5 children. **Family income**: M = $159,596 (SD =$112,661, Mdn =$130 K; Range $21K to $750 K). **Location of residence**: East Coast (34.8%) or West Coast (34.8%) of the United States. | May 1 to June 1, 2020 (not clear if there were any restrictions during this period) | **Mixed-method design.** **Data collected:** A survey of closed- and open-ended questions was used, including: 1) What does the current division of labor (housework, child care, “homeschooling”) between you and your partner look like right now? How is that going for you? What conflicts or issues have come up? 2) How is your relationship with your partner (if relevant)? How has it been affected by the pandemic and resulting changes in your daily schedule, stress levels, etc.? 3) What are your primary worries and concerns right now? 4) How would you describe your mental health since stay-at-home measures have been issued (i.e., since March 2020)? (Improved, Worsened, Stayed the Same.) (This question was repeated for physical health, relationship quality, and sexual intimacy. For each, participants were prompted to “please explain”.  **Analytic approach:** Chi-square analyses for quantitative data and content analysis for qualitative data. |
| Günther‐Bel et al. (2020), Spain | To investigate participants’ lockdown responses in individual, couple, and parental functioning and describe how participants felt their couple and family relationships had changed during the lockdown. | **Participants**: Adults living with a romantic partner and/or children. **n = 407 Age of parents**: M = 42.7 years (SD = 12.7, Range =22-77) **Age of children**: 5 months to 51 years. **Gender of parents**: 77% females. **Race/ethnicity**: Not reported. **Educational level**: 76.7% had university degrees. **Employment status:** 69.9% were at least partly employed; 7.1% had experienced COVID-related job loss; 17.0% were currently unemployed and 5.9% had retired. **Type of family**: Partnered parents living with children (47.4%), partners in couples without children (37.3%), partners in couples whose children were not at home (9.6%), and divorced parents (5.7%). | Between March 24^th^ and April 7^th^, 2020 (during the first 3 weeks of state-regulated lockdown; weeks 2 and 3 of the state-regulated home confinement). | **Mixed-methods design.** **Data collected**: 1) State and trait anxiety: The State-Trait Anxiety Inventory (STAI). 2) Depression: The Beck Depression Inventory (BDI). 3) Dyadic adjustment: The Dyadic Adjustment Scale (DAS). 4) Conjugal, parental, and coparental functioning: The Basic Family Relations Evaluation Questionnaire (CERFB). 5) an open-ended question about perceived changes in couple or family dynamics since the beginning of home confinement.  **Analytic approach:** 1) Descriptive analysis, correlation analysis, and general linear model moderation analysis for the quantitative data. 2) Thematic analysis for the qualitative data. |
| Hamadani et al. (2020), Bangladesh | To evaluate the immediate effects of at least 8 weeks of stay-at-home orders on family economic outcomes and food security, and on women’s mental health and experiences of intimate partner violence, and compare this to their situation before the pandemic. | **Participants:** Mothers whose children had been enrolled in the Benefits and risks of iron interventions in children (BRISC) trial. **n = 2,424 Age of mothers (year)**: M = 24.1 (SD = 4.8) **Age of fathers(year)**: M = 31.1 (SD = 5.9) **Race/ethnicity**: Not reported. **Mothers’ employment status**: Unemployed, n = 2357 (97.3%); Unskilled job, n = 22 (0.9%); Skilled job, n = 43 (1.8%). **Mothers’ educational status**: No education, n = 96 (4.0%); 1–8 years, n = 1242 (51.3%); 9–12 years, n = 996 (41.1%); >12 years, n = 88 (3·6%). **Fathers’ employment status**: Unemployed, n = 24 (1.0%); Unskilled job, n = 451 (18.6%); Skilled job, n = 1830 (75.6%); Other, n = 115 (4.8%). **Fathers’ educational status**: No education, n = 204 (8.4%); 1–8 years, n = 1265 (52.2%); 9–12 years, n = 809 (33.4%); >12 years, n = 144 (5.9%). **Type of family**: Not reported. | Between May 19^th^ and June 18^th^, 2020 (the lockdown started from March 26^th^ to May 30^th^, 2020. After that, the lockdown was reinstated on June 10^th^). | **Quantitative design (an interrupted time-series design)**.  **Data collected**: 1) Awareness and adherence to stay-at-home orders. 2) Food security: The Household Food Insecurity Access Scale (HFIAS). 3) Symptoms of depression: A shortened version of the Centre for Epidemiologic Studies- Depression Scale. 4) Anxiety: The Generalised Anxiety Disorder tool (GAD-7). 5) Experiences of intimate partner violence: The WHO multicounty survey tool.  **Analytic approach:** Descriptive analysis, and regression analysis. |
| Hiraoka and Tomoda (2020), Japan | To quantify parenting stress, and to understand the qualitative structure of parenting stress through textual analysis during this unprecedented situation. | **Participants:** Parents **n = 353 Age of parents (year)**: M = 37.60 (SD = 6.11, Range =23-58) **Age of the youngest child (year)**: M = 6.11 (SD = 4.66, Range =0-18) **Sex of parents**: 273 females, 78 males, and 2 unknown. **Race/ethnicity**: Not reported.  **Type of family**: Not reported. | Between April 29^th^ and 30^th^ 2020 (school closure from March 2^nd^ to the end of April 2020). | **Mixed-methods design.** **Data collected:** Parenting Stress Index – Short Form (PSI-SF) and two open-ended questions.  **Analytic approach:** 1) Descriptive analysis, and T-test were used for quantitative data. 2) Co-concurrence network analysis was used for the qualitative data. |
| Hussong et al. (2022), USA | To assess changes in family functioning as well as associated stressors and youth mental health, using data from an existing longitudinal study with assessments occurring before the pandemic began and during the first months of the pandemic onset (May to July, 2020). | **Participants**: Parents and their children  **n =105 parent-child dyads** (85 families present in both wave 4 and 9, 11 in wave 4 only, and 5 in wave 9 only) **Age of children**: At wave 4, M = 10.6 (SD = 1.17, Range = 8 to 13); at wave 9, M = 13.6 (SD = 1.19, Range = 12 to 16). **Gender of children**: 48% boys. **Gender of parents**: 87% mothers.  **Race/ethnicity of parents**: 80% white, 1% Alaska Native/American Indian, 9% Asian/Asian American, 4% Black/African American, 4% Latinx, and 4% other (summing to over 100% due to individuals identifying with more than one race/ethnicity). **Educational level of parents**: 25% high school graduate without college education, 30% degree from 4-year college, 45% graduate or professional school graduate. | During the first months of the pandemic onset (May 13 to July 1, 2020).  Decisions about Fall 2020 in-person school closures and the move to digital learning that occurred in this area had yet to be announced at the time of this data collection. | **Quantitative design (longitudinal). Data collected**: 1) Demographics. 2) Youth Mental Health (Waves 4 and 9, parent- and child- report): The Pediatric Symptom Checklist. 3) Parent Social Support of Youth (Waves 4 and 9, parent- and child- report): The eight item Arizona Social Support Interview Schedule (ASSIS). 4) Parent’s Marital Satisfaction (Waves 4 and 9, parent-report): The Relationship Assessment Scale. 5) Family Satisfaction (Waves 4 and 9, parent- and child- report): The Family Satisfaction Scale (FSS). 6) Family Open Communication (Waves 4 and 9, parent- and child- report): The Family Communication Scale (FCS). 7) Parent Social Support Scale (Wave 9, parent-report): The abbreviated version of the social support scale from the Support Provision Scale. 8) Child Covid-19-Negative and Positive Life Events (child-report): The Responses to Stress Questionnaire-Covid-19, and 16 researcher developed items. 9) Parent Pandemic Life Events and Distress (Wave 9): The Epidemic-Pandemic Impacts Inventory.  **Analytic approach:** Descriptive analysis, within-person t-tests, and covariate-only path model. |
| Janssen et al. (2020), The Netherlands | To examine the impact of the Covid-19 pandemic on the daily affect and parenting of Dutch parents and adolescents. | **Participants**: Adolescents and parents **n = 101** (adolescents, n = 34; parents: n = 67) **Age (year):** Adolescents, M = 16.95 (SD = 1.01, Range=14.66 -19.01); parents, M = 49.12 (SD = 5.73; Range=36.25 - 71.04) **Gender**: Adolescents, 22 (64.7%) females; parents, 38 (56.7%) females. **Race/ethnicity**: Not reported. **Highest education of parents**: Lower vocational education, n = 2 (3%); Intermediate vocational education, n = 17 (25.4%); Higher vocational education or scientific education (university), n = 48 (71.6%). **Type of family**: Single-parent household, two-parent household, and blended family household (e.g. multiple stepchildren, switching between father and mother). | Before the Covid-19 outbreak (2018–2019) and during the Covid-19 pandemic (April 14^th^ – 28^th^ 2020, during the 5th and 6th week of the lockdown) | **Quantitative (longitudinal) design. Data collected:** 1) Affect: The Positive and Negative Affect Schedule for Children (PANAS-C). 2) Intolerance of uncertainty: The Intolerance of Uncertainty Scale (IUS). 3) Depressive symptoms: The Patient Health Questionnaire (PHQ-9). 4) Some questions about daily parenting, and daily difficulties and helpful activities.  **Analytic approach:** Descriptive analysis and multilevel modelling (using R). |
| Janssens et al. (2021), Belgium | To investigate to what extent the quality of a parent–child relationship is associated with 1) changes in adolescents’ levels of irritability, stress, and loneliness in daily life from before to during the Covid-19 pandemic, and with 2) adolescents’ experiences of Covid-19-related family conflict and its perceived burden. | **Participants**: Adolescents **n = 173**  **Age (Wave 1):** M = 14.2 years (SD = 1.8, Mdn = 14, Range = 11-18) **Age (Wave Covid-19):** M = 16.0 years (SD = 1.9, Mdn = 16, Range = 13-20) **Sex**: 89 females, 84 males. **Family type**: 146 having both a father and a mother in their lives, 2 having two fathers, 2 having only one father or one mother, and 17 indicated ‘Other’ or did not respond. **Race/ethnicity**: Not reported. **Location**: Flanders, the Northern, Dutch-speaking region of Belgium. | Wave 1 (prior-COVID) between January 2018 and June 2019.  Wave 2 (Covid-19): between April 27 and May 10 (during the first national lockdown). | **Quantitative design (longitudinal).**  **Data collected:** 1) Self-report questionnaires: a. Relationship quality (Wave I): A Dutch version of the Inventory of Parent and Peer Attachment (IPPA). Both paternal and maternal relationship qualities was assessed on three dimensions: trust, communication, and alienation. b. Family conflict (Wave Covid-19): Three items from the 22-item Covid-19-related stressors questionnaire adapted from the DynaCORE survey on resilience (https://dynamore-project.eu/). 2) Daily-life data of irritability, stress, and loneliness: Three items were rated on a 7-point Likert scale: ‘I feel irritated’, ‘I feel stressed’, and ‘I feel lonely’.  **Analytic approach:** Descriptive analysis, linear mixed effects models and logistic regressions. |
| Lavenda (2022), Israel | To examine the association between parenting styles, feelings of guilt that these styles might provoke, and the consequences of these feelings on the difficulty of parents to adjust. | **Participants**: Parents of children under 16. **n = 382** **Age**: M = 38.7 (SD = 7.3, Range = 23-57) **Gender**: 78.2% mothers. **Year of education**: M = 16.4 years (SD = 2.3). **Relationship status**: 90% in a committed relationship. **Status of primary caregiver**: 78.4% are primary caregiver. **Average number of children**: 3.1 children per family (SD = 1.6). **Age of the youngest child**: M = 4.9 years (SD = 3.8). **Race/ethnicity**: Not reported. **Type of family**: Not reported. | During the outbreak of Covid-19 (this study did not report the specific date for data collection or any restrictions were existed during that period). | **Quantitative design.** **Data collected:** 1) Demographics and information regarding the extent of exposure to Covid-19-related cases. 2) Parenting Style: The Parent Behaviour Inventory (PBI). 3) Parental Guilt: The Feeling of Guilt about Parenting measure. 4) Adjustment Difficulties: The adapted ultra-brief Adjustment Disorder—New Module 4 (ADNM-4) questionnaire.  **Analytic approach:** Correlation analysis and the PROCESS macro (using for examining a mediation model). |
| Lawson et al. (2020), USA | To evaluate the potential buffering role of parental positive cognitive reframing on the association between parental job loss and psychological maltreatment and physical abuse among parents of 4- to 10-year-olds living in the United States. | **Participants**: Parents **n = 363** (Only 342 parents were included for analysis but the demographics provided are for the sample of 363 parents) **Age of parents**: M = 37.52 years (SD = 6.20) **Age of children**: M = 7.37 years (SD = 2.01, Range = 4 -10) **Gender of parents**: 62% mothers. **Gender of children**: 56.5% male.  **Race/ethnicity of parents**: Caucasian (80.4%), African American (6.3%), Hispanic (5.8%), Asian American (5.0%), and multiracial/other (2.5%). **Annual household income**: 54.4% families had an annual income of $75,000 or higher. Regions of the United States: 18.5% Northeast, 22.9% Southeast, 25.6% Midwest, 20.9% West, and 12.1% Southwest.  **Source of participant recruitment**: Facebook, n = 47; and MTurk, n = 316. | From mid-April to mid-May, 2020, during the peak of unemployment in the United States due to the Covid-19 pandemic (as of August, 2020). | **Quantitative design. Data collected:** 1) Demographics. 2) Experiences with Covid-19 Questionnaire: The economic impact of Covid-19 on their family, including participants’ employment status due to COVID- 19, the time for continuing living at their current address and at their current standard of living without any government support if the Covid-19 pandemic were to last for several months. 3) Psychological Maltreatment and Physical Abuse within the past year and within the past week during the Covid-19 pandemic: The Conflict Tactics Scale Parent-Child version (CTSPC). 4) Cognitive Reframing: The reframing subscale of the Family Crisis Oriented Personal Evaluation Scales (F-COPES). 5) Parental Depression: The Centre for Epidemiologic Studies Depression Scale (CES-D).  **Analytic approach:** Descriptive analysis and logistic regression analysis. |
| Lee et al. (2021), USA | To investigate parent-child activities, at-home educational activities, and parental and child wellbeing six weeks after the World Health Organization (WHO) announced that Covid-19 was a pandemic. | **Participants**: Parents with at least one child 0–12 years of age. **n = 405** **Age of parents**: M = 34.41 years (SD = 7.16). **Gender of parents**: 69% mothers and 31% fathers.  **Race/ethnicity of parents**: White, n = 286 (70.9%); Black, n = 44 (10.9%); Hispanic, n = 42 (10.4%); Other, n = 32 (7.9%). **Employment changed due to Covid-19**: n = 97 (24%). Educational level: High school, n = 57 (14.1%); Some college, n = 180 (44.4%); College plus, n = 168 (41.5%). **Average household income in the prior year**: between $40,000 and $50,000.  **Social distancing days**: M = 26.0 (SD = 10.52). **Lockdown days**: M = 19.58 (SD = 10.67). **Cohabitation status**: Cohabitating (i.e., married; cohabitating with partner; or domestic partnership), n = 323 (80.0%); Not cohabitating (i.e., single, never married; separated; or divorced), n = 82 (20%). **Type of family**: Not reported. | The survey was launched on April 2, 2020, nearly five weeks after the WHO declared that the Coronavirus was a pandemic, and four weeks after the White House issued social distancing guidelines to slow the spread of Covid-19. | **Mixed-method design. Data collected:** 1) Parental perceived preparation to educate at home (3 items, e.g., “I have felt overwhelmed by responsibilities to educate my child at home”). 2) Parental involvement in caregiving: Items adapted from Fragile Families and Wellbeing Study (FFCWS), with two additional items added. 3) Daily schedule disruptions: Parents reported whether their child(ren) had experienced disruptions because of the pandemic (e.g., public school closed, child or daycare closed, etc.). 4) Resources to educate at home: Parents indicate how much they agreed with three statements on a 5-point scale (e.g., “I have support from my child’s school to educate my child at home”). 5) Use of technology for child education: An open-ended question, “What online resources have been the most helpful in educating your child at home?” 6) Child anxiety: The child anxiety subscale of the Child Behaviour Checklist/4–18. 7) Child Behaviour changes in the past 2 weeks during the pandemic: Parents reported whether changes were occurred and if yes, parents described how the child(ren)’s Behaviour changed. 8) Child needs: Parents responded to “What do you think your child(ren) need during this global health crisis?” 9) Parental depression: The Personal Health Questionnaire (PHQ-8). 10) Parental anxiety: The Generalized Anxiety Disorder (GAD-7). 11) Parenting stress: The Aggravation in Parenting Scale.  **Analytic approach:** Descriptive analysis, and multivariate regression analysis for quantitative data, and content analysis for qualitative data. |
| Z. Liu et al. (2021), mainland China | To examine the sleep patterns and sleep disturbances in Chinese preschoolers confined at home, compared to the previous data collected during a normal school term one year prior; to explore behavioural and family associated factors to develop strategies to mitigate negative impacts of Covid-19 on sleep and health in young children. | **Participants:** Caregivers of preschoolers **n = 2,055** (including two sub-samples: 2018 sample and Covid-19 sample) **1) 2018 sample: n = 436** (data collected in late December 2018 during a general school term).  **Relationship with pre-schoolers**: mother, n = 311 (71.3%); father, n = 97 (22.3%); grandparent or other, n = 28 (6.4%) **Gender of caregivers**: females, n = 216 (49.5%); males, n = 220 (50.5%). **Age of children**: 4 years, n = 113 (25.9%); 5 years, n = 204 (46.8%); 6 years, n = 119 (27.3%). **Educational level of caregivers**: Junior high school, n = 136 (31.2%); Senior high school, n = 134 (30.7%); Bachelor’s degree, n = 154 (35.3%); Postgraduate, n = 12 (2.8%). **Race/ethnicity**: Not reported. **Type of family**: Not reported.  **2) Covid-19 sample** (data collected during Covid-19 pandemic)**: n = 1619.** **Relationship with pre-schoolers**: mother, n = 1,264 (78.1%); father, n = 325 (20.1%); grandparent or other, n = 30 (1.8%). **Gender of caregivers**: females, n = 828 (51.1%); males, n = 791 (48.9%). **Age of children**: 4 years, n = 367 (22.7%); 5 years, n = 824 (50.9%); 6 years, n = 428 (26.4%). **Educational level of caregivers**: Junior high school, n = 565 (34.9); Senior high school, n = 494 (30.5%); Bachelor’s degree, n = 537 (33.2%); Postgraduate, n = 23 (1.4%). **Race/ethnicity**: Not reported. **Type of family**: Not reported. | Between February 17^th^ and 19^th^ 2020 (lockdown started from late January 2020, with preschoolers confined to their homes for an extended period). | **Quantitative design.** **Data collected:** 1) Children’s Sleep Habit Questionnaire (CSHQ). 2) Sleep arrangement. 3) The child’s diet, physical activity, and daily electronic device use time. 4) Parenting factors (parenting style, family atmosphere, parent-child communications).  **Analytic approach:** Descriptive analysis, correlation analysis, chi-square test, independent T-test, ANOVA, and regression analysis. |
| J. Liu et al. (2021), mainland China | To examine a complex conceptual model linking daily routines, parent–child conflict, and indices of psychological maladjustment during school closures in the Covid-19 pandemic in a large sample of Chinese children and adolescents. | **Participants**: Children and adolescents, and their mothers **n = 1,594** **Age of children**: M = 13.13 years (SD = 1.54, Range = 9-16) **Gender of children**: 50.6% female, 49.4% male. **Age of mothers**: M = 40.22 years (SD = 4.52) **Race/ethnicity**: Not reported.  **Number of children**: 14% of participants came from single-child households. **Parental education (mother):** 37.0% Middle school or below, 35.9% High school or vocational school degree, 15.9% Vocational college degree, 10.9% Bachelor’s degree, 0.3% Master’s degree or above. **Parental education (father):** 34.3% Middle school or below, 35.4% High school or vocational school degree, 14.9% Vocational college degree, 14.4% Bachelor’s degree, 1.1% Master’s degree or above. **Parental income per month (mother):** 28.7% ¥2,000 or below (approx. $282 or below), 50.6% ¥2,000–¥5,000 (approx. $282–$704), 15.4% ¥5,000–¥10,000 (approx. $704–$1,408), 3.1% ¥10,000–¥15,000 (approx. $1,408–$2,112), 1.3% ¥15,000–¥20,000 (approx. $2,112–$2,816), 0.9% ¥20,000 or above (approx. $2,816 or above). **Parental income per month (father):** 8.0% ¥2,000 or below (approx. $282 or below), 45.7% ¥2,000–¥5,000 (approx. $282–$704), 31.2% ¥5,000–¥10,000 (approx. $704–$1,408), 7.9% ¥10,000–¥15,000 (approx. $1,408–$2,112), 3.6% ¥15,000–¥20,000 (approx. $2,112–$2,816), 3.5% ¥20,000 or above (approx. $2,816 or above). **Location of residence:** Zhengzhou, the capital city of Henan province in China.  **Living place during the Covid-19 pandemic:** 87.6% lived in city, 12.4% lived in rural area. | Data were collected in April of 2020, a period when the Ministry of Education in China still maintained school closure policies and encouraged students to stay at home and study online. | **Quantitative design. Data collected:** 1) Children’s and adolescents’ daily routines during the school closure: The adapted parent-report Child Routine Inventory (CRI). 2) Parent–Child Conflict (child report): The adapted conflict scale of the Parental Environment Questionnaire (PEQ). 3) Loneliness (child report): The Loneliness and Aloneness Scale for Children and Adolescents (LACA). 4) Depressive Symptoms (child report): The Centre for Epidemiologic Studies Depression Scale for Children (CES-DC).  **Analytic approach:** Descriptive analyses, structural equation modeling (SEM, using Mplus) |
| Lucassen et al. (2021), the Netherlands | To investigate whether changes in parental stress from the pre-corona-period (T1) to the height of the first lockdown (T2) were linked to changes in maternal and paternal coercive parenting (e.g., shouting, guilt inducing, spanking) and changes in the Co-parenting relationship (e.g., parental relationship quality, parenting agreement). | **Participants**: Parents  **n = 96 families** (both mothers and fathers participated) **Age of children (T1):** M = 3.44 years (SD = 0.32). **Age of children (T2):** M = 4.72 years (SD = 0.32). **Sex of children (T1):** 51.0% female. **Sex of children (T2):** 53.1% female. **Gender of parents (T1):** 104 mothers and 104 fathers. **Gender of parents (T2):** 94 mothers and 89 fathers. **Family educational background**: 45 families (46.9%) had a lower or medium educational background (neither parents more than intermediate vocational training) and 51 families (53.1%) had a higher educational background (at least one of the parents obtained higher vocational training or [post] university degree). **Race/ethnicity:** Not reported. | Wave 1: Between May 2018 and January 2020, well before the first case of Covid-19 was detected in the Netherlands (February 27, 2020) and the start of the first lockdown in the Netherlands (March 12, 2020).   Wave 2: On April 15, 2020, during the first Covid-19 lockdown (T2) when schools and day-care centres were closed. | **Quantitative design. Data collected:** 1) Parental Stress: The Parental Stress Scale (PSS). 2) Coercive Parenting: The subscale coercive parenting of the Parenting And Family Adjustment Scales (PAFAS). 3) Co-parenting: The subscale Parental Teamwork of the PAFAS.  **Analytic approach:** Descriptive analysis, correlation analysis and Latent Change Score (LCS) modelling. |
| Maggs et al. (2021), USA | To investigate the impact of sociodemographic and alcohol factors (e.g., parents' and adolescents’ drink history) on parents’ permission for adolescents to drink in a family context during the shutdown. | **Participants**: Adolescent siblings and one of their parents **Two adolescent siblings** (n = 911) and **one parent** (n = 456) **Age of older sibling**: M = 15.67 years (SD = 0.68). **Age of younger sibling**: M = 13.14 years (SD = 1.11). **Age gap between the two siblings**: M = 2.2 years. **Age of parents**: M = 45.15 years (SD = 5.37). **Gender of parents**: 85% mothers females, 367 (50.1%) males. **Race/ethnicity of parents**: 85% White, 9% Black, 6% Latino. **Educational level of parents**: 56% college graduate and others not college graduate. **Employment of parents**: 72% currently employed and others not. **Family income**: M =$80,000 - 90,000.  **Type of family**: Not reported. **Alcohol drink status of parents**: 27% abstainers, 49% light drinkers, 24% heavy drinkers. | Wave 1: March 2019 - March 2020. Wave 2: May 1 - June 15, 2020 (during the Covid-19 shutdown with stay-at-home orders). | **Quantitative design (longitudinal).**  **Data collected:** 1) At both waves, parents were asked whether they allowed each child to drink alcoholic beverages at family meals or special occasions. 2) Sociodemographic predictors (W1): Adolescents’ gender and birthdates, and parents’ gender, ethnicity, education, employment, and family income. 3) Alcohol predictors: a. Based on self-reports, parents were coded as abstainers, light drinker, or heavy drinkers. b. Parents rated the perceived risk of drinking alcohol (0 “no risk” to 10 “extremely risky). c. At W1, adolescents reported whether they had ever drunk alcohol, including sips of another person’s drink.  **Analytic approach:** Descriptive analysis and logistic regression analysis. |
| Magson et al. (2021), Australia | To determine the effect of the pandemic and the government-imposed restrictions associated with the response to Covid-19 on the emotional health of adolescents. | **Participants**: Adolescents **n = 248**  **Age of children** (at the time of the Covid-19 survey): M = 14.4 years (SD = 0.5, Range = 13-16). **Gender of children**: n = 126 (51%) girls, n = 122 (49%) boys.  **Race/ethnicity**: 81.8% Caucasian. **Language speaking**: 96.4% speaking English as a first language. **Socioeconomic status**: 79.2% middle to high socioeconomic status. **Type of family**: Not reported. | Time 1 (prior to the Covid-19 pandemic): Data were collected online throughout 2019 as part of the larger Risks to Adolescent Wellbeing Project.  Time 2 (during the pandemic): Between May 5 and May 14 2020, approximately two months after the Australian government had imposed the stay-at-home orders and schools had moved to online learning. | **Quantitative design (longitudinal). Data collected:** 1) Anxiety: The Generalized Anxiety subscale of the Spence Children’s Anxiety Scale (SCAS-C). 2) Depressive symptoms: The Short Mood and Feelings Questionnaire—Child Version (SMFQ-C). 3) Life satisfaction: The Student’s Life Satisfaction Scale (SLSS). 4) Covid-19 related distress: Adolescents indicated how distressed they were about each item listed on a 10-point scale (researcher developed measure, 18 items). 5) Disruption to schooling: Four items were developed to assess: format of school attendance, difficulties during online learning, motivation to complete school work, and impact on education. 6) Media exposure: Two questions assessed students’ exposure to traditional news media and social media related exposure. 7) Interpersonal conflict: Four items were developed to assess change in interpersonal conflict between adolescents and their mothers, fathers, siblings, and friends due to the Covid-19 social distancing rules and stay at home restrictions. 8) Social connectedness of students: The Social Connectedness Scale (SCS). 9) Adherence to Covid-19 Australian government stay-at-home directive: Adolescents indicated how often they had left their home for reasons other than those listed, responding on a 5-point scale.  **Analytic approach:** Descriptive analysis, correlation analysis, paired samples t-test, and moderation analysis. |
| McRae et al. (2021), New Zealand | To examine the impact of parental distress on poor parenting during Covid-19 lockdowns and whether partner support and cooperative Co-parenting can buffer this effect. | **Participants**: Parents **n = 362** (310 were from the same family, i.e., 155 dyads) **Age of children** (T1, pre-pandemic): 4-5 years. **Age of children** (T2, during the lockdown): 5-7 years. **Number of children**: M = 2.26 (SD = 0.87). **Age of parents**: M = 37.03 years (SD = 4.84). **Gender of parents**: 199 mothers, 163 fathers. **Ethnicity of parents:** 55% New Zealand European/Pakeha, 12.7% Non-New Zealand European, 10.8% Asian , 5.8% Maori, 5.5% Pacific nations, 5.0% Indian, and 4.1% Other ethnicity not listed.  **Relationship length**: M = 11.81 years (SD = 4.54). **Marital status**: 86% married, 14% cohabiting. **Educational level**: 32.3% postgraduate (e.g., postgraduate diploma, honors degree, master’s degree), 49.2% Tertiary (e.g., college/ university degree, technical qualification, trade certificate), 14.6% High school certificate. **Personal income prior to the lockdown** (per annum New Zealand dollars): 29% less than 40,000, 14% 41,000-60,000, 16% 61,000-80,000, 16% 81,000-100,000, and 26% more than 100,000. **Personal income during the lockdown** (per annum New Zealand dollars): 38% less than 40,000, 12% 41,000-60,000, 15% 61,000-80,000, 15% 81,000-100,000, and 20% more than 100,000. **Work/income loss during lockdown**: 31% lost work, 34% lost income. | Time 1: Pre-pandemic  Time 2: March 26 – April 28, 2020, during the strict Level-4 lockdown. During this time, families were legally required to stay within their immediate household with no physical contact outside the home allowed, except when one person needed to gather essential resources (groceries, medicine) under strict social distancing protocols. | **Quantitative design (longitudinal). Data collected:**  1) Psychological Distress (T1 & T2): a. Emotional Well-Being: Five items from the RAND 36-Item Health Survey. b. Depressive Symptoms: The Centre of Epidemiological Studies Depression Scale. c. Stress: The Perceived Stress Scale.  2) Parenting (T1 & T2): a. Harsh and Warm/Responsive Parenting: The Parenting Styles and Dimensions Questionnaire (short version). b. Parent–Child Relationship Quality: 10 items adapted from prior measures assessing facets of parent–child relationship quality.  3) Partner Support and Cooperative Co-parenting (T2): a. Perceived Partner Support: Seven items capturing how often partners had been supportive and provided emotional, esteem, and practical support during lockdown. b. Cooperative Co-parenting: Five items capturing cooperative Behaviour, Co-parenting agreement, and competitive Co-parenting during lockdown.  **Analytic approach:** Descriptive analysis, correlation analysis and regression analysis. |
| Marchetti et al. (2020), Italy | To investigate the effect of parents’ psychological distress and verbal aggression on Behavioural and emotional symptoms of children during the Covid-19 lockdown. | **Participants**: Parents **n = 878 Age of parents**: M = 40.58 years (SD = 6.41, Range = 23-67).  **Age of children**: M = 7.54 years (SD = 3.16, Range = 3-13). Gender of parents: Mothers, n = 767 (87.4%); fathers, n = 111 (12.6%)  **Marital status**: Single, n = 32 (3.6%); married, n = 643 (73.2%); living with a partner, n = 127 (14.5%); separated/divorced, n = 72 (8.2%); widowed, n = 4 (0.5%) **Gender of children**: Male, n = 451 (51.7%); female, n = 427 (48.3%). **Race/ethnicity**: Not reported. **Employment status**: Employed, n = 738 (84.1); Unemployed, n = 140 (15.9%). **Educational level**: Less than high school, n = 55 (6.2%); High school, n = 343 (39.1%); More than high school, n = 480 (54.6%). **Geographic area**: North, n = 222 (25.3%); Centre, n = 277 (31.5%); South, n = 379 (43.2%). | In the first weeks (from April 3 to 14, 2020) of the lockdown in Italy. | **Quantitative design. Data collected**: 1) Sociodemographic information. 2) Psychological distress of parents: The General Health Questionnaire-12. 3) Verbal hostility of parents: Three items of the Italian short version of the Parenting Styles and Dimensions Questionnaire. 4) Emotional symptoms and hyperactivity/inattention of children (assessed by parents): The two subscales of the Italian version of the Strengths and Difficulties Questionnaire-Parent Report (SDQ).  **Analytic approach:** Descriptive analysis, Pearson correlation analysis, and PROCESS Model 6 in SPSS 26.0. |
| Montirosso et al. (2021), Italy | To investigate psychosocial responses in a large cohort of children/adolescents with neurodevelopmental disabilities and in their parents during the Covid-19 outbreak in Italy. | **Participants**: Parents who have children/adolescents with neurodevelopmental disabilities (NDD)  **n = 1,472 parents of 1632 NDD children/adolescents** **Age of children**: 609 (37.4%) 0-6 years, 564 (34.6%) 7-10 years, 457 (28.0%) 11-18 years. **Age of parents**: M = 42.48 years (SD = 6.5, Range: 23-71) **Gender of parents**: 83.1% mothers. **Gender of children**: 550 (33.5%) female, 1,082 (66.3%) male. **Marital status**: 1,253 (85.1%) married/ cohabitant, 132 (9.0%) divorced/separated, 77 (5.2%) single, and 6 (0.4%) widowed. **Diagnosis of children**: 1,068 (65.4%) single, 564 (34.6%) multiple. **Race/ethnicity of children**: Not reported. **Race/ethnicity of parents**: Not reported. | Between April 8th and 20th, 2020, after about one month of lockdown and restriction measures due to the Covid-19 pandemic in Italy. | **Quantitative design.**  **Data collected:** An ad-hoc questionnaire was developed for this study, including new specific items and items taken by validated measures. 1) Demographics. 2) Covid-19 outbreak and restriction-related variables included parents’ perception and information about: a) the impact of containment on family routines and daily life; b) if either a parent or a close relative/friend/ colleague had tested positive for Covid-19; c) whether the family had experienced serious consequences or bereavements due to Covid-19. 3) Behavioural regulation problems in children/adolescents: The parent-report Child Behaviour Checklist for the evaluation of peri- and posttraumatic Behavioural response in developmental age (CBCL-PTSD). 4) Parental stress: The Parental Stress subscale from the Parental Stress and Coping Inventory (PSCI). 5) Parental Resilience: The Resilience Scale for Adult (RSA).  **Analytic approach:** Descriptive analysis and hierarchical stepwise regression analysis. |
| Neubauer et al. (2021), Germany | To examine change in parental well-being, perceived family environment, and parent-rated child Behaviour across 3 weeks during a time of school closings and other counter-Corona measures, and to examine daily experiences as potential mechanisms driving change in these adjustment measures. | **Participants**: Parents of school children  **n = 469**  **Age of parents**: M = 42.93 years (SD = 6.40). **Age of target children**: M = 9.81 years (SD = 2.85, Range = 6–19). **Gender of children**: 500 male, 460 female, 3 diverse/nonbinary.  **Race/ethnicity**: Not reported. **School type that children attended:** elementary school, n = 610 (62.9%); the academic tier of secondary school (German Gymnasium), n = 240 (24.7%); basic or vocational secondary school, n = 20 (2.1%); comprehensive school, n = 53 (5.5%); other school types, n = 44 (4.5%). | Baseline online survey: from March 27 to April 3, 2020.  Longitudinal data collection: Across 3 weeks during a time of school closings and other counter-Corona measures (end of March until end of April 2020). | **Quantitative design (longitudinal). Data collected**: 1) Baseline and Post Measures: a. Behaviours of the youngest school-aged child in the household: Three subscales of the Strengths and Difficulties Questionnaire (SDQ). b. Family environment: A German adaptation of the Family Environment Scale. c. Parents’ stress: The 10-item version of the Perceived Stress Scale. d. Vitality: The Subjective Vitality Scale.  2) Daily Measures: a. Autonomy-supportive parenting: Two items adapted from similar assessment approaches in previous research. b. Child well-being: Eight items, including negative affect and positive affect. c. Parental need fulfillment: The daily diary version of the revised Balanced Measure of Psychological Needs Scale.  **Analytic approach:** Descriptive analysis, exploratory and confirmatory factor analyses, dynamic structural equation models, and latent change regression models. |
| Nyanamba et al. (2022), USA | Use both variable- and person-centred approaches, to assess parents’ motivations for involvement in remote learning, the influence of need frustration indicators (burnout, academic concern) on motivation, and the consequences of motivation styles and profiles on need-supportive Behaviour. | **Participants**: Parents with 5- to 8-year-olds **n = 218** **Age of children**: M = 6.5 years (SD = .96, Range = 5-8), **Gender of children**: 81 (37%) females, 112 (51%) males. **Age of parents**: 6 (3%) 18-25 years, 150 (68%) 26-35 years, and 42 (19%) 36-45 years. **Gender of parents**: 114 fathers, 83 mothers, and 21 preferred not to say.  **Parent ethnicity:** 152 (70%) White/Caucasian, 25 (11%) African American, 17 (8%) Native American/Alaskan Native/Hawaiian, 4 (2%) Asian, and 9% did not specify. **Family SES**: 77% of the families qualified for and received free meals from their schools before COVID and during the shelter-in-place period. **Online learning support**: 75% of the families received online learning support from their children’s schools. **Type of family**: Not reported. | This study did not report the time of data collection but the research recruited participants purposely from states in the USA with the most prolonged first shelter-in-place restrictions (by early June 2020) during the Covid-19 pandemic. | **Quantitative design.**  **Data collected:** 1) Demographic information. 2) Parents’ Concern for Their Children’s Academic Lag was rated by an item on a scale of 0–100, “With the Covid-19 pandemic disruption, to what extent are you concerned about your child falling behind in their academics?” 3) Parents’ Motivation for Involvement in Remote Learning: The adapted Parents’ Motivation for Help in Homework scale, which evaluated autonomous and controlled motivations. 4) Quality of Parental Involvement in Remote Learning: The adapted Parent Need-Supportive Behaviours scale. 5) Parental Burnout: The Parental Burnout Assessment (PBA).  **Analytic approach:** Descriptive analysis, correlation analysis, independent samples t-test, multiple regression analysis,  latent profile analysis, and logistic regression analysis. |
| Overall et al. (2021), New Zealand | To investigate whether sexist attitudes assessed prior to the pandemic predicted residual changes in aggressive Behaviour toward intimate partners and children during a nationwide lockdown. | **Participants**: Parents **n = 362** (of which 310 were drawn from the same family) **Age of parents**: M = 37.03 years (SD = 4.84) **Length of relationship**: M = 11.88 years (SD = 4.53) **Marital status**: 87 % married, and 13% cohabiting but not married. **Number of children**: M = 2 (SD = .87).  **Ethnicity**: Ma¯ori (5.8%), Pacific Nations (5.5%), Indian (5%), Asian (10.2%), non-NZ European (12.4%), NZ European/Pa¯keha¯ (56.4%) and Other (4.1%). **Families income prior to the pandemic** (per annum NZD): 27% <40,000, 31% 41,000–60,000, 17% 61,000–80,000, 17% 81,000–100,000, and 26% >100,000. **Families income during the pandemic** (per annum NZD): 38% <40,000, 12% 41,000–60,000, 15% 61,000–80,000, 16% 81,000–100,000, 20% > 100,000. **Work loss status during the pandemic**: 112 (31%) lost work and/or 122 (34%) lost income due to Covid-19, | Time 1: Prior to the Covid-19 pandemic.  Time 2: Between April 8 and 27, 2020, during a nationwide Covid-19 lockdown in which families were legally required to stay within their immediate household with no physical contact outside the home allowed. All schools and businesses were closed, except for essential services (e.g., health services, grocery stores). Occasionally one person could leave the house to gather essential resources (e.g., medicine, groceries) under strict distancing protocols. | **Quantitative design (longitudinal).**  **Data collected:** 1) Sexist Attitudes Assessed Prior to Covid-19 Lockdown: The Ambivalent Sexism Inventory.  2) Aggressive Behaviour Assessed Prior to the Pandemic and During the Lockdown: a. Aggressive Behaviour Toward Intimate Partners: The verbal aggression subscale from previous research. b. Aggressive Parenting: Four items from the Parenting Styles and Dimensions Questionnaire.  3) Power, Relationship Quality, and Stress Assessed During Lockdown: a. Power During Couple Interactions: Four items regarding how parents felt when interacting with their partner during the lockdown, including feeling powerful and assured and powerless and ineffective. b. Power During Parent–Child Interactions: The same four items regarding how parents felt when interacting with their child during the lockdown. c. Couple Relationship Quality: The Investment Model Scale. d. Parent–Child Relationship Quality: The Investment Model Scale. e. Stress: 12 items about how stressful different areas of life had been during lockdown.  **Analytic approach:** Descriptive analysis, t test, and the linear mixed-effects models (MIXED) procedure in SPSS. |
| Peltz et al. (2021), USA | To examine a mediation model of links between Covid-19–related stressors, co-parental conflict, and family cohesion within parents/caregivers recruited within the first five weeks of the pandemic breaking in the United States. | **Participants**: Parents/caregivers of children aged 5-18 **n = 1,003** **Age of parents**: 40.9 years (SD = 8.5). **Gender of parents**: 72.4% female. **Marital status of parents**: 86.1% married, engaged, or in committed relationships.  **Race/ethnicity** **of parents**: 82.1% Caucasian, 5.7% African American, 4.9% Latinx or Hispanic, 2.2% Asian or Pacific Islander, and 3.4% unknown or “other.” **Length of relationship**: M = 14.5 years (SD = 17.2). Educational level of parents: 40.0% holding a graduate degree (i.e., M.A./M.S., J.D., Ph.D., D. D.S., or M.D.), 33.1% holding a bachelor's degree, 22.7% having some education or training after high school (i.e., some college, associate’s degrees, or trade school training/certificates), and 4.3% having a high school education or less. **Employment status prior to and during pandemic**: 83.4% reported working prior to the pandemic (67.2% full time, 16.2% part time). During the pandemic, 69.8% reported working at least part-time from home in the week prior to entering the study and 24.3% reported working at least part-time away from home. **Having a co-parent** **or not**: Roughly 80% respondents reported having a co-parent helping to raise their child(ren).  **Co-parenting partner**: 93.6% romantic partner, 4.8% one of the respondent’s parents (i.e., child’s grandparent), 1.6% another individual (e.g., child’s uncle/aunt/cousin). **Number of children**: M = 1.8 (SD = 1.0). 44.5% having one child living in the home, 37.3% having two children, 12.0% having three children, and 6.3% having four or more children in the home.  **Age of children**: M = 9.8 year (SD = 4.4).  **Biological child or not**: 88.2% had at least one of the children in the home was their biological child. **Location of residence**: 29.4% residing in the Northeast, 21.8% from the Midwest, 30.2% from the South, and 18.7% from the West of the United States. Approximately 2% of the participants reported residing outside of the United States (e.g., Canada, England, or New Zealand). **Household income**: M = $83,631 (SD = $36,334), with 22.2% of parents reporting family incomes of $50,000 or less. | From the end of March to the end of April of 2020 (within the first five weeks of the pandemic breaking in the United States).  All 50 states had ordered some form of school closures by March 16th, with the majority of states closing schools for the remainder of the school year. Although California was the first state to issue lockdown and stay-at-home orders on March 19, by March 28 more than half of the states had issued similar orders, and all but four states had issued some form of stay-at-home orders by April 7. | **Quantitative design (longitudinal).** **Data collected**: Online weekly diary surveys for five weeks. 1) Baseline Stress of New Work/Parenting Demands (researcher developed items). 2) Weekly Stress Over Finances within baseline survey and the weekly diaries (researcher developed items). 3) Weekly Health-Related Stress within baseline survey and the weekly diaries (researcher developed items). 4) Weekly Co-parenting Conflict: Two items from the co-parental interaction scale within the baseline survey and the weekly diaries. 5) Weekly Family Cohesion within baseline survey and the weekly diaries: Two items from the Family Assessment Device.  **Analytic approach:** Descriptive analysis, association analysis, ANOVA and chi-squared analyses, and multilevel SEM models. |
| Penner et al. (2021), USA | To investigate the impact of Covid-19 in a youth subsample one month after stay-at-home measures were put in place; to examine changes in mental health from before the pandemic to during the outbreak, and to examine whether job loss or family functioning influenced change in youth mental health in the Covid-19 survey subsample. | **Participants**: Adolescents aged 10-14 years old **n = 185** [This is a Covid-19 survey subsample of a longitudinal study called Brief Problem Monitor (BMP)] **Age (year)**: M = 12.21 (SD = 1.31) **Gender**: 56.2% female, 42.7% male. **Grade**: 28.1% in 5^th^ grade, 4.9% in 6^th^ grade, 34.6% in 7^th^ grade, and 32.4% in 8^th^ grade. **Race/ethnicity** 71.9% Hispanic/Latinx, 9.7% Black or African American, 5.4% Multiple Races, 1.1% Asian, 1.6% (5) White, 1.1% American Indian, 5.4% multiple races, and 5.4% (14) self-identified their ethnicity or race as “other.” **Type of family**: not reported. | 1) Baseline survey: January 2020, pre-pandemic;  2) Covid-19 survey: at three bi-weekly follow-up points in April-May 2020 (school closure from March 13, 2020, to the end of the school year). | **Quantitative design.** **Data collected**: 1) Effects of Covid-19 Pandemic at Home over the previous two weeks (adapted from a recent Covid-19 survey): physical contact with other people, family finances and access to food, family loss of work, media exposure, loneliness, contact with friends, parent and child stress, family functioning. 2) Youth Mental Health: The Brief Problem Monitor (BPM), including four scales (internalizing problems, attention problems, externalizing problems, and total problems).  **Analytic approach:** Descriptive analysis, correlation analysis, and repeated measures mixed ANCOVA. |
| Polack et al. (2021), USA | To investigate changes in interpersonal dynamics and mental health during Covid-19, and to examine whether the associations between different social contexts changed during Covid-19 and whether changes in social interactions during Covid-19 was associated with changes in depressive symptoms. | **Participants**: Youth aged 8-15 years **n = 112** **Age**: 11.77 years (SD = 2.13, Mdn = 12) in Wave 1, and 12.64 years (SD = 2.12, Mdn = 13) in Wave 2. **Gender**: 62 females, 50 males. **Race/ethnicity**: Caucasian (73%). **School activities** at Wave 2: 97.3% reported school moving to a full or partial online format (104 participants reported attending school completely online and four reported partially online). 91.9% reported that afterschool activities were partially or fully cancelled (93 reported all activities cancelled, nine reported some were cancelled).  **Living status at Wave 2**: 107 (95.5%) reported living with three or more people in their household. 105 (94.06%) reported that at least one parent stayed home (70 participants reported both parents at home, 35 reported at least one parent). | Wave 1 (prior to Covid-19 pandemic): Between January 31, 2019 and September 23, 2019.  Wave 2 (during to Covid-19 pandemic): Between March 30, 2020 and June 8, 2020.  Public schools were closed on March 13, 2020, public amusement places on March 18, 2020, and nonessential businesses on March 23, 2020. “Stay at home” order was issued on March 28, 2020. Connecticut started phase 1 of reopening May 20, 2020; Phase 2 started after the end of data collection. | **Quantitative design (longitudinal).** **Data collected:** 1) Background Covid-19-Related Questions (Wave 2): If school had been moved to an online format, if youth’s usual activities had been canceled, etc. 2) Diary Measures: a. Interpersonal Interactions (Waves 1 and 2): Every day, participants were asked to report whether or not a list of interpersonal interactions occurred, and if they did, with whom. Number of events were summed every day within social context. b. Depressive Symptoms (Waves 1 and 2): The self-report Children’s Depression Inventory–Short version (CDI-S).  **Analytic approach:** Descriptive analysis, correlation analysis, ANOVA, and hierarchical regression analysis. |
| Qu et al. (2021), mainland China | To examine the role of parent–child conflict and intimacy in adolescents’ psychological distress during the transition of school reopening in Covid-19 through a two-wave longitudinal design with Chinese adolescents. | **Participants**: Adolescents **n = 879** **Age**: M = 13.14 years (SD = 1.31, Range = 11-16). **Gender**: 51% females.  **Race/ethnicity**: In the area where the students’ schools located, almost all of the population (99%) is of Han descent. **Location of residence**: Shanghai. **Type of family**: Not reported. | Wave 1: April 24, 2020, immediately before school opening.  Wave 2: July 6, 2020, three months after school opening during Covid-19. | **Quantitative design (longitudinal). Data collected:** 1) Parent–Child Conflict (wave 1, child-report): The conflict subscale from the Network of Relationships Inventory (NRI) at the first wave. 2) Parent–Child Intimacy (wave 1, child-report): The intimacy subscale from the Network of Relationships Inventory (NRI) at the first wave. 3) Adolescents’ Perceived Stress Toward School Reopening (wave 1, child-report): Adolescents’ feeling about school reopening (excited, neutral, or stressed). 4) Adolescents’ Depression (wave 1 & 2, child-report): The “LEVEL 2—Depression—Child Age 11–17” scale from the Diagnostic and Statistical Manual of Mental Disorders (DSM–5). 5) Adolescents’ Anger Problems (wave 1 & 2, child-report): The “LEVEL 2—Anger—Child Age 11–17” scale from DSM–5.  **Analytic approach:** Descriptive analysis, association analysis, and hierarchical regression analyses. |
| Ren et al. (2021), mainland China | To examine how Covid-19 related stressors (i.e., presence of confirmed or suspected cases in parents’ communities and frequency of consuming virus-related information) may contribute to young children’s engagement in disease prevention practices and mental health via parenting (i.e., fear induction practices). | **Participants**: Parents with elementary-school-age children. **n = 240** **Age of parents**: M = 38.50 years (SD = 5.80) **Gender of parents**: 75% mothers **Age of children**: M = 9.48 years (SD = 1.39, Range = 7-12) **Gender of children**: 46% females. **Ethnicity of parents:** Han ethnicity.  **Marital status of parents**: 97% married  **Socioeconomic status**: 81% low- to middle-socioeconomic status with a monthly household income below RMB 8,000 (around USD$1,300).  **The** **highest education level of parents**: 12% elementary school or below, 38% middle school, 42% high school or equivalent, and 8% college or above. **Exposure to Covid-19 or not:** None reported that their family members, close friends, or themselves had Covid-19 or were suspected of having Covid-19. **Location of residence**: Wenzhou, China. | Time 1: From January 28 to 30, 2020 (during which Wenzhou witnessed a steep rise of confirmed Covid-19 cases but the quarantine policy had not yet been implemented).  Time 2: Between March 7–11, 2020 (during which there were no new confirmed cases or deaths reported and the official quarantine policy had ended in Wenzhou). | **Quantitative design (longitudinal). Data collected:** 1) Covid-19 Cases in the Community (T1). 2) Parents’ Information Consumption (T1): How often parents consumed Covid-19-related information from four different sources during the past few days at T1. 3) Parents’ Fear Induction Practices (T1, researcher developed items). 4) Children’s Participation in Covid-19 Prevention Practices (T1, researcher developed items). 5) Children’s Trait Anxiety (T1): The 20-item trait subscale of the State–Trait Anxiety Inventory for Children (STAIC). 6) Children’s Depressive Symptoms (T2): The Centre for Epidemiologic Studies Depression Scale for Children (CES-DC).  **Analytic approach:** Descriptive analysis, association analysis, and the Bayesian moderated mediation model (Mplus 8). |
| Rodriguez et al. (2021), USA | Study 1: to determine whether parents’ economic concerns, worries, and loneliness were significantly associated with perceived increases in adverse parenting during the pandemic.  Study 2: to examine whether mothers’ perceptions of their pandemic-related parenting were associated with their current child abuse risk; explore factors associated with adverse changes in parent-child interaction and increased abuse risk; evaluate whether child abuse risk during the pandemic increased from pre-pandemic levels, controlling for pre-pandemic income; examine whether parents’ perceptions of pandemic-related increases in parent-child conflict significantly related to abuse risk during the pandemic controlling for their pre-pandemic abuse risk or pre-pandemic income. | This paper presented two separate studies with different samples.  **1. Study 1**  **Participants**: Parents of at least one child aged 12 or younger  **n = 405**  **Age of parents (year):** M = 37 (SD = 7.17)  **Gender of parents**: 69% females, 731% males.  **Race**: White (71%), Black (11%), Hispanic (10%), and Other (8%).  **Employment status**: Not reported.  **Educational level**: A bachelor’s degree or higher (42%).  **Cohabitants**: 80% live with a partner.  **Type of family:** Not reported  **2. Study 2**  **Participants**: Mothers enrolled in a prospective longitudinal study in the Southeast U.S., the “Following First Families (Triple-F)” Study.  **n = 106**  **Age of parents (year):** M = 37 (SD = 7.17)  **Age of children (year):** 5 - 6.5.  **Gender of parents**: all females.  **Race**: White (60.4%), African-American (36.8%), Asian (1.9%), and Native American (0.9%), Hispanic/Latina (4.7%), and biracial (5.7%).  **Employment status**: Not reported.  **Educational level**: 15.7% high school; 22.5% some college; 28.4% college degree; 33.3% > college degree.  **Cohabitants**: 83% were living with a partner or spouse.  **Type of family:** Not reported. | **Study 1:** From April 14^th^ to 17^th^, 2020 (about 4 weeks after the White House provided social distancing guidelines for the U.S)  .  **Study 2**: The study used data of the Triple-F Study, which were collected at time 4 (when the focal child was between 4 and 4.5 years old) and time 5 (during the early phase of the pandemic, April 20–May 31, 2020; the time when the focal child was between 5 and 6.5 years old). | **Quantitative design.**  **Study 1**  **Data collected:** 1) Pandemic-related parenting stress and loneliness using statements with a 4-point Likert scale. 2) Pandemic-related perceived changes in parenting: Seven questions measuring conflict, verbal aggression, and neglect). 3) Personal Covid-19 experience: Two questions about Coronavirus infection and social isolation.  **Study 2**  **Data collected: 1)** The Parent-Child Conflict Tactics Scale (CTSPC, measured at time 4 & 5). 2) Child abuse risk: The Adult-Adolescent Parenting Inventory-2 (AAPI-2; measured at time 4 & 5). 3) The Brief Child Abuse Potential Inventory (measured at time 5). 4) Covid-19 pandemic-related perceived changes in parenting (consistent with study 1, measured at Time 5). 5) The financial impact of the pandemic (employment loss and food insecurity).  **Analytic approach:** Descriptive analysis, T-tests, Chi-square analysis, correlation analysis, and regression analyses. |
| Romero et al. (2020), Spain | To examine the effects of the extreme Spanish lockdown in children’s behavioural and emotional adjustment, and the effects of parent-related variables on children’s behaviours. | **Participants:** Caregivers of children aged 3 to 12 years **n = 1,049 Relationship to children**: 940 (89.6%) mothers, 102 (9.7%) fathers and 7 (0.7%) different caregivers. **Age of children (year):** M = 7.29 (SD = 2.39, Range =3-12)  **Gender of children**: 50.4% females. **Race/ethnicity**: Not reported. **Type of family**: Not reported. | From April 8^th^ to 27^th^, 2020 (during the most restrictive period of the lockdown). | **Quantitative design.** **Data collected:** 1) Parent-Related Variables: a. Resilience: The Connor–Davidson Resilience Scale (CD-RISC-10); b. Perceived distress (developed ad hoc for the current study); c. Emotional problems: The Patient Health Questionnaire for Depression and Anxiety (PHQ-4); d. Parenting distress: A newly developed measure for the confinement situation; e. Specific parenting including focused parenting, a soothing attitude by parents, structured parenting, and avoidant parenting behaviours.  2) Child Outcomes: a. Child negative outcomes: 14 items from the parent-reported Strengths and Difficulties Questionnaire (SDQ), addressing conduct problems, emotional problems, and hyperactive behaviours; b. Child positive outcomes (developed ad hoc for the current study) including child adaptation to daily routines, the involvement in prosocial activities, socially-oriented conceptions and attitudes, and the willingness to keep in contact with significant others.  **Analytic approach:** Descriptive analysis, exploratory factor analysis (EFA), correlation analysis, chi-square tests, and path analysis. |
| Rudolph and Zacher (2021), Germany | To model changes in family demands and satisfaction with family life that occurred during the Covid-19 pandemic in Germany and to model how two central aspects of one’s family ecology—partnership status and parental status—modify changes in family demands and satisfaction with family life over time. | **Participants:** Parents with children under 18 **n = 1,042**  No demographics were reported. | Between early April 2020 and early September 2020.  The baseline survey conducted in December of 2019 (T0). Data were collected monthly during the first week of April (T1), May, July, August, and September (T5) of 2020. | **Quantitative design (longitudinal).**  **Data collected:** 1) Demographics. 2) Family Demands: Three items based on a similar measure of perceived demands in the domain of paid employment. 3) Satisfaction with Family Life: A single item, “All in all, how satisfied were you with your family life in the last 4 weeks?” 4) Partnership Status: A single item was used, “Do you live in a stable partnership?” 5) Parental Status: How many children participants have and their ages. 6) Job Demands: reflecting on the past 4 weeks of their lives and responding to: “How often does your job require you to work very fast?”, “How often does your job require you to work very hard?”, and “How often is there a great deal to be done in your job?”  **Analytic approach:** Descriptive analysis, correlation analysis, and unconditional and conditional multivariate latent growth curve modelling. |
| Russell et al. (2020), USA | To examine the potential linkages between mental health indicators and parents’ report of child-parent relationship outcomes. | **Participants**: Caregivers of a child under 18 **n = 420 Age of caregivers (year)**: M = 35.53 (SD = 6.11, Range =23-58) **Gender of caregivers**: 202 (48.1%) females, 218 (51.9%) males. **Age of the focal child (year)**: birth to 5, n = 169 (40.2%), 6 to 11, n = 146 (34.8%), and 12 to 18, n = 105 (25%). **Race**: 303 (72.1%) White and 117 (27.9%) were of ethnic/racial minority including 45 (10.7%) Black/African American respondents, 71 (16.9%) Asian/Asian American respondents, 14 (3.3%) Hawaiian/Pacific Islander respondents, and 38 (9%) of American Indian/Alaskan Native descent. **Ethnicity**: LatinX, n = 81 (19.3%), Non-LatinX, n = 339 (80.7%). **Partner status of caregivers**: Partnered, n = 357 (85%), and Non-partnered, n = 63 (15%). **Type of family**: Not reported. | From April 27^th^ to 28^th^ 2020, approximately 5 weeks after the first U.S. quarantines were advised. | **Quantitative design.**  **Data collected:** 1) The Burden Scale for Family Caregivers (BSFC‑s). 2) The Perceived Stress Scale—Child form.3) The Generalized Anxiety Disorder‑7 (GAD‑7). 4) The Major Depression Inventory (MDI). 5) The Child-Parent Relationship Scale (CPRS).  **Analytic approach:** Descriptive analysis, correlation analysis, independent samples T-tests, one-way ANOVA, and path analysis. |
| Russell et al. (2021), USA | To explore clinically concerning depression symptom trajectories for parents over 30 days early in the pandemic. | **Participants**: Parents **n = 156**  **Age**: M = 37.04 (SD = 8.50, Range = 22-65) **Gender**: 77 (50.0%) female, 78 (49.4%) male, and 1(0.6%) transgender.  **Race**: Black/African American, n = 20 (12.8%); Asian/Asian American, n = 12 (7.7%); Native Hawaiian/other Pacific Islander, n = 1 (0.6%); American Indian/Alaska Native, n = 5 (3.2%); White, n = 122 (78.2%); Other, n = 2 (1.3%)  **Ethnicity**: LatinX, n = 12 (7.7%); Non-LatinX, n = 144 (92.3%). **Marital status**: Married, n = 127 (81.4%); Single, n = 9 (5.8%); Divorced, n = 1 (0.6%); Widowed, n = 1 (0.6%); Cohabiting, but not married, n = 18 (11.5%). **Focal child age category**: Birth to 5 years old, n = 63 (40.4%); 6 to 11 years old, n = 45 (28.8%); 12 to 18 years old, n = 47 (30.1%). **Prior employment**: No, n = 10 (6.4%); Yes, part-time, n = 24 (15.4%); Yes, full-time, n = 122 (78.2%). **Current employment**: No, n = 21 (13.5%); Yes, part-time, n = 32 (20.5%); Yes, full-time, n = 103 (66.0%). **Finances to meet needs**: Not at all, n = 4 (2.6%); A little, n = 19 (12.2%) Moderately, n = 41 (26.3%); Mostly, n = 42 (26.9%); Completely, n = 50 (32.1%). | Baseline data were collected from parents on April 27–28, 2020, five weeks after the first U.S. quarantines began. Follow-up data were collected 30 days later. | **Quantitative design (longitudinal). Data collected:** 1) Demographic Characteristics. 2) Emotion Regulation of caregivers: The 18-item Difficulties in Emotion Regulation Scale-Short Form (DERS-SF). 3) Parent Stress: The 10-item Perceived Stress Scale (PSS). 4) Relationship Indicators: a. Child–parent relationship: The Child–Parent Relationship Scale (CPRS). b. Romantic partner relationship quality: The Couple Satisfaction Index (CSI-16). 5) COVID-Related Stressors: The Covid-19 Stressors Scale—Short Form (CSS-SF). 6) Anxiety: The Generalized Anxiety Disorder-7 scale (GAD-7). 7) Depression: The 12-item Major Depression Inventory (MDI).  **Analytic approach:** Descriptive analysis, t test, ANOVA, binary logistic regression analysis. |
| Schmidt et al. (2021), Germany | To examine on a day-to-day basis whether distance learning and parental involvement therein were associated with negative parent–child interactions as well as parent-reported positive and negative affect of themselves and their children above and beyond daily stressors during the Covid-19 pandemic. | **Participants**: Parents **n = 562**  **Age**: M = 42.79 years (SD = 6.12, Range = 25–63)  **Gender**: 489 (87.0%) mothers.  **Race/ethnicity:** Not reported. **Marital status**: married, n = 382 (68.0%); living with their nonmarital partner in one common Household, n = 64 (11.4%); separated/divorced, n = 64 (11.4%); single, n = 28 (5.0%); with a partner, but living in separate households, n = 14 (2.5%); widowed, n = 9 (1.6%). **Age of the youngest child living in their household**: M = 9.74 years (SD = 2.81, Range = 6–19).  **Gender of the youngest child**: 268 girls, 290 boys, and 4 with no information. **Educational phase of the youngest child**: 367 (65.3%) attended elementary school, 135 (24.0%) attended the academic tier of secondary school (gymnasium). **Number of children living in the household**: M = 2.06 (SD = .82), with 301 (53.6%) participants having two children living in their household. **Highest level of education**: 344 (61.2%) had a university degree. **Net monthly household income**: 277 (49.2%) earned 4000e or more; 116 (20.6%) earned between 3000 € and 4000 €; 128 (22.8%) below 3000€ (41 participants did not provide information on income). | Using data of a 21-day diary study conducted between March 28, 2020 and April 27, 2020 in Germany, during which there were nationwide school closures, workplace closures in some sectors, and restrictions on gatherings to fewer than 10 people. | **Quantitative design (longitudinal).** **Data collected**: 1) Distance Learning: Whether children had received learning material or schoolwork from their school/teachers at that day. 2) Parental Involvement: The degree to which children learned independently or needed parental help. 3) Stressor Occurrence: Parents assessed whether any of the given events, that people could find irritating or disturbing, had occurred on that day. 4) Negative Parent–Child Interactions: Participants rated three items of negative interactions with their children on a 7-point Likert scale. 5) Parental Affect: Participants rated on a 7-point Likert scale the extent to which they experienced each of eight emotional states on that day. 6) Child Affect: Same items as for the assessment of parental affect.  **Analytic approach:** Descriptive analysis, association analysis, and multilevel structural equation models. |
| Schrooyen et al. (2021), Belgium | To examine the role of parental identity, as indicated by clear commitments and autonomous motivation for the parental role, in parents’ adaptation during the Covid-19 lockdown in Belgium. | **Participants**: Parents with a child under 18 **n = 492** **Age**: M = 43.97 years (SD = 7.52). **Gender**: 88.2% female.  **Race/ethnicity:** Not reported. **Number of biological children**: M = 2.08 (SD = 1.03).  **Family type**: 63.7% having an intact family with both parents present, 17.0% being a single parent, 10.1% formed a reconstituted family, 8.5% reported having another family structure (e.g., foster or adoptive family). **Educational level**: 31.2% obtained a university degree, 41.0% a college degree, and 22.5% did not attend higher Education, 5.3% did not provide this information.  **Income and employment status**: 82.4% reported having a sufficient income, and 64% working part time or full time. | From March 26 to April 24, during the lockdown period (schools, nonessential shops, and catering facilities were closed and people had to avoid contact with other people as much as possible). | **Quantitative design (longitudinal).**  **Data collected:**  1) Parental Identity were assessed by three indicators. a. Parental commitment and b. parental rumination: An adapted version of the well-validated Dimensions of Identity Development Scale1 (DIDS). c. Parents’ autonomous motivation: Three items from the Parenting Motivation Scale.  2) Parental Psychological Needs: An adjusted 12-item version of the Basic Psychological Need Satisfaction and Need Frustration.  3) Parenthood Experiences: a. Negative parenthood experiences: Three items from the Parental Burnout Assessment, one rating the amount of parenting stress they currently experience and the other one about a need for professional help. b. Positive parenthood experiences: Three face valid items regarding positive feelings that parents may experience when spending time with their children.  4) General Subjective Well-Being and Ill-Being (administered twice, with on average a 2-week interval between the two surveys): a. The shorted State Trait Anxiety Inventory and tHE 6-item version of the Centre for Epidemiological Studies— Depression scale. The items of anxiety and depression were averaged in a total score for ill-being. b. Subjective well-being: The most face valid item from the Satisfaction with Life Scale.  5) Cumulative Risk was indexed by three factors: a. Characteristics of the family itself. b. Characteristics of the children. c. The context of the family. The cumulative risk was constructed by dichotomizing each retained risk factors (0 = no risk; 1 = risk) and by summing these dichotomous scores.  **Analytic approach:** Descriptive analysis, correlations analysis, t test, multivariate analysis of covariance, and path analysis (using R-package “Lavaan”). |
| Shockley et al. (2021), USA | To investigate whether, in the Covid-19 context, couples are creating novel work-family (WF) management strategies that transcend gender roles or if they are falling back on the familiar patterns seen in previous research during “typical” times, and to examine the relationship of these strategies with men and women’s job performance and well-being (including health and family functioning). | **Participants**: Dual-earner couples with young children (under age 6) **n = 274 dual-earner couples** with young children  **Time 1 (n = 274)** **Age (year)**: 34.81 (SD = 5.78) for wives, 36.33 (SD = 6.60) for husbands.  **Race/Ethnicity**: White, 78.6% for wives, 76.7% for husbands; Hispanic/Latino, 6.2% for wives, 7.6% for husbands; Black/African American, 5.8% for wives, 6.1 for husbands; Asian, 6.6% for wives, 7.1% for husbands; American Indian/Alaskan Native, 1.2% for wives, 0.5% for husbands; Other, 1.6% for wives, 1.8% for husbands.  **Number of children** **aged younger than 6**: 1.36 (SD = .56).  **Number of children aged older than** **18:** 1.84 (SD =.82). **Weekly work hours**: 41.27 (SD = 5.66) for wives, 44.17 (SD = 7.88) for husbands.  **Household income** (USD): 140,702.70 (SD = 72,147.89).  **Time 2 (n = 133)** **Age (year)**: 35.17 (SD = 6.37) for wives, 36.34 (SD = 6.96) for husbands.  **Race/Ethnicity**: White, 78.7% for wives, 78.2% for husbands; Hispanic/Latino, 7.4% for wives, 7.3% for husbands; Black/African American, 4.1% for wives, 6.4% for husbands; Asian, 9.0% for wives, 7.3% for husbands; American Indian/Alaskan Native, 0.8% for wives, 0 for husbands; Other, 0 for wives, 0.9% for husbands.  **Number of children** **aged younger than 6**: 1.34 (SD =.54).  **Number of children aged older than** **18**: 11.74 (SD =.75). **Weekly work hours**: 40.68 (SD = 4.70) for wives, 44.02 (SD = 8.29) for husbands. **Household income** (USD): 139,402.60 (SD = 81,745.43). | Time 1: March 18–23, 2020. This was immediately following the closings of most schools and day-care facilities across the United States, and the declaration of a national health emergency on March 13.  Time 2: May 7–18, 2020. At this point in time, many states had only just begun their initial phases of reopening and schools and the majority of childcare facilities remained closed. | **Quantitative design (longitudinal). Data collected**: Both spouses completed all items. For Time 2 measures participants were asked to reflect on the period since they took the first survey. 1) Plan for Managing Childcare and Work Commitments (Time 1): Three open-ended questions, e.g., “What is you and your spouse’s plan for dealing with both of your work commitments during the upcoming period when your normal childcare is disrupted? Please be as detailed as possible.” 2) Implementation of Plan (Time 2): Participants rated how well they followed that plan on a 10- point Likert scale. 3) Family Functioning (Time 2): Family cohesion and marital relationship tension was assessed using instruments taken from previous research. 4) Health: Sleep quantity (hours for sleep at night) and psychological distress (a measure drew from previous research). 5) Job Performance: Quality of job performance ((a measure drew from previous research) and the extent to which participants felt they had met their work responsibilities during this period.  **Analytic approach:** Content analysis for qualitative data; Chi-square tests and latent class analysis for quantitative analysis. |
| Shorer and Leibovich (2020), Israel | To examine the contribution of parental emotion regulation and parental playfulness to young children’s stressogenic reactions to exposure to the Covid-19 crisis. | **Participants**: Israeli parents of children aged 2-7 years **n = 351**  **Age of parents**: M = 37.92 years (SD = 5.25, Range = 25-52). **Age of children**: M = 4.82 years (SD = 1.69, Range = 2-7).  **Gender of parents**: 309 (88.0%) mothers and 41 (11.7%) fathers.  **Gender of children**: 174 (49.6%) male and 176 (50.1%) female.  **Race/ethnicity of parents**: 97.4% Jewish. **Parents’ level of education** (years): M = 16.71 (SD = 2.25). **Marital status**: 87.2% married. **Income**: 6.8% reported below average income, 31.9% reported average income, and 55.3% reported above average income. | During one week of the Covid-19 outbreak in Israel but not reporting the exact date for data collection.  At this time, the Israeli school system had already been shut down for two weeks, most people were working from home, and leaving the house was permitted only for essential purposes. | **Quantitative design. Data collected**: 1) Children’s stress reactions before the Covid-19 outbreak: The Stress Reaction Checklist (SRCL, 15 items). 2) Children’s exposure to stressful Covid-19 related situations and information: An adapted 6-item version of the SRCL Exposure Scale (e.g., a close acquaintance who is sick with Coronavirus, experiencing lockdown, etc.) 3) Parents’ emotion regulation: The Difficulties in Emotion Regulation Scale (DERS). 4) Parental Playfulness: The Parental Playfulness Questionnaire (PPQ, 20 items) for parents of children aged 2–8.  **Analytic approach:** Descriptive analysis, Pearson correlation tests, T-test for independent samples, linear regression model, and mediation model analysis. |
| Spinelli et al. (2020), Italy | To explore how pandemic-related variables, structural aspects of the home and family environment, and parental subjective experience of stress and adjustment to the quarantine, affect the wellbeing of parents and children, and how in turn the well-being of parents and children are associated. | **Participants:** Parents of children aged 2-14 **n = 854 Age of parents (year)**: mothers, M = 38.96 (SD = 6.02); fathers, M = 41.9 (SD = 6.75). **Age of the focal child (year)** M = 7.14 (SD = 3.38, Range =2-14). **Gender of parents**: 797 females, 57males. **Race/ethnicity**: Not reported. **Educational level of mothers:** 49% had a high school degree or less, 37% had a bachelor’s or master's degree, and 21% had a higher education degree. **Educational level of fathers**: 41% had a high school degree or less, 33% had a bachelor’s or master's degree, and 26% had a higher education degree. **Location of residence**: 271 parents lived in the north of Italy where most Covid-19 cases, were registered (i.e., Lombardia and Veneto, the Red Area). **Type of family:** Not reported. | From April 2^nd^ to 7^th^, 2020.  The first restrictive measures started on February 21^st^ in the regions of Lombardia and Veneto, the areas most affected by the infection. On March 9^th^ the restrictions were extended to the entire national territory and ended on May 3^rd^. | **Quantitative design.** **Data collected:** 1) COVID-Contact Risk Index: An ad-hoc index to evaluate the amount of contact the parent had with people directly affected by the virus. 2) Home Environment Risk Index: An ad-hoc risk index to evaluate the house and family situation. 3) Quarantine Parent Risk Index: A newly developed pool of 13 items, 7-point Likert scale. 4) Parent’s Dyadic Parenting Stress: The Parent/Child Dysfunctional interaction domain of the Parenting-Stress Index Short Form (PSI). 5) Parent’s Individual Stress: The Stress subscale of the Depression Anxiety Stress Scale–Short form (DASS). 6) Children’s Psychological Problems: The parent-report form of the Strengths and Difficulties Questionnaire (SDQ).  **Analytic approach:** Descriptive analysis, correlation analysis, and multivariate regression models (using R). |
| Spinelli et al. (2021), Italy | To investigate factors related to the outbreak and to the household condition of the family that may contribute to parenting stress, and how this, in turn, was predictive of parental involvement in the child’s everyday activities and child’s emotion regulation adjustment. | **Participants**: Parents of children aged 2-14 years **n = 810 Age of parents**: Mothers, M = 39.09 years (SD = 5.98); fathers, M = 41.9 years (SD = 6.68), **Age of children**: M = 7.16 years (SD = 3.34). **Gender of parents**: 93% mothers, 7% fathers. **Gender of children**: 50% boys.  **Race/ethnicity of parents or children**: Not reported. **Educational level of mothers**: Mothers: 6% less than high school degree, 39% high school degree, 34% bachelor or master degree, 20% higher education degree.  **Educational level of fathers**: 2% less than high school degree, 36% high school degree; 38% bachelor or master degree, 24% higher education degree. **Number of children**: 32.4 % had one child and 52.7 % two children.  **Region**: 45% from the North, 37% from the Centre, and 18% from South of Italy.  **Socioeconomic risk status**: 185 (22.84%) participants were in the SES at-risk group. Of these, 58% lost the job due to the pandemic. | April 2–7, 2020, during the lockdown when everyone was banned from leaving home except for non-deferrable work or health reasons or other urgent matters. | **Quantitative design. Data collected:** 1) Socioeconomic risk index was calculated based on the potential risk factors, including loss of job due to the pandemic, total family income less than 1250 € per month, parent education level lower than high school. 2) Quarantine parent risk index was assessed by a newly developed pool of 13 items (e.g., finding space and time for themselves, the partner, and kids, balance family and work, etc.). 3) Parent dyadic parenting stress: The subscale Parent-Child Dysfunctional Interaction of the Parenting Stress Index Short Form (PSI). 4) Parent involvement with the child: Seven items of the Family involvement subdomain of the Parent Report Form CHIP-Child Edition. 5) Household chaos: A shortened version of the CHAOS-Chaos, Hubbub, and Order Scale. 6) Children’s emotion regulation: The Italian version of the 24-item Emotion regulation checklist (ERC).  **Analytic approach:** Descriptive analysis, correlation analysis, and multivariate mediation model. |
| Sun et al. (2021), USA | Using daily diary data to prospectively examine whether and how both school closure as well as days since school closure were related to sibling positivity and negativity among Latinx children. | **Participants**: Latinx children **n = 215** (from 116 families)  **Age of children**: M = 9.72 years (SD = 1.22, Range = 6.8-12.2) **Race/ethnicity of children**: Mexican (88%), Guatemalan (5%), Puerto Rican (<1%), and Salvadoran (<1%), or a combination of two Latinx subgroups (6%). **Nature of siblings**: biological siblings (90%), half siblings (9%), step siblings (< 1%), or cousins (<1%). **Nature of mother figures**: biological mothers (97%) or other female caregivers (3%, e.g., stepmothers, grandmothers). Nature of father figures: biological fathers (74%), stepfathers (13%), or other male figures (13%). **Residence of children**: Residing with two biological parents (74% for older siblings; 78% for younger siblings) or with biological mothers and stepfathers (17% for older siblings; 15% for younger siblings). **Sibship size** (i.e., total number of children living in the household at least 50% of the time): 37.6% families had 3 children, 27.3% had 4 children, 12% had 5 children, 11.1% had 2 children, and the remainder 12% had 6 or more children (Range = 2 -11). **Ethnicity of parents**: 92% mother and 96% father figures were Latinx.  **Race of parents**: Mother and father figures identified as White (47% and 48%, respectively) or “other” (49% and 51%, respectively), with the remainder (4% and 1%, respectively) racially identifying as Native American, Asian, Black, or Hawaiian. Most parents who selected “other” for race specified a panethnic (i.e., Latino/a, Hispanic) or ethnic (e.g., Mexican) category. **Parents’ place of birth outside the U.S.:** 68% and 81%, for mother and father figures respectively. **Parents’ education level**: 53.9% of mothers and 69.4% of fathers had less than a high school education, and 29.6% of mothers and 19.4% of fathers had a GED or high school degree, with the remainder having some college or a degree (associate’s, bachelor’s, or master’s). **Employment status** at T1: 96.6% fathers and 49% mothers were employed. **Annual median family income at T1**: $27,600 (SD = $24,353) for an average of 6.18 (SD = 1.86) people living in the home, with 88% of households receiving some form of public assistance. | Time 1 (pre-pandemic): Fall 2019.  Time 2 (during the outbreak): February to May 2020, all 10 participating schools closed on the same day (March 16, 2020). | **Quantitative design (longitudinal).**  **Data collected:** 1) Covid-19 School Closure (T2): a. School not closed or school closed due to Covid-19. b. Days since school closure. 2) Daily Sibling Relationship Positivity and Negativity (T2): Children reported on their positive and negative Behaviours toward their participating siblings, from the time they woke up until the time of the call, during each of the seven nightly phone calls using a 3-point scale. 3) Moderators (T1) including family SES scores, parents’ education level, family income, sibship size, child enculturation and global ratings of sibling positivity and negativity.  **Analytic approach:** Descriptive analysis, association analysis, t-test, and multilevel tobit regression models. |
| Taha et al. (2022), USA | To explore the relationships among sociodemographic characteristics, parental worrying, family functioning, and child health-related quality of life (HRQoL) while primarily focusing on the direct and interaction effects of parental worrying and family functioning on child HRQoL. | **Participants**: Parents and their children **n = 93** parent-child dyads **Age of parents**: M = 46 years (SD = 8.1) **Age of children**: M = 14 years (SD = 2.3) **Gender of parents**: 77 (82.8%) females, 16 (17.2%) males. **Gender of children**: 50 (53.8%) females, 43 (46.2%) males.  **Race/ethnicity of parents**: White, n = 68 (73.1%), Black, n = 3 (3.2%); Hispanic/Latino, n = 6 (6.5%); Asian 6 (6.5%); Pacific Islander, n = 2 (2.2%); Middle Eastern, n = 8 (8.6%). **Parental education**: Graduate degree, n = 49 (52.7%); 4-year degree, n = 28 (30.1%) Some college, n = 9 (9.7%); High school, n = 6 (6.5%); Below high school, n = 1 (1.1%). **Total household annual income before taxes**: $200,000 or more, n = 30 (32.3%) $199,999–$150,000, n = 12 (12.9%); $149,999–$100,000, n = 21 (22.6%); $99,999–$75,000, n = 8 (8.6%); $74,999–$50,000, n = 13 (14.0%); $49,999–$35,000, n = 4 (4.3%); $34,999–$25,000, n = 4 (4.3%); $24,999–$15,000, n = 1 (1.1%). **Location of residence**: The West Coast in Oregon (n = 49), California (n = 37), Hawaii (n = 3),Washington state (n = 1), Arizona (n = 1), Minnesota (n = 1), and Illinois (n = 1). **Compliance with Covid-19 state guidelines**: Always, n = 42 (45.2%); Often, n = 45 (48.4%); Sometimes, n = 4 (4.3%); Rarely, n = 1 (1.1%); Never, n = 1 (1.1%). | From December 2020 to February 2021, during the Covid-19 pandemic (not clear whether there were any restrictions during this period). | **Quantitative design. Data collected:** 1) Sociodemographic characteristics. 2) Family functioning (General Functioning Scale–Family Assessment Device, GF-FAD). 3) Parental worrying (Worry Domains Questionnaire, WDQ). 4) Child health-related quality of life (KIDSCREEN-10, KS10). Parents completed the GF-FAD and WDQ measures while the children completed the KS10 measure.  **Analytic approach:** Descriptive analysis, association analysis, and hierarchical linear regression analysis. |
| Tang et al. (2021), mainland China | To estimate the prevalence of depressive, anxiety, and stress symptoms, and levels of life satisfaction, among children and adolescents experiencing home quarantine and school closure in Shanghai; identify factors related to their mental health status, examining the role of the perceived impact of home quarantine and parent-child discussion relating to mental health status during the Covid-19 pandemic. | **Participants:** Primary and secondary school students **n = 4,342 Age of students (year):** M = 11.86 (SD = 2.32, Range = 6-17) **Gender of students:** 49% females, 51% males. **Race/ethnicity:** Not reported. **Type of family:** Not reported. | During March 13^rd^ - 23^rd^, 2020 (Shanghai launched its highest-level emergency response on January 24, 2020, and the level 1 emergency response had been active until March 23, 2020, when all public venues were closed and all large-scale public events were cancelled). | **Quantitative design. Data collected:** 1) Depression, anxiety, and stress scale (Chinese version of DASS-21). 2) Life satisfaction (current life satisfaction and a change in life satisfaction since the pandemic). 3) Perceived impact of home quarantine: A self-constructed questionnaire by authors. 4) Parent-child discussion on Covid-19 (whether parents had discussed Covid-19 with participants and the frequency of discussion).  **Analytic approach:** Descriptive analysis, Chi-square tests, independent sample t-tests, one-way analyses of variance (ANOVA), multivariate analyses of variance (MANOVA), binary logistic regression analysis, and linear regression analysis. |
| Uzun et al. (2021), Turkey | To investigate the factors affecting the relationship between parents and their children aged 4-6 in the Covid-19 process. | **Participants**: Parents with children aged 4–6 **n = 219** **Age of mothers**: 11% of the mothers were between age 23 and 27, 34.2% between age 28 and 32, 33.8% between age 33 and 37, 20.1% between age 38 and 43, and 0.9% between age 44 and 48. **Age of fathers**: 0.9% of the fathers were between the ages of 23–27, 15.5% between 28–32, 37.4% between 33–37, and 32.9% between 38–43, 12.3% between 44–48, and 0.9% are 49 and older.  **Race/ethnicity of parents**: Not reported. **Educational level of mothers**: 18.7% have graduated from primary school, 13.2% have graduated from middle school, 18.3% have graduated from high school, 39.7% have graduated from college or university, 10% graduated from Graduate school.  **Employment status of mothers**: 82 of them are employed, 129 of them are unemployed and 8 of them are employed occasionally. **Educational level of mothers**: 9.6% of the fathers have graduated from primary school, 13.2% have graduated from middle school, 23.3% have graduated from high school, 40.2% have graduated from college or university, 13.7% have graduated from graduate school.  **Employment status of fathers:** 85.5% of them are employed, 5.5% of them are unemployed, 8.7% of them are employed occasionally. **Childcare status during quarantine process**: 155 (70.5%) parents took care of the child during quarantine process. **Quarantine status**: 184 (84%) parents have been in quarantine for 31 days and more and during this process 81 (37%) parents had gone out alternately in case of necessity. Father is more likely to have to go out due to reasons such as work etc., n = 94 (42.9%). | Between April and June 2020. Not clear if any restrictions were imposed during this time period. | **Quantitative design. Data collected:** 1) Demographic Information. 2) The Turkish form of Child-Parent Relationship Inventory (PCRI), including 7 sub-dimensions, communication, discipline, support, satisfaction, autonomy, participation, and role.  **Analytic approach:** Descriptive analysis, t-test and one-way variance analysis (One-Way ANOVA). |
| Verweij et al. (2021), the Netherlands | To examine whether, to what extent and for whom (by sex and educational attainment) work-to-family conflict (W→F-conflict) and family-to-work conflict (F→W-conflict) increased from the pre-Covid-19 period to the first lockdown period, and to investigate whether and to what extent the negative associations between W→F-conflict/F→W-conflict and perceived parenting became stronger. | **Participants**: Employed parents with children aged 3 **n = 131** parents  **Age of children:** M = 3.5 years at wave 1, M = 4.7 years (57 months) at wave 2 (during the Covid-19 lockdown) **Gender of parents**: 55 mothers and 76 fathers. **Race/ethnicity of parents**: Not reported. **Educational level of mothers** (n = 55): high, n = 26 (47.27%), low/medium, n = 29 (52.73%). **Working hours of mothers**: 26.25 (SD = 8.81) at wave 1, 21.35 (SD = 10.35) at wave 2. **Partner’s working hours of mothers**: 39.23 (SD = 15.72) at wave 1, 31.72 (SD = 14.32) at wave 2. **Educational level of fathers** (n = 76): high, n = 37 (48.68%), low/medium, n = 39 (51.32%). **Working hours of fathers**: 40.40 (SD = 11.78) at wave 1, 35.30 (SD = 10.06) at wave 2. **Partner’s working hours of fathers**: 21.26 (SD = 13.54) at wave 1, 14.86 (SD = 12.79) at wave 2. | Wave 1: Between May 2018 to January 2020.  Wave 2: During the first Covid-19 lockdown between April 15 and May 11 2020. During this period, all schools and day-care centres were closed, and other informal forms of childcare, such as care by grandparents, were discouraged. Also, from March 2020 onwards, the Dutch government recommended employees to work from home as much as possible. | **Quantitative design (longitudinal).** Data collected: Wave 1: Home-based observation and online survey (using a tablet). Wave 2 (during the lockdown period): online survey (through email) 1) W→F-conflict (T1 & T2): The work-family conflict scale. 2) F→W-conflict (T1 & T2): The other half of the work-family conflict scale. 3) Perceived parenting (T1 & T2): Three subscales of Parenting and Family Adjustment Scale (PAFAS), namely coercive discipline, positive encouragement, and the parent-child relationship. 4) Parental highest educational level: Lower/medium education (primary school, secondary school, or intermediate vocational training) or high education (higher vocational training or (post) university degree).  **Analytic approach:** Descriptive analysis, association analysis, and multilevel regression analysis. |
| Waller et al. (2021), USA | To examine associations between family characteristics and exposure to, and worries about, Covid-19 and to explore whether established links between parenting and Conduct Problems (CP) and  Callous Unemotional (CU) traits persist in the context of heightened familial stress; whether parental Covid-19 exposures and worries were related to parenting behaviours; examined whether parent reports of risk exposures to, and worries about, Covid-19 directly impacted children’s CP and CU traits, over and above the impact of harsh and warm parenting practices. | **Participants**: Parents of children aged 3 to 10. **n = 303 Age of parents (year)**: M = 38.04 (SD = 5.21) **Relationship to the focal child**: biological mothers (92.4%), biological fathers (2.0%), foster or adoptive mother (5.0%), other female caregiver (0.7%). **Age of children (year)**: M = 6.43 (SD = 2.13, Range =3.00-10.94). **Gender of children:** 51.8% females, 48.2% males. **Race of parents**: White (n = 224; 73.9%), Black/African-American (n = 36; 11.9%, including n = 4 biracial), Asian (n = 30; 9.9%, including n = 5 biracial), and other (n = 13; 4.3%).  **Race of children**: White (n = 199; 65.7%), Black/African-American (n = 49; 16.2%, including n = 13 biracial), Asian (n = 37; 12.2%, including n = 22 biracial), and other (n = 18; 5.9%). **Ethnicity**: 13 parents (4.3%) and 31 children (10.2%) were reported to be Hispanic/Latino/a. **Educational level of parents**: 60.5% of parents reported having a graduate-level degree, 24.4% had a Bachelor level degree, 4.7% had an Associates level qualification, and 10.4% had a high school qualification or less.  **Type of family**: Not reported. | From April to July 2020 (encompassing a period with high community transmission and stay-at-home orders due to Covid-19) | **Quantitative design.** **Data collected**: 1) Covid-19 Exposures and Worries. 2) Child Conduct Problems (CP): The 5-item CP subscale of the Strengths and Difficulties Questionnaire (SDQ). 3) Child Callous‑Unemotional (CU) Traits: The 24-item Inventory of Callous-Unemotional Traits (ICU). 4) Parental Warmth: The 6-item positive involvement subscale of the Alabama Parenting Questionnaire (APQ). 5) Harsh Parenting: The 9-item over-reactivity subscale of the Parenting Scale.  **Analytic approach:** Descriptive analysis, correlation analysis, and regression analyses within a structural equation modeling framework. |
| Walters et al. (2021), USA | To examine whether the Covid-19 pandemic and associated restrictions have had an effect on the social and psychological well-being of Pennsylvania schoolchildren. | **Participants**: Middle school students **n = 309** **Age**: 12.38 years (SD = 0.98, Range = 10–16). **Grade of students**: sixth (29.2%), seventh (27.5%), and eighth (43.3%) grades. **Gender**: 141 females, 134 males, and 34 failed to list their sex on the questionnaire.  **Race/ethnicity**: 44.5% white, 21.8% black, 20.5% Hispanic, and 13.3% other (total student population: white = 51%, black = 23%, Hispanic = 21%, other = 5%). **Live arrangement**: 68.9% lived with both biological or adoptive parents, 17.1% lived with a biological/adoptive parent and a stepparent, 12.0% lived with one biological parent only, and 1.4% lived with grandparents or other relatives. **Housing conditions**: 84.2% reported living in a single-family dwelling (84.2%), another 10.0% reported living in a duplex or condominium, and 5.8% indicated that they were living in an apartment.  Longitudinal analyses were performed on a subset of participants: specifically, 174 students (85 boys, 89 girls) who completed the survey in both 2019 and 2020. | Time 1: November 2019. Time 2: November 2020 (nine months after the start of the pandemic).  On March 13, 2020, the governor closed all schools for two weeks, and eventually cancelled all in-person instruction for the rest of the 2019–2020 academic year. The Fall 2020–2021 semester was subsequently modified so that students spent some, if not all, of their time in virtual learning. | **Quantitative design (longitudinal). Data collected:** 1) Parental Support: The 7-item Support scale from the Quality of Relationships Inventory (QRI). 2) Parental Knowledge: The 8-item Parental Management Scale (PMS). 3) Peer Deviance: A three-item scale (e.g., “how many of your four closest friends have taken a drink of alcohol in the last year”). 4) Neutralization: The 11-item Denver Youth Survey (DYS) Neutralization scale. 5) Cognitive Impulsivity: The 8-item Weinberger Adjustment Inventory Impulse Control scale (WAI-IC). 6) Depression: The Centre for Epidemiological Studies Depression scale (CES-D). 7) Delinquency: The 14-item Self-Reported Offending (SRO) scale. 8) Bullying Victimization: Six items from the Middle School and Pennsylvania Youth surveys. 9) Bullying Perpetration: A self-report measure of bullying perpetration adapted from the combined Middle School Survey and Pennsylvania Youth Survey.  **Analytic approach:** Descriptive analysis, association analysis, and t test. |
| Wang (2021), mainland China | To contribute to the urgent discussion through exploring the social emotional development of young children during the domestic quarantine and the factors which contribute to or mitigate the negative effects of quarantine on children’s psychological well-being. | **Particip**  **ants**: Parents of preschoolers **n = 31**  **Age of children**: M = 59.06 months (SD = 3.27). **Gender of children**: 52% males. **Number of children**: 61% have only one child. **Domestic quarantine days**: M = 48.58 (SD = 14.85). **Community type**: 45.16% rural buildings or resettlement housing, 9.68% danwei apartment or affordable housing or public rental housing, 45.16% general commercial housing. [Note. Danwei is a work unit in China and danwei apartment is a type of community where people from the same work unit live in the same neighbourhood] **Race/ethnicity:** Not reported. | T1 (pre-lockdown): Jan 10 to 12, 2020.  T2 (during lockdown): March 13 to 17, 2020.   The central government of China mandated an unprecedented lockdown on 23 January 2020. Outdoor restrictions were implemented and residents were requested to stay at home except for special reasons. The mandatory domestic quarantine lasted for 76 days ending on 8 April 2020 in Wuhan. | **Mixed-method design (longitudinal). Data collected:**  1) Demographics. 2) Parental distress: the Hopkins Check List (HSCL- 10). 3) Parent-child relationship: A simplified version (7-item) of the Chinese version of Parental Caring Stress Scale. 4) Children’s social emotional competence: The Ages & Stages Questionnaires: Social–Emotional Second Edition (ASQ:SE-2). 5) Children’s general developmental: A Chinese version of the Ages & Stages Questionnaires, Third Edition (ASQ-C). **Analytic approach:** For quantitative analysis, using t-test and ordinary linear regression analysis. For qualitative analysis, not explicitly reporting the method for qualitative analysis (only reported using coding approaches). |
| Westrupp et al. (2021), Australia | To investigate the impact of Covid-19 on the health and wellbeing of parents, children, and families, and identifies risk factors that are associated with poorer outcomes in the context of large-scale public health crises. | **Participants**: Parents of a child aged 0–18 years **n = 2,365**  **Age of parents**: M = 38.30 years (SD = 7.07). **Age of children**: M = 8.66 years (SD = 5.14). **Gender of parents**: Cisgender men, 19.2%; Cisgender women, 80.7%; Transgender or non-binary, 0.1% **Gender of children**: Cisgender boy, 51.0%; Cisgender girl, 48.6%; Transgender or non-binary, 0.4%.  **Parent born overseas**: 18.0%.  **Language other than English**: 4.6%.  **Aboriginal or Torres Strait Islander**: 2.0%. **Number of children**: one child, 28.4%; two children, 46.0%; three children, 18.2%; four or more children, 7.3%. **Geographic location**: Major Cities of Australia, 70.1%; Inner Regional Australia, 22.8%; Outer Regional Australia, 6.1%; Remote Australia, 1.0%. **Low household income**: 14.1%. **Receiving government benefit**: 5.8%. **Single parent household**: 11.0%. **Did not complete high school**: 9.4%. **Deprivation index**: M = 0.38 (SD = 0.96). | During ‘a ‘level three’ national lockdown in Australia in April 2020.  The restrictions required that Australians avoid leaving their house except for four reasons: (1) shopping for food and supplies, (2) care and caregiving, (3) exercise, and (4) study or work—if unable to do so from home. | **Quantitative design.**  **Data collected**:  1) Participant demographics and characteristics, including a. Pre-pandemic family demographics; and b. Individual parent and child factors.  2) Covid-19 stressors: a. Covid-19 psychological stressors: Participants reported their feelings about catching COVID- 19, Covid-19 as serious health risk, and their perceptions of dealing with Covid-19. b. An index of Covid-19 environmental stressors summed seven items (e.g., financial insecurity, job loss, reduction in work hours, etc.). c. The frequency of use of news media during the pandemic. d. Whether parents had a child at home while working. e. Housing (renting, equivalized number of bedrooms, satisfaction with quality of house, and size of private outdoor space).  3) Parent outcomes: a. The Depression, Anxiety and Stress Scales-21. b. The Difficulties in Emotion Regulation Scale-16. c. Frequency of alcohol consumption. d. Frequency of smoking.  4) Child and family outcomes: a. Child depression: The Short Mood and Feelings Questionnaire. b. Child anxiety: Four selected items from Brief Spence Children’s Anxiety Scale. c. Parenting irritability: Four items from the Longitudinal Study of Australian Children (LSAC). d. Couple verbal conflict: Four-item Argumentative Relationship Scale from LSAC.  **Analytic approach:** Descriptive analysis, t test, chi-squared test, sensitivity analyses and linear regression analysis. |
| Wolf et al. (2021), USA | To examine whether parental stress is associated with use of punitive parenting, as well as whether this association is modified by drinking pattern. | **Participants**: Parents **n = 329**  **Focal child age**: M = 6.18 years (SD = 3.05). **Child biological sex**: 143 (43.5 %) females, 186 (56.5 %) males. **Child race/ethnicity**: White, n = 255 (77.5 %); African American/Black, n = 41 (12.5 %); Other Race or Ethnicity, n = 33 (10.0 %). **Caregiver biological sex**: 305 (92.7%) females, 24 (7.3%) male **Number of children**: M = 2.14 (SD = 0.96) **Marital status**: Single/Widowed/Divorced, n = 46 (14.0 %); Married or living in marriage-like relationship, n = 283 (86.0 %). **Parent education**: Some college or less, n = 78 (23.7 %); Bachelor’s Degree, n = 114 (34.7 %); Graduate Degree, n = 137 (41.6 %). **Frequency of alcohol use**: Abstained from alcohol past 12 months, n = 33 (10.0 %); Drank at least once a year, n = 80 (24.3 %); Drank at least once a month, n = 61 (18.5 % ); Drank at least once a week, n = 155 (47.1 %). | Between April 13, 2020 and May 27, 2020. Ohio’s stay-at-home orders began on March 24, 2020 and most businesses were opened with restrictions by May 12, 2020. | **Quantitative design (longitudinal).** **Data collected**: Participants completed a 30–45-minute online survey at baseline; and complete ecological momentary assessment surveys by answering 3–5 questions during three time periods (10 a.m., 3 p.m., and 9 p.m.) each day for fourteen days using an app downloaded to their cellular telephone. 1) Punitive and non-punitive parenting Behaviours: The Dimensions of Discipline (DD). 2) At-the-moment stress: A 10-point scale measuring the stress during each daily observation period. 3) Frequency of alcohol use (baseline): How often participants had at least one alcoholic drink in the past twelve months (recoded into four categories: weekly drinkers, monthly drinkers, yearly drinkers, and abstainers). **Analytic approach:** Descriptive analysis, one-way ANOVA, chi-square bivariate analysis, and nested multilevel ordinal regression models. |
| Wong et al. (2021), Hong Kong, | To investigate the association between Covid- 19 and child maltreatment and whether job loss, income reduction, and parenting affect child maltreatment. | **Participants**: Parents having and living with a child or children aged under 10 years **n = 600** **Age (year)**: M = 38.1 (SD = 6.43) **Gender**: 416 (69.3%) females. **Race/ethnicity**: Not reported. **Educational level**: All had attained secondary/tertiary level or above. **Type of family**: Married or cohabited, and having and living with a child or children aged under 10 years. | Between 29 May to 16 June 2020. | **Quantitative design.** **Data collected: 1)** Child maltreatment during Covid-19: The Conflict Tactics Scale-Parent Child (CTSPC) scale. 2) Parental health literacy: 16-item European Health Literacy Questionnaire short form (HLS-EU-Q16). 3) Parent’s relationship with partner during Covid-19: The Abuse Assessment Screen. 4) Parent’s mental health distress: The Patient Health Questionnaire-4 (PHQ-4). 5) Other measurements of income instability and parent’s time staying at home.  **Analytic approach:** Descriptive analysis and logistic regression analysis. |
| Wu et al. (2020), mainland China | To investigate the mental health of students’ parents and its influence factors during the Covid-19 pandemic. | **Participants**: Parents **n = 1,163**  **Child's learning stage**: Primary school, n = 299 (25.7%); Middle school, n = 354 (30.4%); High school, n = 307 (26.4%); College, n = 203 (17.5%) **Gender of parents**: 230 males (19.8%) and 933 females (80.2%). **Domicile**: 162 parents (13.9%) from central China and 1001 (86.1%) from non-central China.  **Marital status of parents**: 1098 parents of married or remarried (94.4%) and 65 divorced or widowed parents (5.6%). **Occupation of parents**: Most of the parents were clerks (39.9%) and professional and technical workers (25.7%). **Quarantine status**: 146 (12.5%) parents themselves or their family members had been quarantined for 2 weeks.  **Parents’ history of mental illness**: 93 (8%) of the parents had a history of mental illness.  **Family economic leve**l: 847 (72.8%) of parents had a medium or high economic level. | In March 2020 (the study did not indicate whether there were any restrictions taken during this period in China). | **Quantitative design.**  **Data collected:** 1) Demographic information. 2) Depressive symptoms: The Patient Health Questionnaire-9 (PHQ-9). 3) Anxiety symptoms: The Generalised Anxiety Disorder-7 (GAD-7). 4) Perceived Stress Scale-10 (PSS-10). 5) Social Support Rating Scale (SSRS).  **Analytic approach:** Descriptive analysis, one-way analysis of variance (ANOVA), Spearman’s correlation, and regression analysis. |
| Xu et al. (2020), USA | To examine relationships between parenting stress, mental health, and risky parenting Behaviours among grandparent kinship caregivers during Covid-19, and to investigate whether grandparent kinship caregivers’ mental health is a potential mediator between parenting stress and caregivers’ risky parenting behaviours. | **Participants**: Grandparent kinship caregivers  **n = 362** **Age of grandparents**: M = 56.5 years (SD = 7.75, Range = 42–90). **Age of children**: M = 9.53 years (SD = 4.68, Range = 0-19). **Gender of grandparents**: Male, n = 136 (37.57%); Female, n = 226 (62.43%). **Child gender**: Male, n = 195 (54.02%); Female, n = 166 (45.98%).  **Grandparent race:** White, n = 246 (68.72%); Non-White, n = 112 (31.28%). **Grandparent education**: Below college, n = 218 60.22%); College and above, n = 144 39.78%). **Grandparent marital status**: Married, n = 252 (69.61%); Other, n = 110 (30.39%). **Number of children in the household**: One child 64 (17.68%); More than one child, n = 298 82.32%). **Years of care**: One year or less than one year, n = 77 (19.15%); More than one year, n = 325 (80.85%). **Licensed kinship caregivers**: Yes, n =143 (39.61%); No, n =218 (60.39%). **Labor force status**: Full time, n =157 (43.37%); Part time, n =83 22.93%); Don’t work, n =122 (33.70%). **Lost job during the pandemic**: Yes, n =82 (22.71%); No, n =279 (77.29%). **Grandparent household income in 2019**: ≤30,000, n = 103 (29.28%); 30,000 - ≤60,000, n = 135 (37.29%); >60,000, n = 121 (33.43%). | In June 2020 (the study did not indicate whether there were any restrictions taken during this period in the US) | **Quantitative design. Data collected:** 1) Demographics of child and grandparents. 2) Psychological Aggression, Corporal Punishment, and Neglectful Behaviours: The three subscales of the Conflict Tactics Scales Parent-Child (CTSPC). 3) Grandparents’ parenting Stress during Covid-19: The shortened Parent Stress Index. 4) Grandparents’ mental Health: The Mental Health Inventory-5 (MHI-5) scale.  **Analytic approach:** Descriptive analyses, negative binomial regression analyses, and mediation analyses. |
| Zhang et al. (2021), mainland China | To examine family resources and resilience across four major groups of Chinese children (urban children, migrant children, rural non-migrant children, and rural left-behind children) during the pandemic, using data from a 2020 survey conducted in Shaanxi Province during the Covid-19 outbreak. | **Participants**: Middle school and high school students **n = 10,255** **Gender**: 48.76% female. **Age**: M = 15.22 years (SD = 1.60).  **Race/ethnicity**: Not reported. **Children’s migration status:** 74.78% urban, 8.37% rural non-migrant, 8.75% left-behind and 8.10% migrant children. | February 2020 (during the pandemic). This study did not report if there were any restrictions in Shanxi Province where this study conducted. | **Quantitative design. Data collected**: 1) Psychological well-being (outcome variable): Five items adapted from the Achenbach Youth Self-Report (YSR). 2) Parent–child relationships (PCR, 4 items): Two items on parent–child closeness and two on parent–child communication. 3) Family economic status (FES). 4) Children’s migration status: Urban children, non-migrant rural children, left-behind rural children, and migrant rural children. 5) Demographics.  **Analytic approach:** Descriptive analysis, random-effect (RE) and fixed-effect (FE) regression analysis, and lagged dependent variable (LDV) regression analysis. |
